# GestaltMatcher: Overcoming the limits of rare disease matching using facial phenotypic descriptors

**DOI:** 10.1101/2020.12.28.20248193

**Authors:** Tzung-Chien Hsieh, Aviram Bar-Haim, Shahida Moosa, Nadja Ehmke, Karen W. Gripp, Jean Tori Pantel, Magdalena Danyel, Martin Atta Mensah, Denise Horn, Stanislav Rosnev, Nicole Fleischer, Guilherme Bonini, Alexander Hustinx, Alexander Schmid, Alexej Knaus, Behnam Javanmardi, Hannah Klinkhammer, Hellen Lesmann, Sugirthan Sivalingam, Tom Kamphans, Wolfgang Meiswinkel, Frédéric Ebstein, Elke Krüger, Sébastien Küry, Stéphane Bézieau, Axel Schmidt, Sophia Peters, Hartmut Engels, Elisabeth Mangold, Martina Kreiß, Kirsten Cremer, Claudia Perne, Regina C. Betz, Tim Bender, Kathrin Grundmann-Hauser, Tobias B. Haack, Matias Wagner, Theresa Brunet, Heidi Beate Bentzen, Luisa Averdunk, Kimberly Christine Coetzer, Gholson J. Lyon, Malte Spielmann, Christian Schaaf, Stefan Mundlos, Markus M. Nöthen, Peter Krawitz

## Abstract

A large fraction of monogenic disorders causes craniofacial abnormalities with characteristic facial morphology. These disorders can be diagnosed more efficiently with the support of computer-aided next-generation phenotyping tools, such as DeepGestalt. These tools have learned to associate facial phenotypes with the underlying syndrome through training on thousands of patient photographs. However, this “supervised” approach means that diagnoses are only possible if the disorder was part of the training set. To improve recognition of ultra-rare disorders, we created GestaltMatcher, which uses a deep convolutional neural network based on the DeepGestalt framework. We used photographs of 17,560 patients with 1,115 rare disorders to define a “Clinical Face Phenotype Space”. Distance between cases in the phenotype space defines syndromic similarity, allowing test patients to be matched to a molecular diagnosis even when the disorder was not included in the training set. Similarities among patients with previously unknown disease genes can also be detected. Therefore, in concert with mutation data, GestaltMatcher could accelerate the clinical diagnosis of patients with ultra-rare disorders and facial dysmorphism, as well as enable the delineation of novel phenotypes.

## Introduction

Rare genetic disorders affect more than 6.2% of the global population^1^. Because genetic disorders are rare and diverse, accurate clinical diagnosis is a time-consuming and challenging process, often referred to as the “diagnostic odyssey,^2^” and all informative clinical features have to be taken into consideration. A large fraction of patients, particularly those with neurodevelopmental disorders, exhibits craniofacial abnormalities^3^. If the facial phenotype (“gestalt”) is highly recognizable, such as in Down syndrome, it may also play an important role in establishing the diagnosis. Sometimes the gestalt is so characteristic or distinct that it reduces the search space of candidate genes or can be used to delineate novel phenotype-gene associations^4^. However, the ability to recognize these syndromic disorders relies heavily on the clinician’s experience. Reaching a diagnosis is very challenging if the clinician has not previously seen a patient with an ultra-rare disorder or if the patient presents with a novel disorder, both of which are increasingly common scenarios.

With the rapid development of machine learning and computer vision, a considerable number of next-generation phenotyping tools have emerged that can analyze facial dysmorphology using two-dimensional (2D) portraits of patients^5–13^. These tools can aid in the diagnosis of patients with facial dysmorphism by matching their facial phenotype with that of known disorders. In 2014, Ferry *et al.* proposed using a Clinical Face Phenotype Space (CFPS) formed by facial features extracted from images to perform syndrome classification; the system in that study was trained on photos of more than 1,500 controls and 1,300 patients with eight different syndromes^5^. Since then, facial recognition technologies have improved significantly and constitute the core of the deep-learning revolution in computer vision^14,15^. The current state-of-the-art framework for syndrome classification, DeepGestalt (Face2Gene, FDNA inc, USA), has been trained on more than 20,000 patients and currently achieves high accuracy in identifying the correct syndrome for roughly 300 syndromes^12,16^. DeepGestalt has also demonstrated a strong ability to separate specific syndromes and subtypes, surpassing human experts’ performance^12^. Hence, pediatricians and geneticists increasingly use such next-generation phenotyping tools for differential diagnostics in patients with facial dysmorphism. However, most existing tools, including DeepGestalt, need to be trained on large numbers of photographs, and are therefore limited to syndromes with at least seven images of different patients. The number of submissions to diagnostic databases of pathogenic variants, such as ClinVar^17^, has become a good surrogate for the prevalence of rare disorders. When submissions to ClinVar of disease genes with pathogenic mutations are plotted in decreasing order, most of the supported syndromes are on the left, indicating relatively high prevalence (Figure 1). For instance, Cornelia de Lange syndrome (CdLS), which has been modeled by multiple tools^5,12^, is caused by mutations in *NIPBL*, *SMC1A*, or *HDAC8*, as well as other genes, and has been linked to hundreds of reported mutations. However, more than half of the genes in ClinVar have fewer than ten submissions each (Figure 1). As a result, most phenotypes have not been modeled because sufficient data are lacking. Thus, the need to train on large numbers of photographs is a major limitation for the identification of ultra-rare syndromes.

**Figure 1:**
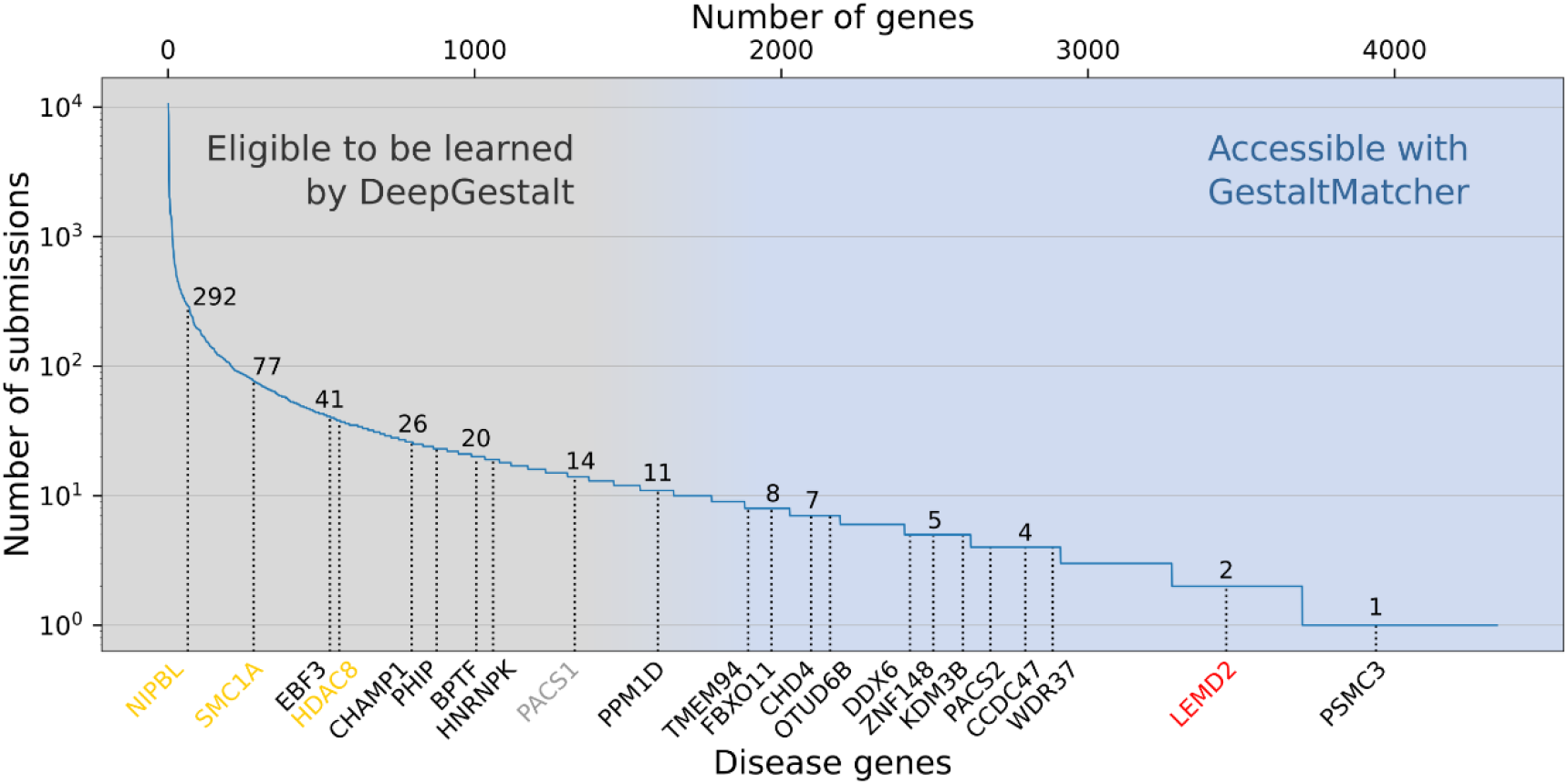
Subsets of disorders supported by DeepGestalt and GestaltMatcher. The lower x-axis shows examples of disease genes, and the upper x-axis is the cumulative number of genes. The y-axis shows the number of pathogenic submissions in ClinVar for each gene. The numbers on the curve indicate the number of submissions for each of the indicated genes. Most of the rare disorders that DeepGestalt supports have relatively high prevalence based on their ClinVar submissions, e.g. Cornelia de Lange syndrome (CdLS) which is caused by mutation in *NIPBL*, *SMC1A*, or *HDAC8*, among other genes. Disease genes such as *PACS1* cause highly distinctive phenotypes but are ultra-rare, representing the limit of what current technology can achieve. The first novel disease that was characterized by GestaltMatcher is caused by mutations in *LEMD2*. A candidate disease gene associated with a characteristic phenotype that can be identified by GestaltMatcher is *PSMC3*.

A second limitation of classifiers such as DeepGestalt is that their end-to-end, offline-trained architecture does not support new syndromes without additional modifications. In order to model a new syndrome in a deep convolutional neural network (DCNN), the developer has to go through six separate steps (Supplementary Figure 1), including collecting images of the new syndrome; changing the classification head, which is the last layer of the DCNN; retraining the network; and more. In addition, the model cannot be used to quantify similarities among undiagnosed patients, which is crucial in the delineation of novel syndromes.

A third shortcoming of current approaches is that they are not able to contribute to the longstanding discussion within the nosology of genetic diseases about distinguishability. Syndromic differences have been hard to measure objectively^18^, and decisions to “split” syndromes into separate entities on the basis of perceived differences or to “lump” syndromes together on the basis of similarities have been made subjectively. Current tools are unable to quantify the similarities between syndromes in a way that could shed light on the underlying molecular mechanisms and guide classification.

Our objective is to improve phenotypic decision support for rare disorders. Here we describe GestaltMatcher, an innovative approach that uses an image encoder to convert all features of a facial image into a vector of numbers. The encoder can also be thought of as the penultimate layer of a DCNN that was trained on known syndromes, such as DeepGestalt. The vectors resulting from the encoder are then used to build a CFPS for matching a patient’s photo to a gallery of portraits of solved or unsolved cases. The distance between cases in the CFPS quantifies the similarities between the faces, thereby matching patients with known syndromes or identifying similarities between multiple patients with unknown disorders and thereby helping to define new syndromes. Because GestaltMatcher quantifies similarities between faces in this way, it addresses all three of the limitations described above: (1) it can identify “closest matches” among patients with known or unknown disorders, regardless of prevalence; (2) it does not need new architecture or training to incorporate new syndromes; and (3) it creates a search space to explore similarity of facial gestalts based on mutation data, which can point to shared molecular pathways of phenotypically similar disorders.

## Results

The feature encoder of GestaltMatcher computes a Facial Phenotypic Descriptor (FPD) for each portrait image (Figure 2a). Each FPD can be thought of as one coordinate in the CFPS (Figure 2b). The distances between the FPDs in the CFPS form the basis for syndrome classification, delineation of novel phenotypes, and patient clustering. The performance for all three of these use cases depends on the composition of the training set and the gallery. All benchmarking results described in this section, as well as those available through the web service, are based on data from Face2Gene (F2G). The F2G dataset was used to construct a CFPS consisting of 26,065 images from 17,502 subjects who had been diagnosed with a total of 1,115 different syndromes, each supported by at least two cases. We divided the dataset into two categories, the *rare* dataset consisting of 816 ultra-rare and novel syndromes, representing syndromes that we aim to identify, and the *frequent* set, consisting of 299 syndromes already identified by DeepGestalt. The latter set of known syndromes was also used to train the encoder. Each category was further split into a gallery (90% of each syndrome) and a test set (the remaining 10% of each syndrome) (see the Online methods for details).

**Figure 2:**
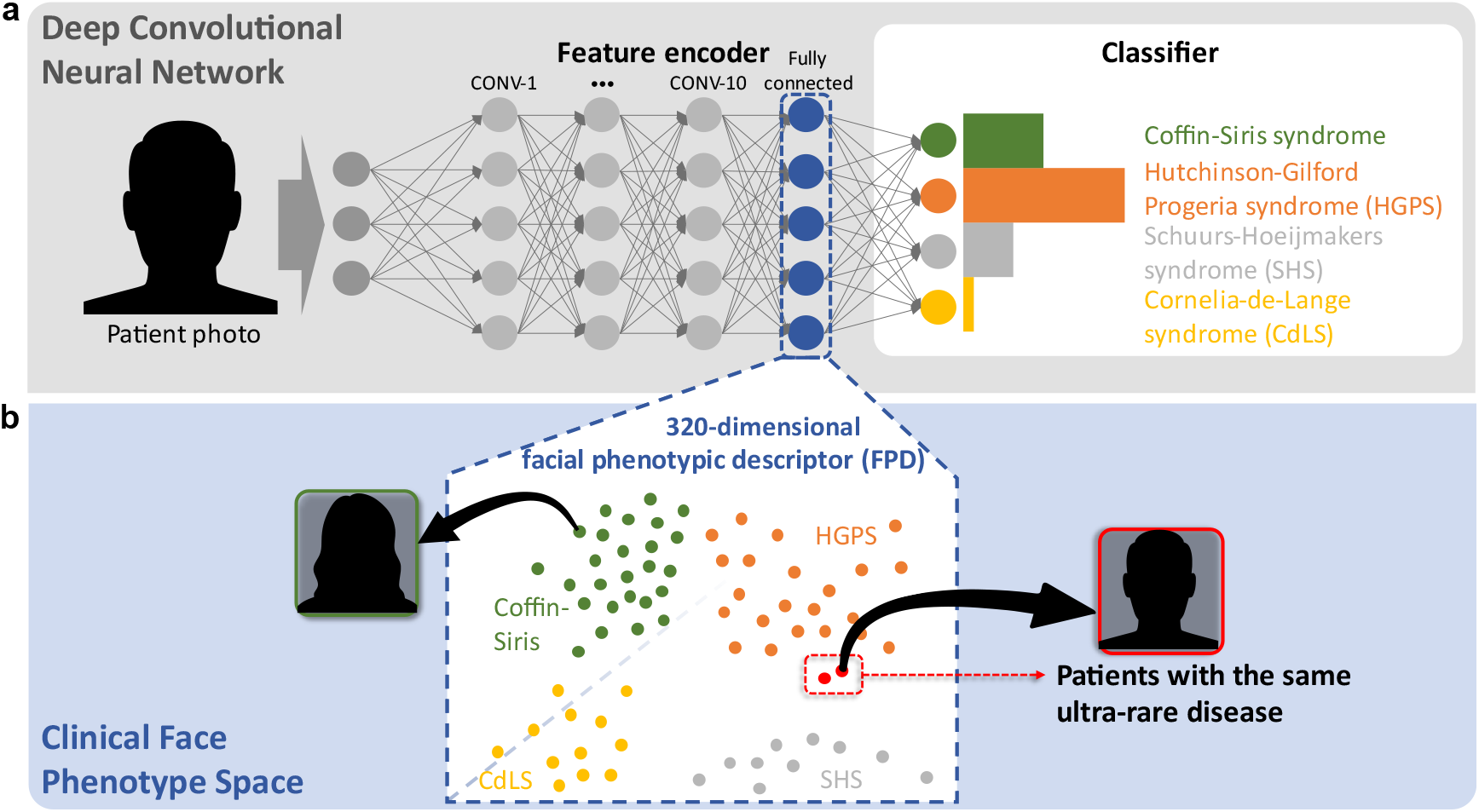
Concept of GestaltMatcher. **a**, Architecture of a deep convolutional neural network (DCNN) consisting of an encoder and a classifier. Facial dysmorphic features of 299 frequent syndromes were used for supervised learning. The last fully connected layer in the feature encoder was taken as a Facial Phenotypic Descriptor (FPD), which forms a point in the Clinical Face Phenotype Space (CFPS). **b**, In the CFPS, the distance between each patient’s FPD can be considered as a measure of similarity of their facial phenotypic features. The distances can be further used for classifying ultra-rare disorders or matching patients with novel phenotypes. Take the input image as an example: the patient’s ultra-rare disease, which is caused by mutations in *LEMD2*, was not in the classifier, but was matched with another patient with the same ultra-rare disorder in the CFPS^4^.

Since F2G data cannot be shared, we compiled the GestaltMatcher database (GMDB), consisting of 4,306 images from 3,693 subjects with 257 different syndromes. This second data set is based on 902 publications, and further cases for which we obtained consent for sharing. All findings described in this section that are based on the F2G data can be reproduced qualitatively on the GMDB data and are listed in the Supplemental Material.

### Training on images of dysmorphism improves the performance of the FPD

To investigate the importance of using a syndromic features encoder rather than a normal facial features encoder, we compared FPDs that are based on the same architecture, but trained on different data. The first encoder, which we refer to as *Enc-healthy*, was only trained on data from healthy individuals in CASIA-WebFace^19^. The second encoder, which we refer to as *Enc-F2G* (for Face2Gene), was first trained on the faces of healthy subjects and then fine-tuned by training on dysmorphic faces from the gallery of patients with frequent syndromes. All images were encoded separately for each encoder. We then evaluated the performance of the encoders on test sets of syndromes from the frequent set and from the rare set. The performance metric was the percentage of test cases (with known diagnosis) for which an FPD with the matching disorder was within the *k* closest diagnoses in the CFPS (the top-*k* accuracy). The features created by Enc-F2G performed better in the matching process than those created with Enc-healthy (Table 1). This emphasizes the importance of training the encoder on data from faces with dysmorphic phenotypes and not only on healthy faces. The features created by Enc-F2G improved the accuracy of matching within the top-10 closest images from 31.46% to 49.12% for the frequent category. Furthermore, the top-10 accuracy improved from 21.77% to 29.56% for the rare syndromes, which do not overlap with the frequent syndromes. The larger relative improvement of 56% on the frequent test set versus 36% for the rare set could possibly be explained as Enc-F2G being better suited to encode syndromes of the frequent set because it was previously trained on these disorders. Likewise, for some of the 816 novel disorders, the characteristic features were not yet optimally represented by Enc-F2G because features of these disorders were not part of the training set.

**Table 1:** Performance comparison between classification and clustering with different encoders on sets of known disorders. The DCNNs of Enc-F2G (softmax), Enc-F2G, and Enc-healthy have the same architecture. Enc-F2G (softmax) and Enc-F2G training were initiated with CASIA-WebFace and further fine-tuned on photos of patients in the Face2Gene frequent set. The Enc-F2G (softmax) model is the same as Enc-F2G, but using the softmax values of the layer instead of cosine distances between the FPDs in the CFPS. For the top-1 to top-30 columns, the best performance in each set is boldfaced. The numbers of images and syndromes in the rare set are averaged over ten splits. Enc-F2G outperformed Enc-healthy on both types of syndromes, showing the importance of fine-tuning on patient photos for learning facial dysmorphic features. The top-10 accuracy of Enc-F2G only drops by 2.27 percentage points after increasing the number of cases in the gallery and almost quadrupling the number of supported syndromes from 299 to 1,115. ^a^ Number of images in frequent gallery and rare gallery. ^b^ Average of ten splits in the frequent gallery and rare gallery. ^c^ Number of syndromes in the frequent gallery and rare gallery.

| Test set | Model | Images |  | Supported syndromes | Null top-1 accuracy | Top-1 | Top-5 | Top-10 | Top-30 |
| --- | --- | --- | --- | --- | --- | --- | --- | --- | --- |
|  |  | Gallery | Test |  |  |  |  |  |  |
| F2G-frequent | Enc-F2G (softmax) | - | 2,669 | 299 | 0.33% | **35.94%** | **52.45%** | **63.91%** | **78.13%** |
| F2G-frequent | Enc-F2G | 19,950 | 2,669 | 299 | 0.33% | 21.06% | 39.62% | 49.12% | 67.98% |
| F2G-frequent | Enc-healthy | 19,950 | 2,669 | 299 | 0.33% | 10.69% | 23.69% | 31.46% | 50.80% |
| F2G-rare | Enc-F2G | 2,348.8 | 1,183.3 | 816 | 0.12% | **13.66%** | **23.62%** | **29.56%** | **40.94%** |
| F2G-rare | Enc-healthy | 2,348.8 | 1,183.3 | 816 | 0.12% | 9.46% | 16.87% | 21.77% | 31.77% |
| F2G-frequent | Enc-F2G | 22,298<sup>a</sup> | 2,669 | 1,115<sup>c</sup> | 0.09% | **20.15%** | **37.81%** | **46.85%** | **64.21%** |
| F2G-frequent | Enc-healthy | 22,298<sup>a</sup> | 2,669 | 1,115<sup>c</sup> | 0.09% | 9.70% | 22.51% | 29.80% | 48.24% |
| F2G-rare | Enc-F2G | 22,298.8<sup>b</sup> | 1,183.3 | 1,115<sup>c</sup> | 0.09% | **7.07%** | **14.19%** | **17.67%** | **24.41%** |
| F2G-rare | Enc-healthy | 22,298.8<sup>b</sup> | 1,183.3 | 1,115<sup>c</sup> | 0.09% | 4.02% | 8.84% | 11.73% | 16.61% |

The same trend of improvement by fine-tuning on a diverse but smaller set of syndromic photos is also seen on the public GMDB dataset (Enc-GMDB vs Enc-F2G in Supplementary Table 1). These results suggest that an encoder that is fine-tuned on as many syndromic faces as possible, such as DeepGestalt, is a better fit for the task of syndrome classification than one trained only on healthy faces. Moreover, DeepGestalt’s FPD provides a better generalization or clustering than the FPD encoded by CASIA for rare syndromes that it had not previously seen.

### Syndromic diversity improves the performance on novel disorders

Earlier definitions of the FPD were mainly based on training a network with a small selection of common and highly characteristic syndromes^5,9^. In principle, we could train GestaltMatcher’s encoder on all 1,115 different syndromes in our dataset. However, most of the facial phenotypes that have recently been linked to a gene are either ultra-rare or less distinctive, and using a very unbalanced training set with many ultra-rare disorders linked to only few cases may add noise without substantial additional benefit. We therefore analyzed the influence of the number of syndromes on the encoder’s fine-tuning by incrementally increasing their number starting with the most frequent ones. Due to the imbalance among the disorders added each time, the improvement could be affected by the additional number of training subjects. Therefore, we used the same number of subjects for each syndrome. In this section, the test set consists only of disorders from the rare set that the encoder has not seen. The training procedure and averaging of the readout is described in detail in the Online methods.

When we increase the number of training syndromes, the accuracy increases (Figure 3). In general, the performance is also higher when more individuals per syndrome are used for training. Particularly when more than 50 syndromes are used, the curve for training with 20 subjects/syndrome is above the curve for 10 subjects/syndrome, and so on. The same trend is also shown in the public GMDB dataset (Supplementary Figures 2 and 3).

**Figure 3:**
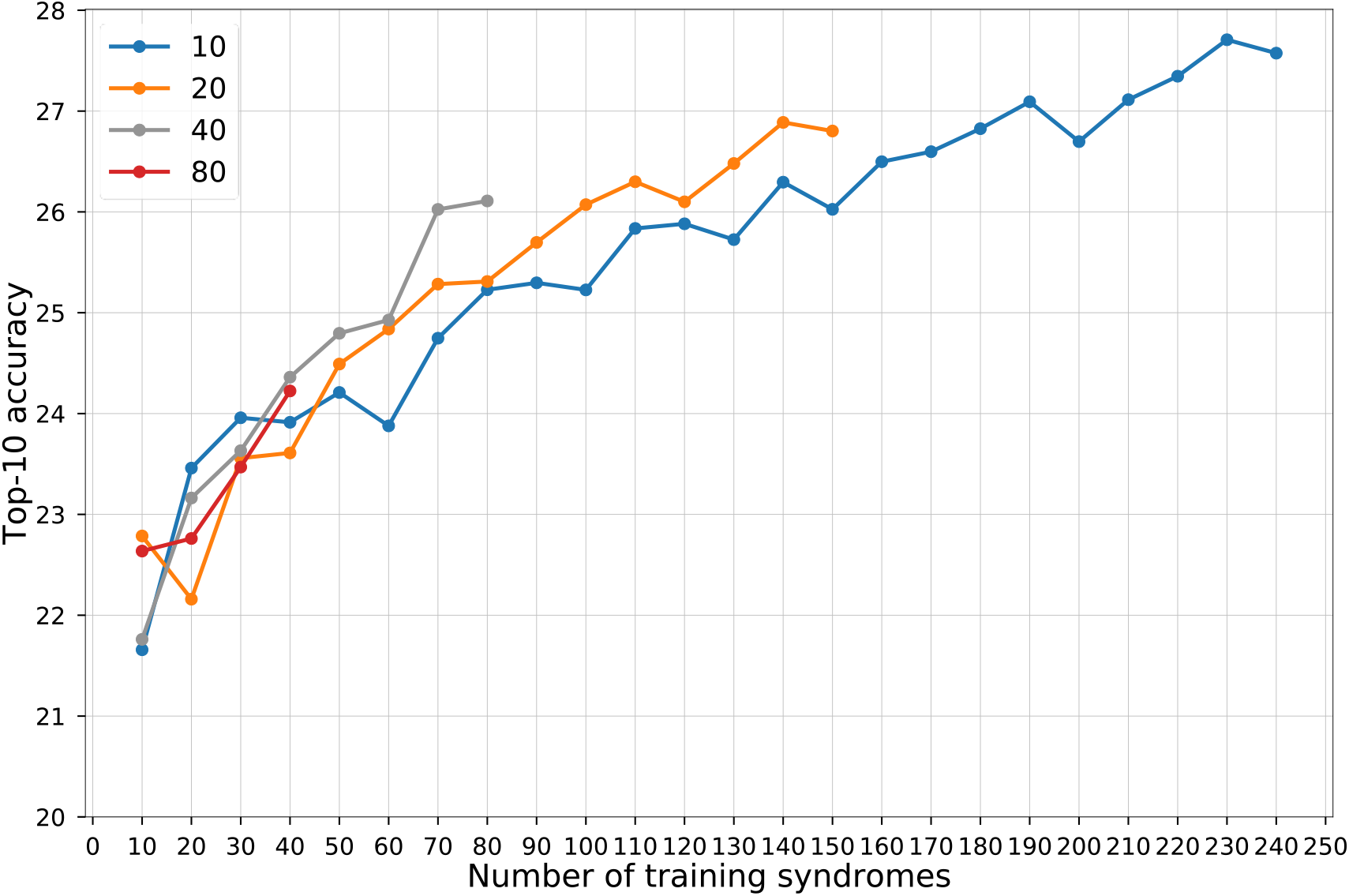
Influence of the number of syndromes included in model training. The x-axis is the number of syndromes used in model training. The y-axis shows the average top-10 accuracy of testing on the rare set. Each line uses the same number of subjects per syndrome, which is shown in the key. For each point, we train the models five times with five different splits, and average the results. The null accuracy (the expected value if the encoder returned random predictions) is 1.2% (10/816).

Moreover, double the number of syndromes is better than double the number of subjects in most of the combinations (Supplementary Figure 4). The effect of doubling the number of syndromes used for training is greater when the base sample size is larger than 1200 subjects (Supplementary Figures 5 and 6). Therefore, both of the findings suggest that increasing the syndromic diversity in the training set improves the performance on novel disorders.

### Top-10 accuracy plateaus when encoders are fine-tuned on more than 150 syndromes

In the previous section, we analyzed the impact of syndromic diversity in a balanced setting, that is, the dynamics of increasing the number of syndromes while keeping the size of the increments (the number of added subjects) equal. In this section we analyze the influence of the number of syndromes on model training in the real-world scenario; that is, when using all of the subjects per syndrome (Supplementary Figure 7). The top-10 accuracy improved considerably until about 150 syndromes, representing roughly 90% of the subjects in the entire training set. Almost doubling the number of syndromes to 299 with the remaining 10% of subjects only increases the performance marginally. From these dynamics, we can conclude that including additional syndromes beyond 299 for defining the FPD will provide little benefit, and we decided to proceed with the Enc-F2G encoder in the following section that is based on the 299 syndromes described in the original DeepGestalt paper.

### Performance comparison between GestaltMatcher and DeepGestalt

To validate the GestaltMatcher approach, we first worked with the 323 images of patients with 91 syndromes from the London Medical Database (LMD)^20^ that were already used for benchmarking the performance of DeepGestalt^12^. When using the frequent gallery, which contains syndromes that DeepGestalt currently supports, GestaltMatcher achieved 64.30% and 86.59% accuracy within the top-10 and top-30 ranks, respectively, which was lower than the 81.28% top-10 accuracy and 88.34% top-30 accuracy achieved by DeepGestalt with a Enc-F2G softmax approach (Supplementary Table 2 and 3). However, when we used the gallery of all 1,115 syndromes for GestaltMatcher (frequent + rare) which is a search space that is roughly four times larger, the top-10 and top-30 dropped by only 2.40 percentage points and 5.17 percentage points, respectively (Supplementary Table 2). Moreover, we performed the same evaluation on the F2G-frequent test set and GMDB-frequent test set. When the number of syndromes in the gallery was increased from 299 to 1,115, the top-10 and top-30 also dropped slightly by 2.27 and 3.77 percentage points for the F2G-frequent test set (Table 1). The results of the GMDB frequent test also dropped slightly while supporting more than twice the number of syndromes (Supplementary Table 1). These results indicate that the GestaltMatcher clustering approach is highly scalable and robust to adding new disorders, without the limitations of a classification approach.

### Matching undiagnosed patients from unrelated families

In the second use case, we envision GestaltMatcher as a phenotypic complement to GeneMatcher^21^. To prove that we can match patients from unrelated families who have the same disease by using only their facial photos, we selected syndromes from 15 recent GeneMatcher publications with titles containing the phrase “facial dysmorphism”. In contrast to the benchmarking of the previous section, the gallery now consists of subjects with rare syndromes to simulate undiagnosed subjects and as a consequence, ranks refer to individuals and not disorders. For the evaluation we still have to reveal in the end whether an individual from the gallery is a match for a test case or not. This implies that non-matching cases can harm the performance more than in the previous section. For instance, if the first matching individual is at rank 30, but the 29 non-matching individuals with higher similarity to the test case all together have only four non-matching disorders, then this match would contribute to the top-5 accuracy in the previous section that matched on disorders but to the top-30 accuracy in this section that matches to individuals. Only the top-1 accuracy remains the same in both benchmarks.

In this scenario, we matched 30 of 91 subjects and connected 26 of 79 families when using the top-10 criterion (Table 2 and Supplementary Figure 8). When using the top-30 rank, 48 of 91 subjects were matched, and 40 of 79 families were connected. Enc-healthy, which is trained only with healthy subjects, matched only 40 out of 91 subjects and connected 34 out of 79 families using the top-30 rank (Supplementary Table 4). Hence, using the encoder trained with facial dysmorphic subjects improves the matching considerably.

**Table 2:** Matching of novel phenotypes on a GeneMatcher validation set. In the discovery mode for novel phenotypes, all cases in the gallery are without diagnosis. For the performance readout, only the correct disease gene of a match is revealed. For individuals of the *TMEM94* study, e.g. eight out of ten subjects had an image from another family within the top-10 rank, and five of the six families had at least one subject from another family in their top-10 rank. For top-30 all subjects and families matched. This table is based on the ranks from the similarity matrices in Figure 4 and Supplementary Figure 8. The accuracy of connected subjects corresponds to the accuracy of using Enc-F2G on the F2G-rare test set in the Table 1 in discovery mode in the gallery of almost the same size.

| Gene | PMID | Total families<br>(Subjects) | Connected families (subjects)<sup>a</sup> |  |
| --- | --- | --- | --- | --- |
|  |  |  | Top-10 | Top-30 |
| *BPTF*<sup>32</sup> | 28942966 | 6 (6) | 0 (0) | 2 (2) |
| *CCDC47*<sup>33</sup> | 30401460 | 4 (4) | 0 (0) | 0 (0) |
| *CHAMP1*<sup>34</sup> | 27148580 | 4 (4) | 2 (2) | 4 (4) |
| *CHD4*<sup>35</sup> | 27616479 | 3 (3) | 0 (0) | 0 (0) |
| *DDX6*<sup>36</sup> | 31422817 | 4 (4) | 4 (4) | 4 (4) |
| *EBF3*<sup>37</sup> | 28017373 | 6 (7) | 0 (0) | 0 (0) |
| *FBXO11*<sup>38</sup> | 30679813 | 17 (17) | 5 (5) | 9 (9) |
| *HNRNPK*<sup>39</sup> | 26173930 | 3 (3) | 3 (3) | 3 (3) |
| *KDM3B*<sup>40</sup> | 30929739 | 9 (9) | 0 (0) | 2 (3) |
| *LEMD2*<sup>4</sup> | 30905398 | 2 (2) | 2 (2) | 2 (2) |
| *OTUD6B*<sup>41</sup> | 28343629 | 4 (9) | 3 (4) | 3 (6) |
| *PACS2*<sup>42</sup> | 29656858 | 6 (6) | 0 (0) | 2 (2) |
| *TMEM94*<sup>22</sup> | 30526868 | 6 (10) | 5 (8) | 6 (10) |
| *WDR37*<sup>43</sup> | 31327508 | 4 (4) | 2 (2) | 3 (3) |
| *ZNF148*<sup>44</sup> | 27964749 | 3 (3) | 0 (0) | 0 (0) |
| Total | - | 79 (91) | 26 (30) | 40 (48) |
| Average | - | - | 32.91% (32.97%) | 50.63% (52.75%) |

<sup>a</sup> Number of families (subjects) matched by a photo from another family in the top-10 or top-30 rank.

As an example, in a study of *TMEM94*^22^, eight of the ten photos in six different families were matched, and five of six families were connected within the top-10 rank. When the three test images in family 2 (F-2-5, F-2-7, F-2-9) were tested, the other five families were among those in the top-30 rank (Figure 4). The youngest brother, F-2-5, matched families 1, 3, 5, and 6, and one sister, F-2-7, matched families 1, 4, and 6. Another sister, F-2-9, matched families 1, 4, 5, and 6. The six families were recruited at five different institutes in India, Qatar, the United States (NIH Undiagnosed Diseases Network), and Switzerland, indicating that GestaltMatcher can also connect patients of different ethnic origins. However, a more systematic analysis of pairwise distances still revealed considerably smaller distances between subjects with *de novo* mutations and their family members than between these subjects and unrelated individuals (Supplementary Figure 9). This reflects similarities in the nonclinical features of the face, which is also higher within the same ethnicity and is a known confounding factor for the GestaltMatcher approach. However, it is a bias that can be attenuated^23^ and will also diminish over time when more diverse training data become available^24^.

**Figure 4:**
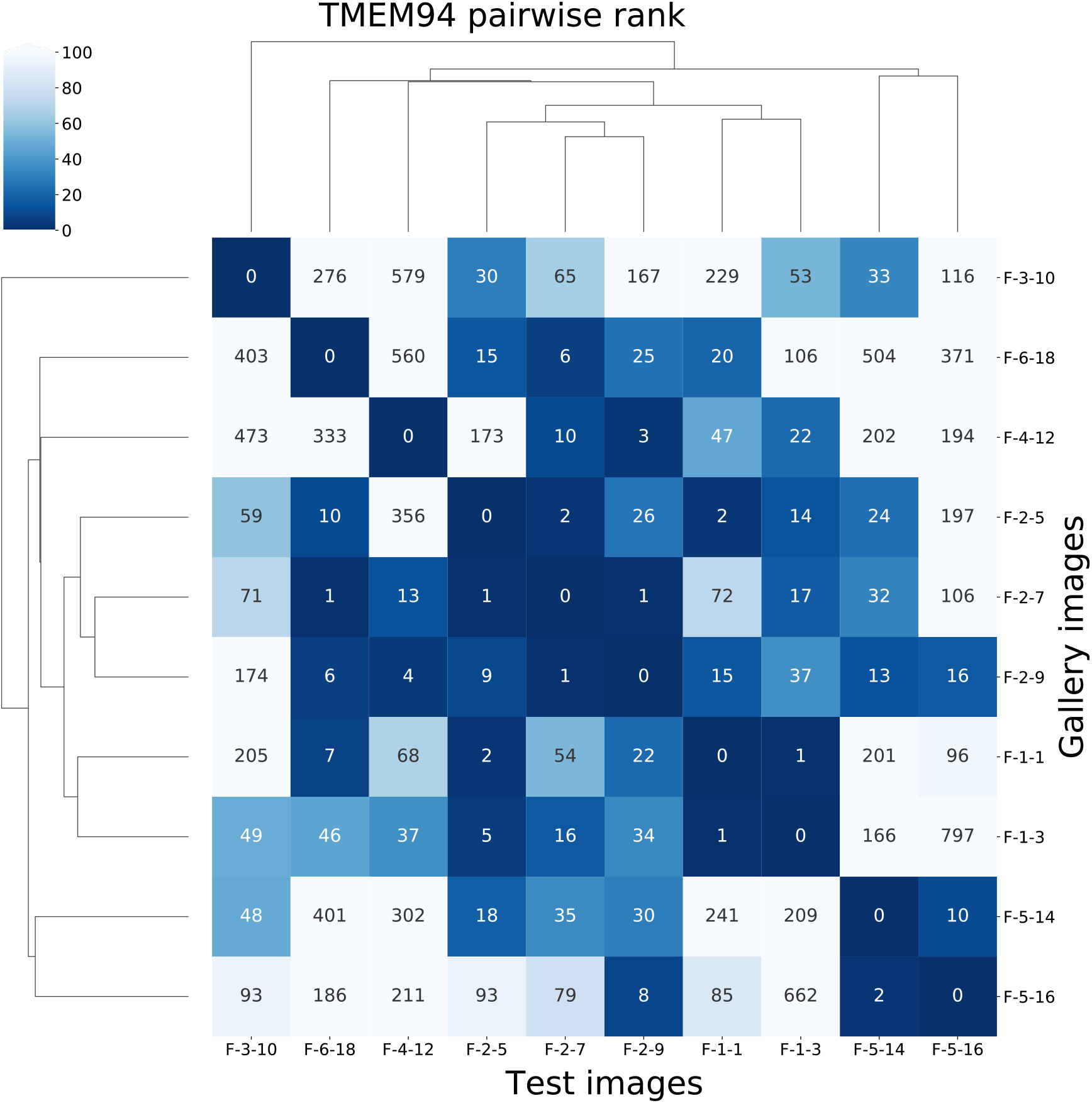
Pairwise ranks of subjects with mutations in *TMEM94*. Each label consists of family numbering and subject numbering, which are the same as in the original publication^22^. For example, F-2-7 means the seventh subject in the second family. Each column is the result of testing the image indicated at the bottom of the column. The number in the box is the rank to the corresponding image in the gallery. The fourth column starting from the left is the result of testing F-2-5, and the fourth row from the bottom shows that F-1-1 has a rank of 2 for F-2-5. In the fifth to seventh rows from the bottom are the ranks from family 2, which is the same family that F-2-5 is from.

### GestaltMatcher and human experts agree on syndrome distinctiveness

We hypothesized that some of the ultra-rare disorders that were linked to their disease-causing genes early on, such as Schuurs-Hoeijmakers syndrome in 2012,^25^ have particularly distinctive facial phenotypes. To systematically analyze the dependence of disease-gene discovery on the distinctiveness of a facial gestalt, we asked three expert dysmorphologists (S.M., N.E., and K.W.G.) to grade 299 syndromes on a scale from 1 to 3. The more easily they could distinguish the diseases, and the more characteristic of the disease they deemed the facial features, the higher the score. All three syndromlogists agreed on the same score for 195/299 syndromes, yielding a concordance of 65.2%. We then selected 50 syndromes as a test set and trained the model with the remaining 249 syndromes. We analyzed the correlation of the mean of the distinctiveness score from human experts with the top-10 accuracy that GestaltMatcher achieves for these syndromes without having been trained on them (Figure 5a, Supplementary Table 6). The Spearman’s rank correlation coefficient was 0.400 (*P* = 0.004), indicating a clear positive correlation between distinctiveness score and top-10 accuracy. Syndromes with a higher average score tended to perform better, with Schuurs-Hoeijmakers syndrome being amongst the best-performing syndromes in GestaltMatcher. The analysis on 20 selected syndromes from the GMDB dataset also showed a positive correlation between distinctiveness score and top-5 accuracy (Supplementary Figure 10 and Supplementary Table 7).

**Figure 5:**
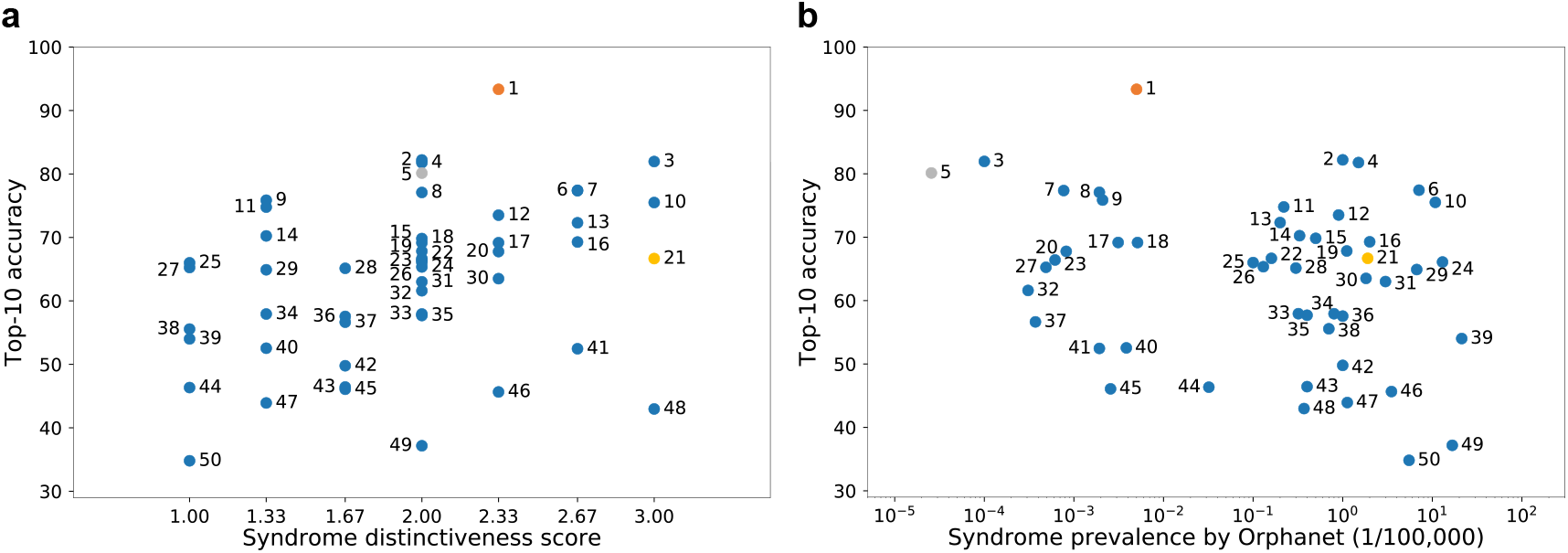
Correlation among syndrome prevalence, distinctiveness score, and top-10 accuracy. **a**, Distribution of top-10 accuracy and distinctiveness score. The Spearman rank correlation coefficient was 0.400 (*P* = 0.004). **b**, Distribution of top-10 accuracy and prevalence. The Spearman rank correlation coefficient was –0.217 (*P* = 0.130) The details of each syndrome can be found in Supplementary Table 6 using the syndrome ID shown in the figure; syndrome 5 is Schuurs-Hoeijmakers syndrome. The y-axis shows the average top-10 accuracy of the experiments over 100 iterations.

The correlation for GestaltMatcher accuracy and disease prevalence was not significant (*P* = 0.130; Figure 5b). This also means that ultra-rare disorders share a similar distribution of distinctiveness with more common ones, which is important for estimates about the performance of GestaltMatcher on novel phenotypes in the real world.

### Characterization of phenotypes in the CFPS

When syndromologists cannot find a molecular cause for a patient’s phenotype in diagnostic-grade genes after extensive work up in the lab, it becomes a research case and they may compare the patient’s condition to known disorders. For example a potentially novel phenotype could be described as “syndrome *XY*–like” to build a case group for further molecular analysis through genome sequencing. In GestaltMatcher, this is the third use case, and such comparisons can be supported by cluster analysis in the CFPS with the cosine distance as a similarity metric (Supplementary Table 8).

If a novel disease gene has been identified and the similarities of the patients to known phenotypes outweigh the differences, OMIM groups them into a phenotypic series. On the gene or protein level, such phenotypic series often correspond to molecular-pathway diseases, such as GPI-anchor deficiencies for hyperphosphatasia with mental retardation syndrome (HPMRS) or cohesinopathies for CdLS. For our cluster analysis, we sampled subjects in our database with subtypes of four large phenotypic series and found high intersyndrome separability in addition to considerable intrasyndrome substructure in Noonan syndrome, CdLS, Kabuki syndrome, and mucopolysaccharidosis. A *t*-SNE^26^ projection of the FPDs into two dimensions yielded the best visualization results (Supplementary Figure 11). Although any projection into a smaller dimensionality might cause a loss of information, the clusters are still clearly visible for the 743 subjects sampled from these four phenotypic series. This observation provides further evidence that characteristic phenotypic features are encoded in the FPDs.

To demonstrate the separability of syndromes with facial dysmorphism, we also used *t*-SNE to project 4,353 images of the ten syndromes from the frequent set with the largest number of subjects and 872 images of ten non-distinct syndromes (syndromes without facial dysmorphism) into 2D space. In addition, we calculated the Silhouette index^27^ for both of these datasets. The FPDs of the frequent syndromes showed ten clear clusters of subjects (Supplementary Figure 12), but the *t*-SNE projection of subjects with non-distinct syndromes created no clear clusters. Moreover, the Silhouette index of the frequent syndromes (0.11) was higher than that of the non-distinct syndromes (–0.005); the negative Silhouette index indicates poor separation of the non-distinct syndromes.

### GestaltMatcher as a tool for clinician scientists

The transition of a research case to a diagnostic case is best described by the process of matching unrelated patients in the CFPS who share a molecular abnormality until statistical significance is reached. We illustrate this process for the novel disease gene *PSMC3* in a demonstration on the GestaltMatcher web service (Supplementary Figure 13, www.gestaltmatcher.org). Ebstein *et al.* (not yet published) report 18 patients with a neurodevelopmental disorder of heterogeneous dysmorphism that is caused by *de novo* missense mutations in *PSMC3*, which encodes a proteasome 26S subunit. Although not all *PSMC3* patients have the same facial phenotype, the proximity of two unrelated patients in the CFPS who share the same *de novo PSMC3* mutation is exceptional. Their distance is comparable to the pairwise distances of patients with the recurring missense mutation R203W in *PACS1*, which is the only known cause of Schuurs-Hoeijmakers syndrome. On the one hand, the high distinctiveness of these two *PSMC3* cases with the same mutation allows direct matching by phenotype. On the other hand, the pairwise similarities of 10 out of 18 patients in the CFPS for which portraits were available also hints that the protein domains have more than one function. The previously described scalability of GestaltMatcher makes an exploration of such similarities in the CFPS possible for any number of cases as soon as they have been added to the gallery of undiagnosed patients.

## Discussion

GestaltMatcher’s ability to match previously unseen syndromes, that is, those for which no patient is included in the training set, distinguishes it from other approaches. Since matching of unseen syndromes is not only of importance for ultra-rare disorders but can be considered for the discovery of novel diseases, GestaltMatcher could also speed up the process of delineating new disorders.

Importantly, GestaltMatcher provides the flexibility to easily scale up the number of supported syndromes or the number of unsolved cases without substantial loss in performance. The LMD validation analysis revealed that the use of the softmax approach, that is classification based on the values of the last layer representing disorders, outperformed GestaltMatcher. However, the GestaltMatcher encoder, that is clustering in the CFPS with values of the penultimate layer representing features, demonstrated high scalability by yielding similar performance when the number of supported syndromes was increased from 299 to 1,115. Furthermore, the distinctiveness of a syndrome correlated with the performance (Figure 5a), whereas syndrome prevalence did not (Figure 5b). Thus, GestaltMatcher can match a syndrome with a distinguishable facial gestalt even if it is of extremely low prevalence. This enables us to avoid the long development flow currently required to support and discover novel syndromes (Supplementary Figure 1). Instead, matching can be offered instantly for all unsolved cases with available frontal images for which consent has been provided for inclusion in the tool. If the gallery is populated by cases with a disease-causing mutation in a diagnostic-grade gene, we consider this a diagnostic work-up. In contrast, if the gallery is populated by further undiagnosed cases, it is a use case comparable to GeneMatcher.

GestaltMatcher’s framework also allows us to abstract the encoding of a dataset away from the classification task. For example, one can evaluate both phenotypic series and pleiotropic genes within a single CFPS, or obtain the most-similar patients for each of the matched syndromes, with minor computational cost (i.e., in real time). Furthermore, the GestaltMatcher framework computes the similarity between each of the test set images across the entire dataset of images. This similarity can be computed using different metrics, e.g., cosine or Euclidean distance. The results are then aggregated according to the chosen configuration. For example, image similarity can be aggregated at the patient level or the syndrome level. Furthermore, the dataset can be filtered according to different parameters (such as ethnicity, disease-causing genes, or age) to further customize the evaluation.

One of the key features of GestaltMatcher is the ability to match patients and quantify their syndromic similarity. For clinician scientists who often face two different tasks in their daily practice, this means: (1) assessing whether the patient’s phenotype is specific for a known disorder. If e.g. a variant of unclear clinical significance is found in a diagnostic grade gene, this would be considered as supporting evidence for the pathogenicity^28,29^. (2) assessing whether the phenotypic similarity of an unsolved case to other individuals without a diagnosis is high enough to form e.g. a case group that is further analyzed. This could e.g. result in the identification of potentially deleterious variants in a novel disease gene and would represent the phenotypic complement to existing matching approaches on the molecular level. Several online platforms, such as GeneMatcher, MyGene2 (https://mygene2.org/MyGene2), and Matchmaker Exchange^30^, already allow physicians to look for similar patients based on sequencing information, and over the past few years these platforms have enabled the matching of thousands of patients. However, although phenotypic data, encoded e.g. in HPO terms, are usually exchanged after contact has been established, automated facial matching technology has not yet been included in any of these platforms.

Since its first proof of concept, in which GestaltMatcher was used to identify two unrelated patients from different countries with the same novel disease, caused by the same *de novo* mutation in *LEMD2*^4^, our approach has successfully been applied to further ultra-rare disorders (Figure 1). We matched 40 of 79 different families in 15 GeneMatcher publications by top-30 rank (Table 2 and Supplementary Figure 8), and 11 candidate genes are currently under evaluation. This result shows the power and potential of GestaltMatcher to identify novel syndromes. Although the number of individuals and the diversity of their phenotypes will affect the performance, cases with a high syndromic similarity will remain matchable due to the high dimensionality of the CFPS.

We therefore hope that GestaltMatcher will be readily integrated into other matching platforms to aid in determining which phenotypes should be grouped together into a syndrome or phenotypic series, as well as linking individual patients to a molecular diagnosis.

## Code availability

GestaltMatcher is a partially proprietary framework. Although the source code for cropping the face cannot be shared, the architecture of the CNN, as well as a web service of the trained version of the tool is accessible for use by health care professionals free of charge at www.gestaltmatcher.org.

## Data availability

The data that support the findings of this study are divided into two groups, sharable data (GMDB) and non-sharable data (F2G). GMDB is accessible via www.gestaltmatcher.org. Restricted data are curated from Face2Gene users under a license and cannot be published in order to protect patient privacy.

## Online methods

### Study approval

This study is governed by the following Institutional Review Board (IRB) approval: Charité–Universitätsmedizin Berlin, Germany (EA2/190/16); UKB Universitätsklinikum Bonn, Germany (Lfd.Nr.386/17). The authors have obtained written informed consent given by the patients or their guardians, including permission to publish photographs.

### Face2Gene datasets

We collected images of subjects with clinically or molecularly confirmed diagnoses from the Face2Gene database (https://www.face2gene.com). Extracted, deidentified data were used to remove poor-quality or duplicated images from the dataset without viewing the photos. After removing images of insufficient quality, the dataset consisted of 26,152 images from 17,560 subjects with a total of 1,115 syndromes (Supplementary Table 9).

GestaltMatcher was designed to distinguish syndromes with different properties. We separated syndromes by the number of affected subjects and whether they had already been learned by the DeepGestalt model. Supplementary Figure 14 provides an overview of how the dataset was divided. The current DeepGestalt approach requires at least seven subjects to learn a novel syndrome. We first used this threshold to separate the syndromes into “frequent” and “rare” syndromes. The objective of our study was to improve phenotypic decision support for “rare disorders”. However, frequent syndromes that are not associated with facial dysmorphic features cannot be modeled by DeepGestalt. We therefore further selected 299 frequent syndromes that possess characteristic facial dysmorphism recognized by DeepGestalt as “frequent syndromes”. The frequent syndromes were used to validate syndrome prediction and the separability of subtypes of a phenotypic series because these syndromes are known to have facial dysmorphic features that are well recognized by the DeepGestalt encoder. For rare syndromes, we sought to demonstrate that GestaltMatcher could predict a syndrome even if facial images were publicly available for only a few subjects. It is noteworthy that, for more than half of all known disease-causing genes, fewer than ten cases with pathogenic variants have been submitted to ClinVar (Figure 1). Of the 1,115 syndromes in the entire dataset, 299 were frequent and 816 were rare. DeepGestalt cannot yet be applied to rare syndromes.

We further divided each of these two datasets into a gallery and a test set. The gallery is the set of subjects that we intend to match, given a subject from the test set. First, 90% of subjects with each frequent syndrome were used to train the models, and the remaining 10% of subjects were used to validate the DeepGestalt training; the 90% then became the frequent gallery and the 10% were assigned to the frequent test set. For the rare dataset, we performed 10-fold cross-validation. In each syndrome, 90% and 10% of subjects were assigned to the gallery and test set, respectively. The test sets were designed to have the same distribution of distinctiveness as the training sets.

Matching only within a dataset would not represent a real-world scenario. Therefore, the galleries of the two datasets were later combined into a unified gallery that was used to search for matched patients.

Please note that the threshold of seven subjects to divide the dataset into frequent and rare is to compare GestaltMatcher to DeepGestalt, which both use the same training data. We could adjust this threshold higher or even remove this threshold in the future.

### GMDB dataset

We collected images of subjects with clinically or molecularly confirmed diagnoses from publications and individuals that gave appropriate informed consent for the purpose of this study. This dataset can be used as a public training and test set for benchmarking and is available at GestaltMatcher Database (https://gestaltmatcher.gene-talk.de).

At the time of the data freeze on 9 June 2021, the dataset consisted of 4,306 images of 3,693 subjects with a total of 257 syndromes from 902 publications (Supplementary Table 9). Six of the 3,693 subjects have not yet been published, but appropriate consent has been obtained. For a fair comparison with the Face2Gene dataset, we performed the data separation in the same way. The dataset was first split by the same threshold (seven subjects) into frequent and rare datasets, giving 139 syndromes in the frequent dataset and 118 syndromes in the rare set. Both datasets were also later separated into gallery and test sets. The data split is shown in Supplementary Figure 15. Of the 3,693 subjects in GMDB, 963 are also in Face2Gene dataset. To use the GMDB rare set as the test set for both the GMDB frequent set and the Face2Gene frequent set, we made sure that there is no syndrome that is in both the GMDB rare set and Face2Gene frequent set (Supplementary Figure 16).

### DeepGestalt encoder

The preprocessing pipeline of DeepGestalt includes point detection, facial alignment (frontalization), and facial region cropping. During inference, a facial region crop is forward passed through a deep convolutional network (DCNN) and ultimately gives the final prediction of the input face image. The DeepGestalt network consists of ten convolutional layers (Conv) with batch normalization (BN) and a rectified linear activation unit (ReLU) to embed the input features. After every Conv-BN-ReLU layer, a max pooling layer is applied to decrease spatial size while increasing the semantic representation. The classifier part of the network consists of a fully connected linear layer with dropout (0.5). In this study, we considered the DeepGestalt architecture as an encoder–classification composition, pipelined during inference. We chose the last fully connected layer before the softmax classification as the facial feature representation (facial phenotypic descriptor, FPD), resulting in a vector of size 320.

DeepGestalt was first trained on images of healthy individuals from CASIA-WebFace^19^, and later fine-tuned on a dataset with patient images (Face2Gene or GMDB). The encoder without fine-tuning on patient images was called Enc-healthy. The encoder later trained on 299 frequent syndromes in the Face2Gene dataset was named Enc-F2G. The encoder trained on 139 frequent syndromes in GMDB was named Enc-GMDB. In the following sections, we have several encoders trained on different subsets of the Face2Gene and GMDB datasets. The summary of all the encoders used in this study is shown in Supplementary Table 5. To compare GestaltMatcher and DeepGestalt, we used a model using softmax for predicting syndromes, which we called “Enc-F2G (softmax)”. This model is the same as Enc-F2G; the only difference is that Enc-F2G (softmax) used softmax in the last layer for prediction, as in DeepGestalt, and Enc-F2G used the cosine distance of FPDs for prediction.

Our first hypothesis was that images of patients with the same molecularly diagnosed syndromes or within the same phenotypic series, and who also share similar facial phenotypes, can be encoded into similar feature vectors under some set of metrics. Moreover, we hypothesized that DeepGestalt’s specific design choice of using a predefined, offline-trained, linear classifier could be replaced by other classification “heads”, for example, *k*-Nearest Neighbors using cosine distance, which we used for GestaltMatcher.

### Descriptor projection: Clinical Face Phenotype Space

Each image was encoded by the DeepGestalt encoder, resulting in a 320-dimensional FPD. These FPDs were further used to form a 320-dimensional space called the Clinical Face Phenotype Space (CFPS), with each FPD a point located in the CFPS, as shown in Figure 2. The similarity between two images is quantified by the cosine distance between them in the CFPS. The smaller the distance, the greater the similarity between the two images. Therefore, clusters of subjects in the CFPS can represent patients with the same syndrome, similarities among different disorders, or the substructure under a phenotypic series.

### Evaluation

To evaluate GestaltMatcher, we took the images in the test set as input and positioned them in the CFPS defined by the images of the gallery. We calculated the cosine distance between each of the test set images (for which the diagnoses were known in this proof-of-concept study) and all of the gallery images. Then, for each test image, if an image from another subject with the same disorder in the gallery was among the top-*k* nearest neighbors, we called it a top-*k* match. We then benchmarked the performance by averaging the top-*k* accuracy (percent of test images with correct matches within the top *k*) of each syndrome to avoid biasing predictions toward the major class. We further compared the accuracy of each syndrome in the frequent and rare syndrome subsets to investigate whether GestaltMatcher can extend DeepGestalt to support more syndromes. To compare its performance on predicting syndromes with DeepGestalt, we first performed image aggregation on the syndrome level before calculating top-*k* accuracy, which means that only the nearest image of each syndrome will be taken into account.

### London Medical Dataset validation analysis

We compiled 323 images of patients diagnosed with 91 frequent syndromes from the LMD^19^ and used this as the validation set for frequent syndromes. We first evaluated the validation set using softmax, which is a DeepGestalt method. To compare the performance with that of GestaltMatcher, we evaluated the performance of GestaltMatcher on two different galleries: a gallery of frequent syndromes consisting of 19,950 images of patients with 299 syndromes, and a unified gallery consisting of 22,298 images of patients with 1,115 syndromes. We then reported the top-*k* accuracy and compared the results of these three settings (DeepGestalt with softmax, GestaltMatcher with frequent gallery, and GestaltMatcher with unified gallery).

### Rare syndromes analysis

To understand the potential for matching rare syndromes, we trained an encoder, denoted Enc-F2G-rare, on 467 out of 816 rare syndromes with more than two and fewer than seven subjects. Ninety percent of the subjects were used to train Enc-F2G-rare and were later assigned to the gallery. The remaining 10% of subjects were assigned to the test set. We then compared the performance of Enc-F2G-rare and Enc-F2G using cosine distance and the softmax classifier.

### Matching undiagnosed patients from unrelated families

We selected 15 articles published from 2015 to 2019 in which GeneMatcher was used to establish an association of a gene with a novel phenotype with facial dysmorphism from unrelated families. In total, these studies contained 108 photos of 91 subjects from 79 families. The details are shown in Table 2. The 15 genes were not among the Face2Gene frequent syndromes, so we can consider them each as a novel phenotype to the model. We performed leave-one-out cross-validation on this dataset; that is, we kept one photo as the test set, and we assigned the rest of the photos to a gallery of 3,533 photos with 816 rare syndromes to simulate the distribution of patients with unknown diagnosis. We then evaluated the performance by top-1 to top-30 rank. If a photo of another subject with the same disease-causing gene from an unrelated family was among the top-*k* rank, we called it a match.

Moreover, we used top-k rank to measure how many unrelated families were connected. If one unrelated family was among the test photo’s top-*k* rank, the families were considered to be connected at that rank. How many families were matched to at least one unrelated family was also represented.

When using the GeneMatcher data, we did not perform syndrome aggregation because aggregation cannot be performed if the syndrome is not known. Instead, we matched patients rather than predicting disorders.

### Syndrome facial distinctiveness score

To evaluate the importance of the facial gestalt for clinical diagnosis of the patient, we asked three dysmorphologists (co-authors Shahida Moosa, Nadja Ehmke, and Karen W. Gripp) to score the usefulness of each syndrome’s facial gestalt for establishing a diagnosis. Three levels were established:

1. Facial gestalt can be supportive in establishing the clinical diagnosis.
2. Facial gestalt is important in establishing the clinical diagnosis, but diagnosis cannot be made without additional clinical features.
3. Facial gestalt is a cardinal symptom, and a visual or clinical diagnosis is possible based only on the facial phenotype.

We then averaged the grades from the three dysmorphologists for each syndrome.

### Syndrome prevalence

The prevalence of each syndrome was collected from Orphanet (www.orpha.net). Birth prevalence was used when the actual prevalence was missing. If only the number of cases or families was available, we calculated the prevalence by summing the numbers of all cases or families and dividing by the global population, using 7.8 billion for the global population and a family size of ten for each family^31^.

### Unseen syndromes correlation analysis

To investigate the influence of prevalence and distinctiveness score on the performance of novel syndromes with facial dysmorphism, we selected 50 frequent syndromes and kept them out of the training set. The 50 syndromes were selected to have evenly distributed distinctiveness scores and prevalence distribution; the distributions are shown in Supplementary Figure 17 and Supplementary Table 6. The encoder (Enc-F2G-exclude-50) was trained on 90% of the subjects from the other 249 frequent syndromes. In addition, we performed random downsampling to remove the confounding effect of prevalence. For each iteration, we randomly downsampled each syndrome by assigning five subjects to the gallery and one subject to the test set. We then averaged the top-10 accuracy of 100 iterations. We calculated Spearman rank correlation coefficients for the following two pairs of data: between top-10 accuracy and the syndrome’s distinctiveness score, and between top-10 accuracy and the prevalence of syndromes collected from Orphanet.

The same analysis was also performed on the GMDB dataset. We selected 20 syndromes from GMDB frequent instead of 50 syndromes because the GMDB dataset is smaller than the Face2Gene dataset, and we trained the Enc-GMDB-exclude-20 on the remaining 119 frequent syndromes. The details of the 20 selected syndromes and the results are reported in Supplementary Table 7. Please note that we report the top-5 accuracy in the GMDB dataset instead of top-10 accuracy because of the smaller number of syndromes in the gallery.

### Analysis of number of training syndromes and subjects

In this analysis, we evaluated the influence of training with additional syndromes and subjects to the novel disorders. To avoid an imbalance among the syndromes, we used the same number of subjects for each syndrome. We first used four different settings for the number of subjects: 10, 20, 40, and 80. However, not all syndromes have the four numbers of subjects we mentioned above for training: for 10, 20, 40, and 80 subjects, there are 242, 156, 84, and 40 syndromes. We then defined the ordering of syndromes we added each time. To add the same syndromes for the four numbers of subjects each time, we first sorted syndromes with the number of subjects in descending order. To avoid bias due to having specific disorders added at each position, we then performed random sorting five times within each of the intervals [1:40], [41, 80], [81, 150], and [151, 240] to generate five different lists of syndromes. Thus, the ordering from common disorders to rare disorders was by interval rather than by syndrome. For example, Kabuki syndrome might be in the 9^th^ position in the first list, but in the 20^th^ position in the second list, but in each randomly sorted list Kabuki syndrome is in the first interval.

For each of five different lists of training syndromes, we performed the same training described as follows. We first trained X number of syndromes with ten subjects, where X = 10 to 240, incremented at an interval of ten syndromes. As mentioned above, there are only 156 syndromes with more than 20 subjects. Thus, we trained syndromes with 20 subjects with X = 10 to 150 syndromes with the same increment of ten syndromes. We performed the same process for 40 and 80 subjects, with maximums of 80 and 40, respectively.

For each setting (number of subjects, number of syndromes), we had five models. We then encoded the photos separately with each model and tested them on the rare syndromes, which had not been seen by the models. In the end, we averaged the performance by the five models and report the top-10 accuracy for each setting in Figure 3. We also used the models described above to encode the GMDB dataset, tested them with the GMDB rare set, and report the results in Supplementary Figure 2.

Because the GMDB dataset is smaller than Face2Gene dataset, we were not able to use the same number of subjects and syndromes to perform the analysis. For the GMDB dataset, we used 10, 20, 40 for the number of subjects, and the syndrome intervals of [1, 10], [11, 40], and [41, 80]. The results of training on GMDB and testing of the GMDB rare set are shown in Supplementary Figure 3.

We next wanted to compare two scenarios, double the number of training syndromes and double the number of training subjects. For example, we first set training on ten subjects for each of ten syndromes as the base setting, then compared this performance to training ten subjects for each of 20 syndromes (double syndromes) and training 20 subjects for each of ten syndromes (double subjects). The base setting had 100 subjects in total. Double syndromes and double subjects each had 200 subjects. This comparison allows us to understand the different influence of adding more syndromes and adding more subjects. The results are shown in Supplementary Figures 4-6.

### Analysis of number of training syndromes in real-world scenario

In this analysis, we trained the encoders with different numbers of syndromes to simulate the real-world scenario. The difference to the previous section is that we used all available subjects with each syndrome for the training. To make a fair comparison, we first used the same ordering of syndromes as in the previous section, and we added a fifth interval of [241, 299]. For each of the five lists of syndromes, we then trained 16 encoders, each with a different number of training syndromes. The interval of syndromes was 20 in this analysis due to the long training time. For example, we used the first ten syndromes in the training list for the first encoder. For the second encoder, we trained on the first 30 syndromes, and continually increased the number of syndromes for each subsequent encoder by 20 until we reached 299 syndromes. Thus, we simulated how syndromes would be included in model training in the real world. We took the rare syndromes as the test set. We then averaged the performance of five models with the same number of training syndromes and report the top-10 accuracy in Supplementary Figure 7.

## Data Availability

This work can be accessed by the gestaltmatcher.org.

https://gestaltmatcher.org

