## Supplementary Materials for "GestaltMatcher: Overcoming the limits of rare disease matching using facial phenotypic descriptors"

### Supplementary Material

Tzung-Chien Hsieh[1,+], Aviram Bar-Haim[2,+], Shahida Moosa[3], Nadja Ehmke[4], Karen W. Gripp[5], Jean Tori Pantel[1,4], Magdalena Danyel[4,6], Martin Atta Mensah[4,7], Denise Horn[4], Stanislav Rosnev[4], Nicole Fleischer[2], Guilherme Bonini[2], Alexander Hustinx[1], Alexander Schmid[1], Alexej Knaus[1], Behnam Javanmardi[1], Hannah Klinkhammer[1,8], Hellen Lesmann[1], Sugirthan Sivalingam[1,8,9], Tom Kamphans[10], Wolfgang Meiswinkel[10], Frédéric Ebstein[11], Elke Krüger[11], Sébastien Küry[12,13], Stéphane Bézieau[12,13], Axel Schmidt[14], Sophia Peters[14], Hartmut Engels[14], Elisabeth Mangold[14], Martina Kreiß[14], Kirsten Cremer[14], Claudia Perne[14], Regina C. Betz[14], Tim Bender[14,15], Kathrin Grundmann-Hauser[16], Tobias B. Haack[16], Matias Wagner[17,18], Theresa Brunet[17], Heidi Beate Bentzen[19], Luisa Averdunk[20], Kimberly Christine Coetzer[3], Gholson J. Lyon[21,22], Malte Spielmann[23], Christian Schaaf[24], Stefan Mundlos[4], Markus M. Nöthen[14], Peter Krawitz[1,*]

[1]Institute for Genomic Statistics and Bioinformatics, University Hospital Bonn, Rheinische Friedrich-Wilhelms-Universität Bonn, Bonn, Germany;

[2]FDNA Inc., Boston, MA, United States;

[3]Division of Molecular Biology and Human Genetics, Stellenbosch University and Medical Genetics, Tygerberg Hospital, Tygerberg, South Africa;

[4]Institute of Medical Genetics and Human Genetics, Charité-Universitätsmedizin Berlin, Humboldt-Universität zu Berlin and Berlin Institute of Health, Berlin, Germany;

[5]A.I. DuPont Hospital for Children/Nemours, Wilmington, DE, USA;

[6]Berlin Center for Rare Diseases, Charité-Universitätsmedizin Berlin, Humboldt-Universität zu Berlin and Berlin Institute of Health, Berlin, Germany;

[7]Berlin Institute of Health (BIH), Berlin, Germany;

[8]Institute for Medical Biometry, Informatics and Epidemiology, Medical Faculty, University of Bonn, Bonn, Germany;

[9]Core Unit for Bioinformatics Data Analysis, Medical Faculty, University of Bonn, Bonn, Germany;

[10]GeneTalk, Bonn, Germany;

[11]Institut für Medizinische Biochemie und Molekularbiologie (IMBM), Universitätsmedizin Greifswald, Greifswald, Germany;

[12]CHU Nantes, Service de Génétique Médicale, Nantes, France;

[13]l'Institut du Thorax, INSERM, CNRS, Université de Nantes, Nantes, France;

[14]Institute of Human Genetics, University of Bonn, Medical Faculty & University Hospital Bonn, Bonn, Germany;

[15]Center for Rare Diseases Bonn, University Hospital Bonn, Bonn, Germany;

[16]Institute of Medical Genetics and Applied Genomics, University of Tübingen, Tübingen, Germany;

[17]Institute of Human Genetics, School of Medicine, Technical University Munich, Munich, Germany;

[18]Institute of Neurogenomics, Helmholtz Zentrum München GmbH, German Research Center for Environmental Health, Neuherberg, Germany;

[19]Norwegian Research Center for Computers and Law, Faculty of Law, University of Oslo, Oslo, Norway;

[20]Institute of Human Genetics and Department of Pediatrics, Medical Faculty, Heinrich Heine University, Düsseldorf, Germany;

[21]Department of Human Genetics and George A. Jervis Clinic, NYS Institute for Basic Research in Developmental Disabilities, Staten Island NY 10314, USA;

[22]Biology PhD Program, The Graduate Center, The City University of New York, New York, United States of America;

[23]Institute of Human Genetics, University of Lübeck, Lübeck, Germany;

[24]Department of Human Genetics, University Hospital of Heidelberg, Heidelberg, Germany;

+ equally contributing first authors

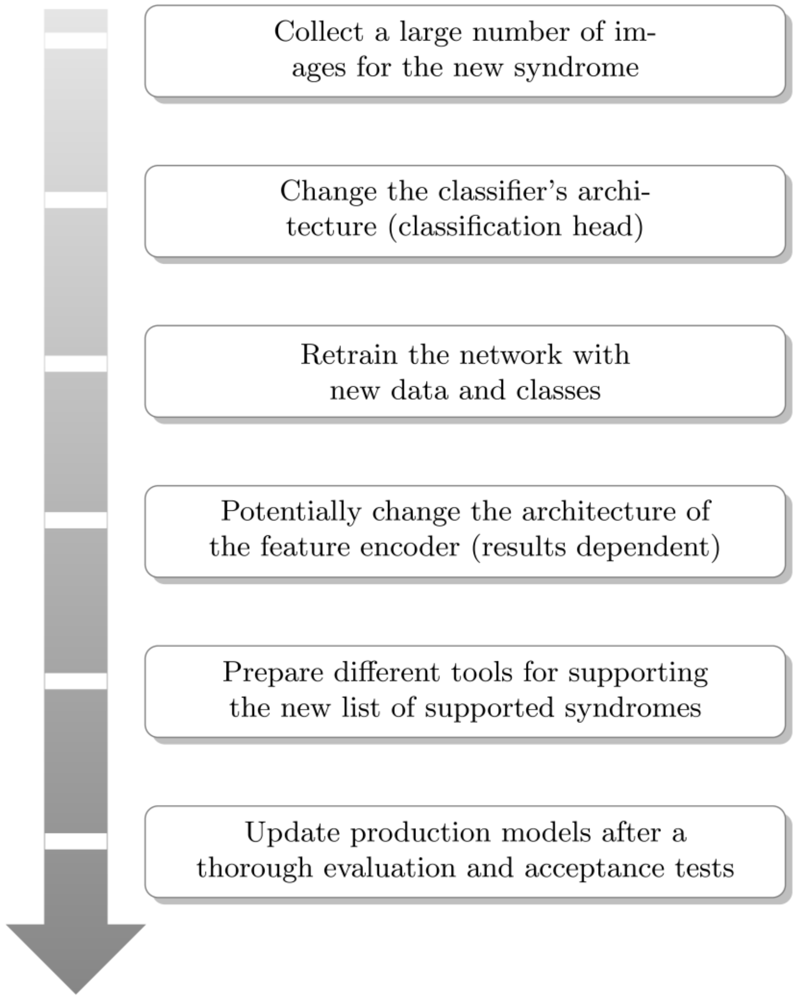


**Supplementary Figure 1: The developmental flow for supporting new syndromes in the DeepGestalt model.** To include new syndromes in an “end-to-end” multi-syndrome classification framework such as DeepGestalt, the developer should go through these six steps. The model retraining might require a lot of money and time, resulting in low scalability for supporting novel diseases or ultra-rare syndromes.

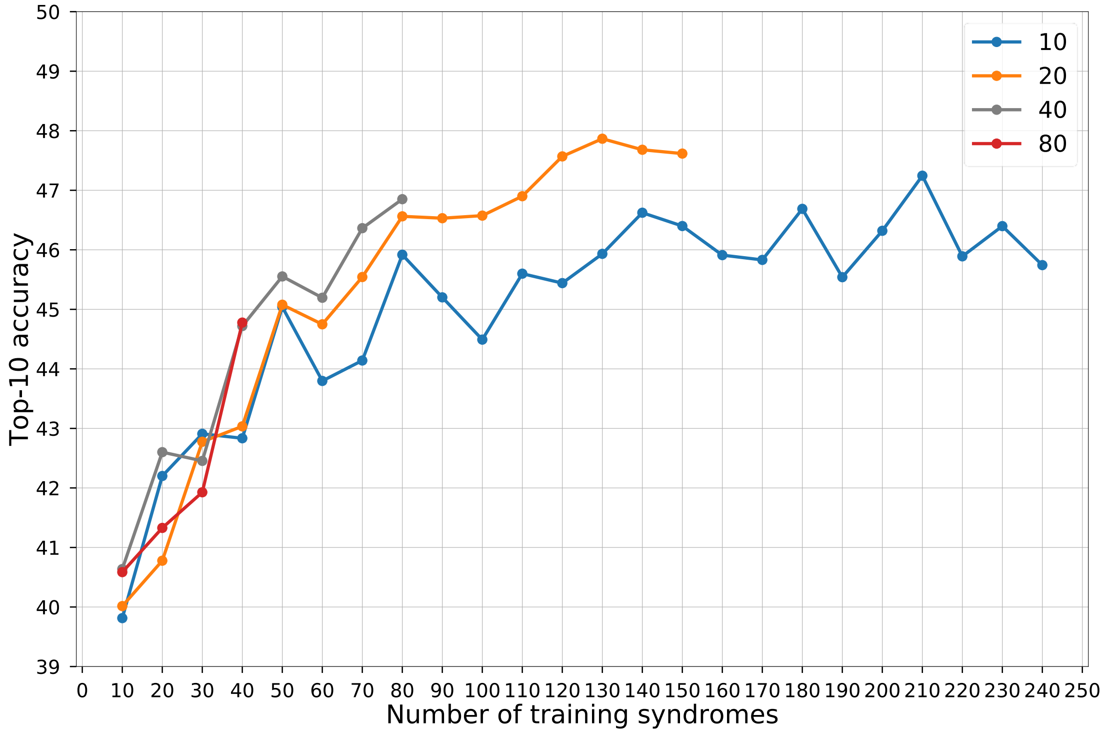


**Supplementary Figure 2: Influence of the number of syndromes from the Face2Gene dataset included in model training and tested on the GMDB rare set.** The x-axis is the number of syndromes from the Face2Gene dataset used in the model training. The models used in this analysis are the same models as in Figure 3. The y-axis shows the average top-10 accuracy of testing on the GMDB rare set. Each line uses the same number of subjects per syndrome, which is shown in the key. For each point, we train the models five times with five different splits and average the results. The null accuracy (the expected value if the encoder returned random predictions) is 8.47% (10/118).

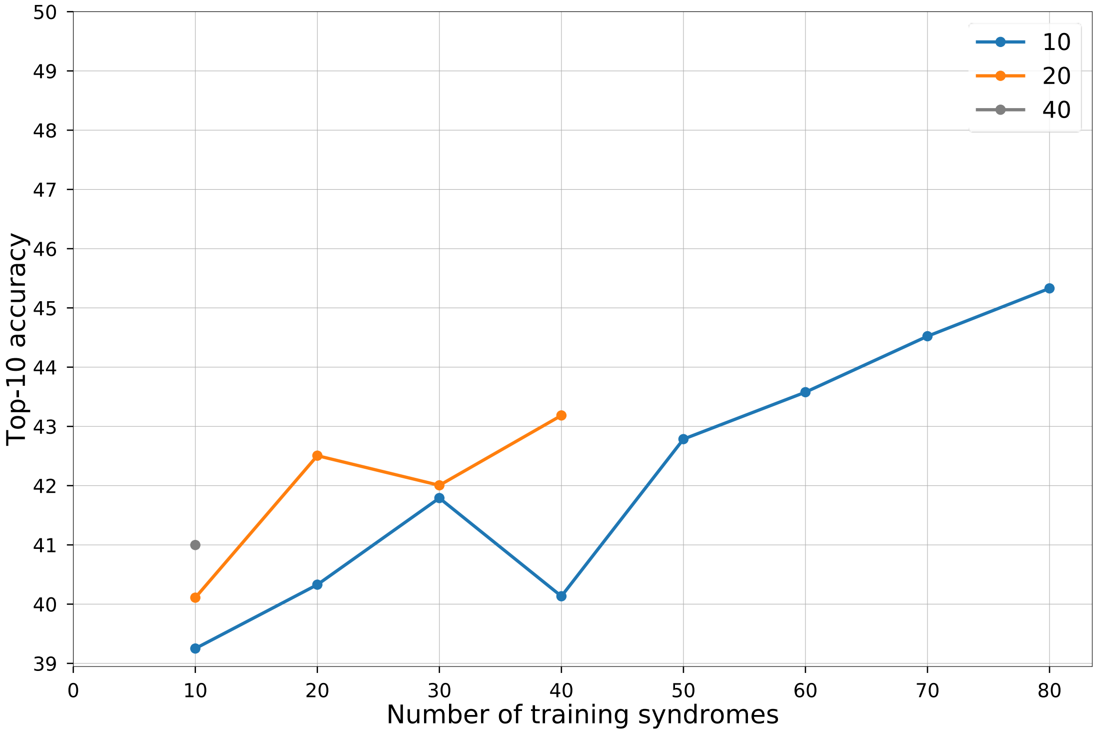


**Supplementary Figure 3: Influence of the number of syndromes from the GMDB dataset included in model training and testing on the GMDB rare set.** The x-axis is the number of syndromes from the GMDB dataset used in model training. The models used in this analysis are the same models as in Figure 3. The y-axis shows the average top-10 accuracy of testing on the rare set. Each line uses the same number of subjects per syndrome, which is shown in the key. For each point, we train the models five times with five different splits and average the results. The null accuracy is 8.47% (10/118).

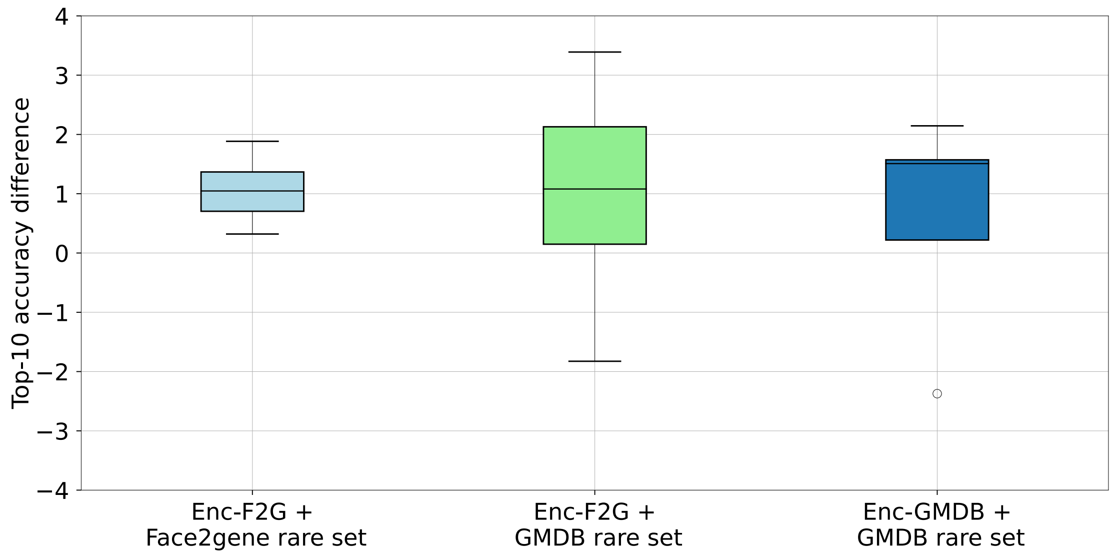


**Supplementary Figure 4: Pairwise improvement difference between double syndromes and double subjects.** Pairwise improvement was calculated by subtracting the improvement of double subjects from the improvement of double syndromes. Taking Supplementary Figure 3 as an example, the base accuracy of 10 subjects and 10 syndromes (10, 10) is 39.25%. The double syndromes of the point (10, 10) is the point of 10 subjects and 20 syndromes (10, 20) which is 40.33%. The double subjects of the point (10, 10) is the point of 20 subjects and 10 syndromes (20, 10) which is 40.11%. Then the difference between (10, 20) and (20, 10) is 0.22%, and 0.22% is a dot in the third column. The three columns were calculated by all the combinations in the three settings (Figure 3, Supplementary Figures 2 and 3). We can also think about the double syndromes as going right in the graphs shown in Figure 3 and Supplementary Figures 2 and 3 (including more new syndromes), and double subjects as going up (adding more subjects to existing syndromes).

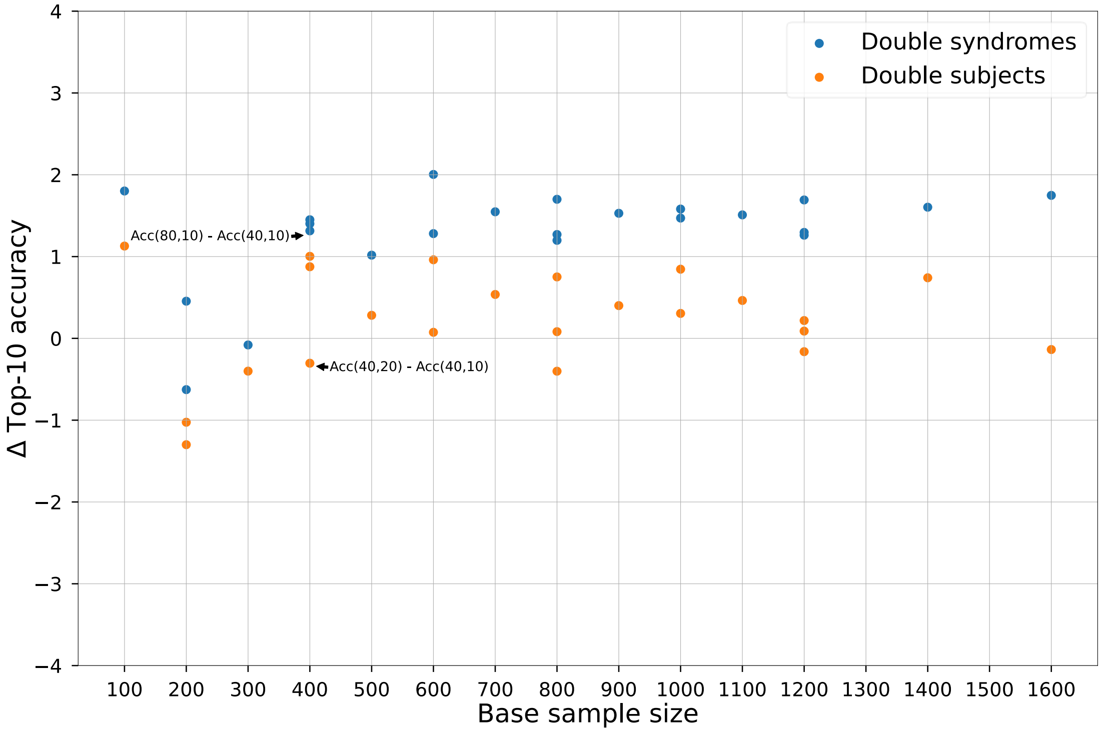


**Supplementary Figure 5: Performance improvement of double syndromes and double subjects when using different base sample sizes with Face2Gene models and the Face2Gene rare set.** Base sample size is calculated by the number of subjects multiplied by the number of syndromes. For example, the point of 40 subjects and 10 syndromes has sample size of 400, and it equals the point of 10 subjects and 40 syndromes, and the point of 20 subjects and 20 syndromes. $\Delta$Top-10 accuracy is the difference of accuracy of double syndromes/subjects to the base point, and is calculated based on Figure 3. Take the two points annotated in the figure as two examples. The base point is 10 subjects and 40 syndromes with sample size 400. The upper point is subtracting the point of 10 subjects and 40 syndromes from point of 10 subjects and 80 syndromes in Figure 3. The lower point is subtracting the point of 10 subjects and 40 syndromes from point of 20 subjects and 40 syndromes in Figure 3. When the base sample size grows, the effect of double syndromes becomes more important because it becomes the indicator of how we decide adding more data into the training set (adding more syndromes or adding ore subjects to existed syndromes).

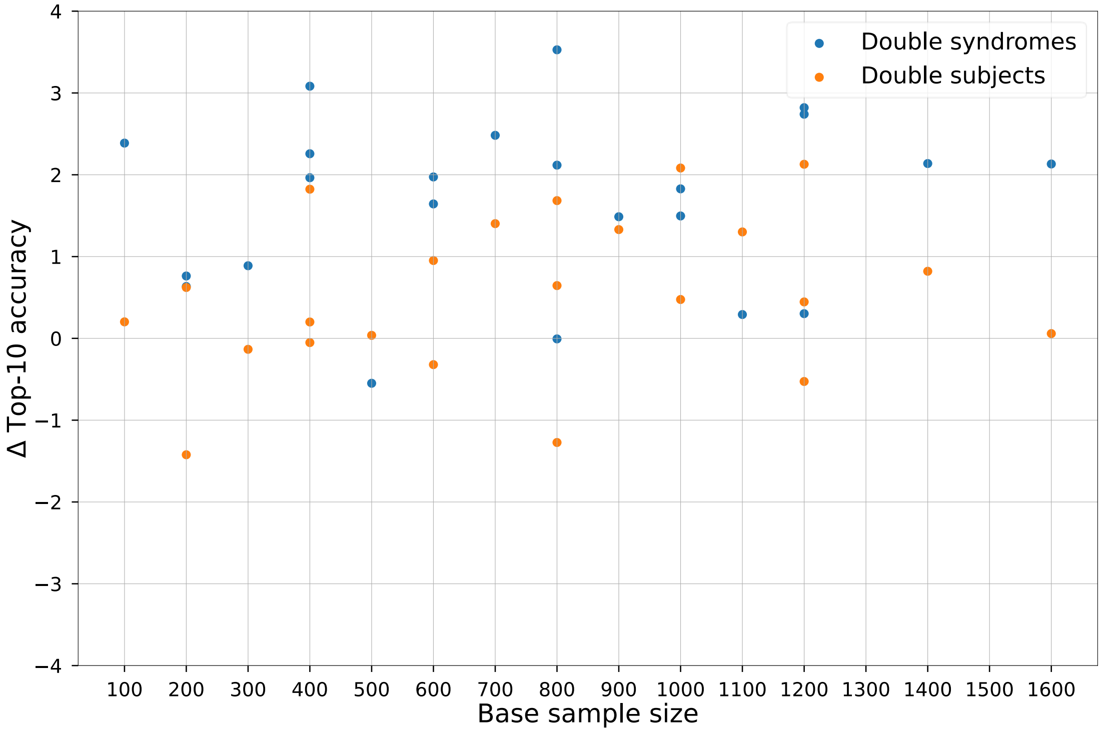


**Supplementary Figure 6: Performance improvement of double syndromes and double subjects when using different base sample size on Face2Gene models and the GMDB rare set.** This figure is calculated as described in Supplementary Figure 5, and is calculated based on Supplementary Figure 2.

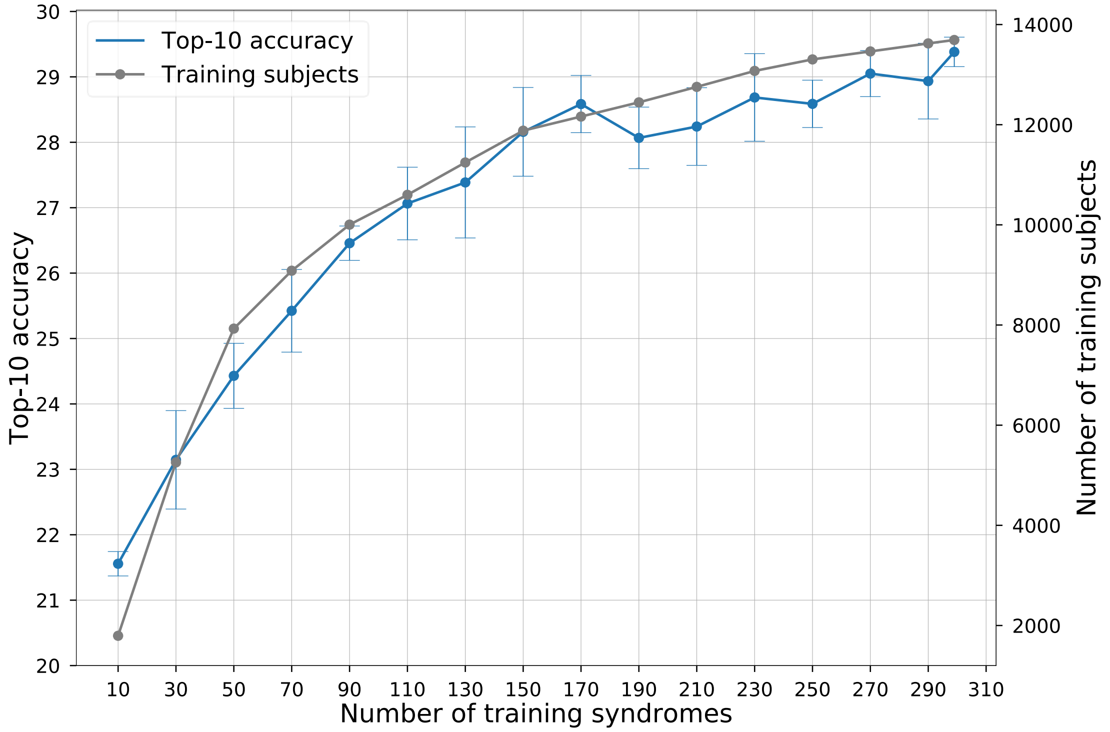


**Supplementary Figure 7: Influence of the number of syndromes included in model training.** The x-axis is the number of syndromes used in model training. The left y-axis shows the average top-10 accuracy for five models, and the error bars show standard deviation. The right y-axis is the cumulative number of subjects in the training syndromes. Each point is the average of testing five different models with different data splits. The null accuracy is 1.23% (10/816).

a

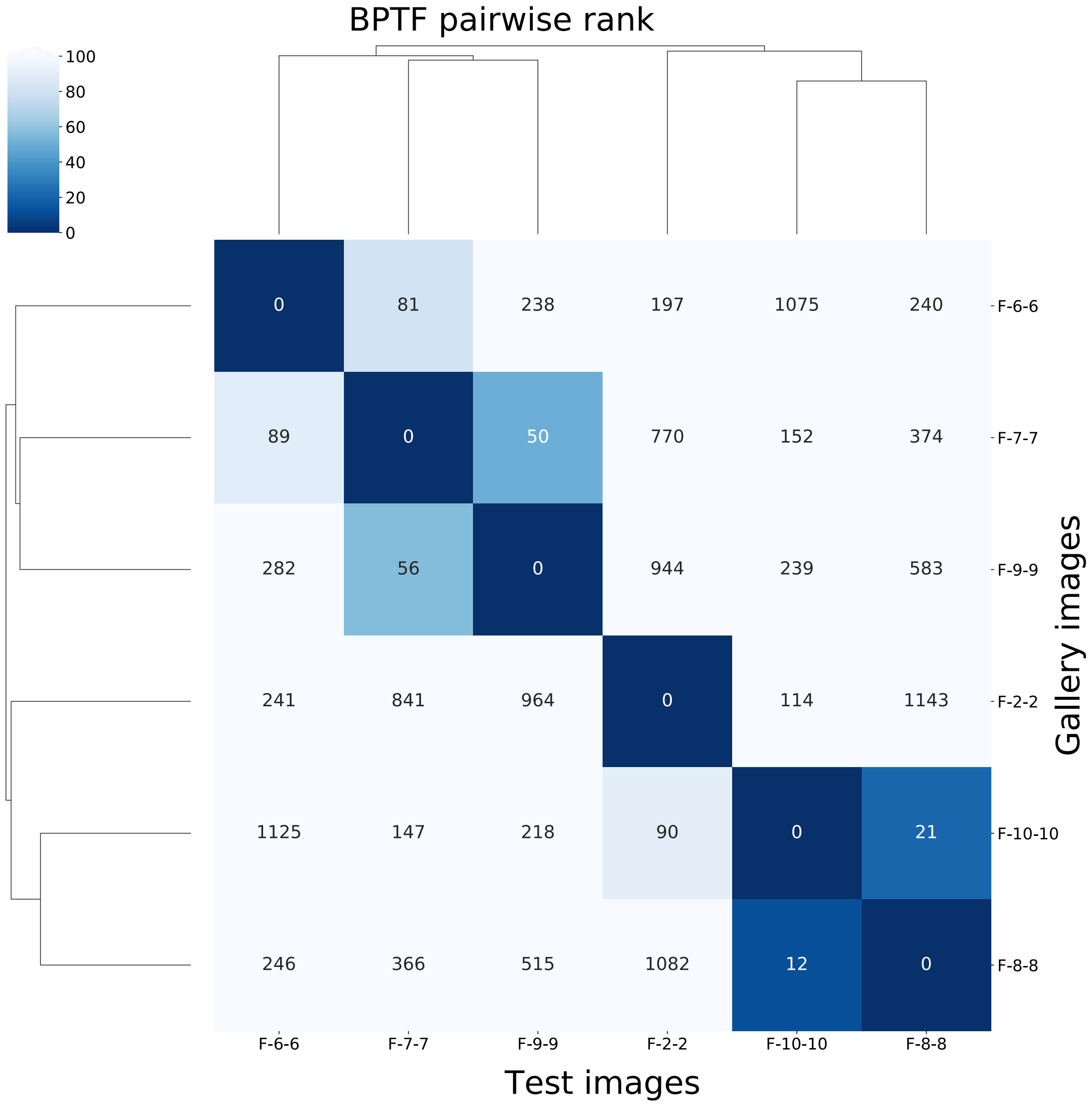

BPTF pairwise rank
100
80
60
40
20
0
0 81 238 197 1075 240 F-6-6
89 0 50 770 152 374 F-7-7
282 56 0 944 239 583 F-9-9
241 841 964 0 114 1143 F-2-2
1125 147 218 90 0 21 F-10-10
246 366 515 1082 12 0 F-8-8
F-6-6 F-7-7 F-9-9 F-2-2 F-10-10 F-8-8
Test images
Gallery images

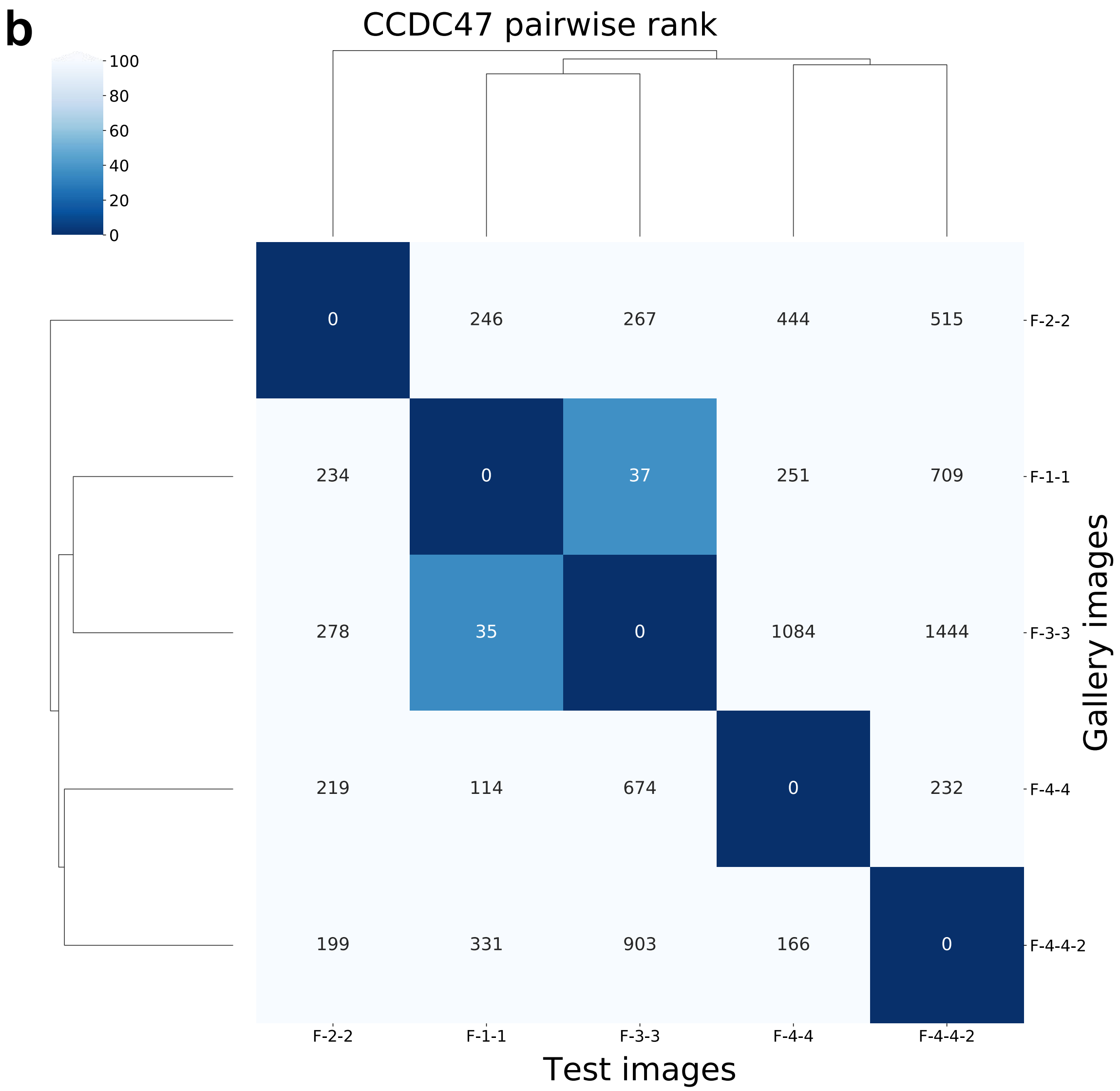
b
CCDC47 pairwise rank
100
80
60
40
20
0
0
246
267
444
515
234
0
37
251
709
278
35
0
1084
1444
219
114
674
0
232
199
331
903
166
0
F-2-2
F-1-1
F-3-3
F-4-4
F-4-4-2
Gallery images
F-2-2
F-1-1
F-3-3
F-4-4
F-4-4-2
Test images

c

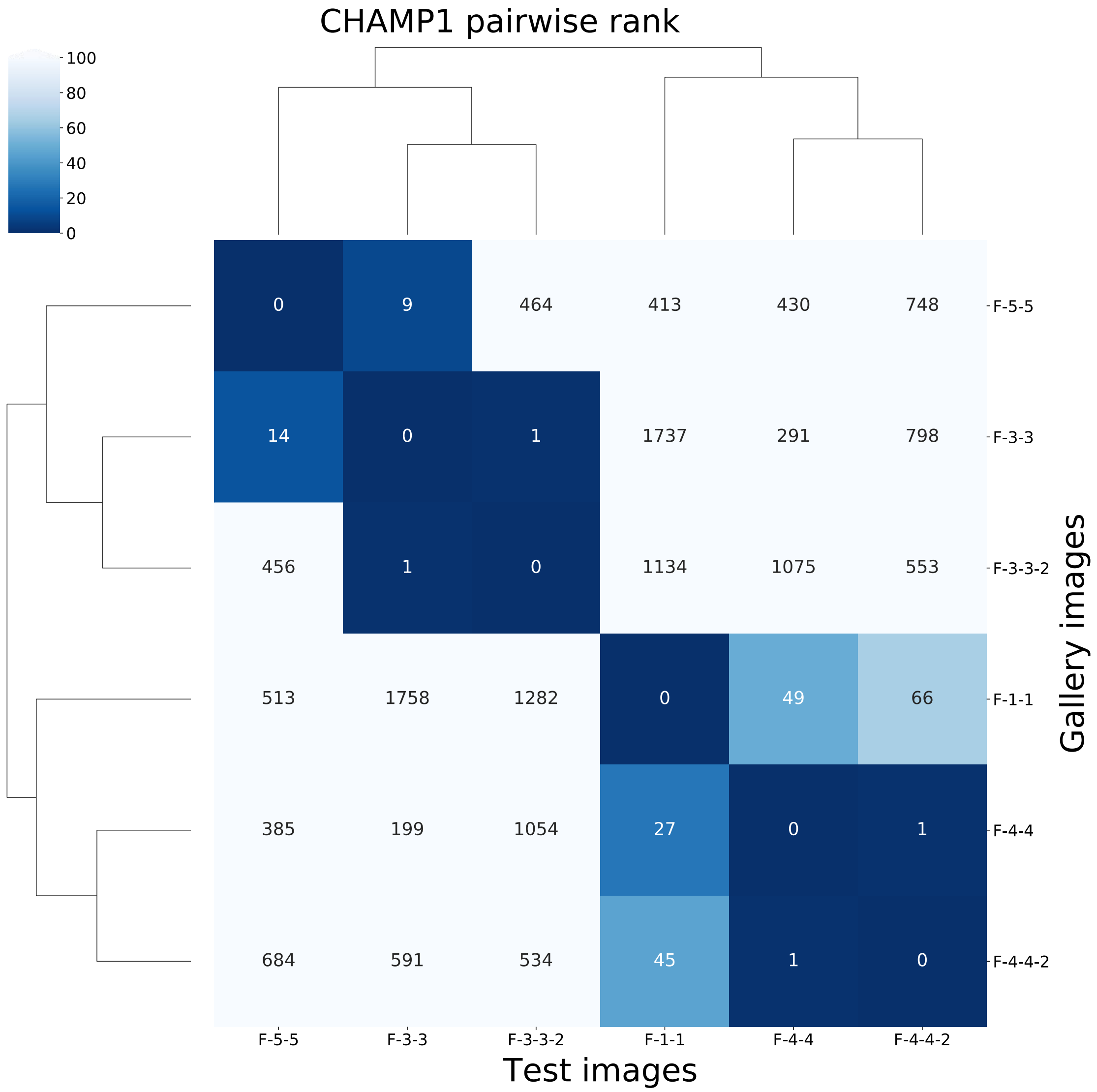
CHAMP1 pairwise rank
100
80
60
40
20
0
0 9 464 413 430 748 F-5-5
14 0 1 1737 291 798 F-3-3
456 1 0 1134 1075 553 F-3-3-2
513 1758 1282 0 49 66 F-1-1
385 199 1054 27 0 1 F-4-4
684 591 534 45 1 0 F-4-4-2
F-5-5
F-3-3
F-3-3-2
F-1-1
F-4-4
F-4-4-2
Test images
Gallery images

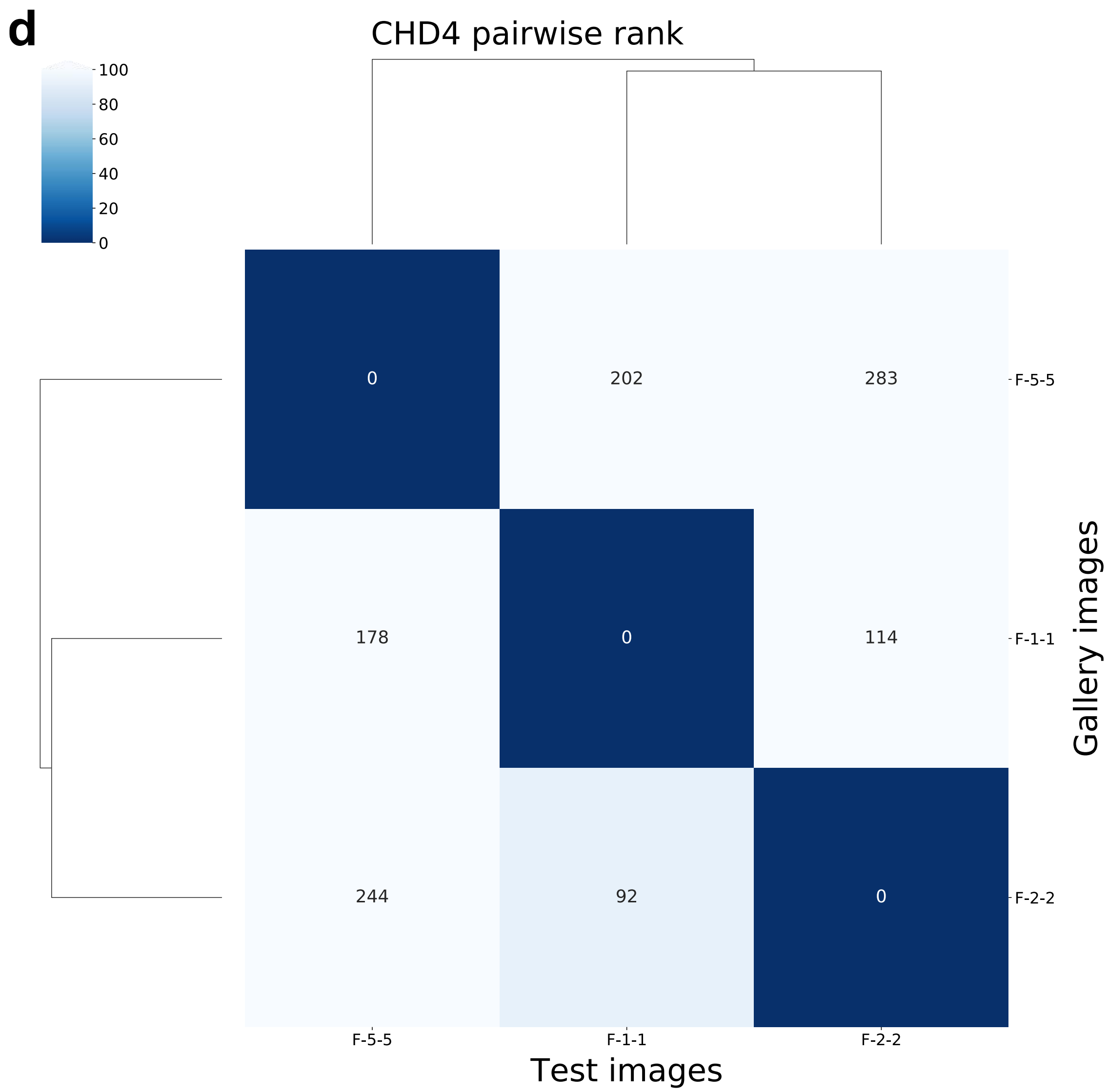
d
CHD4 pairwise rank
100
80
60
40
20
0
0
202
283
178
0
114
244
92
0
F-5-5
F-1-1
F-2-2
Gallery images
F-5-5
F-1-1
F-2-2
Test images

e

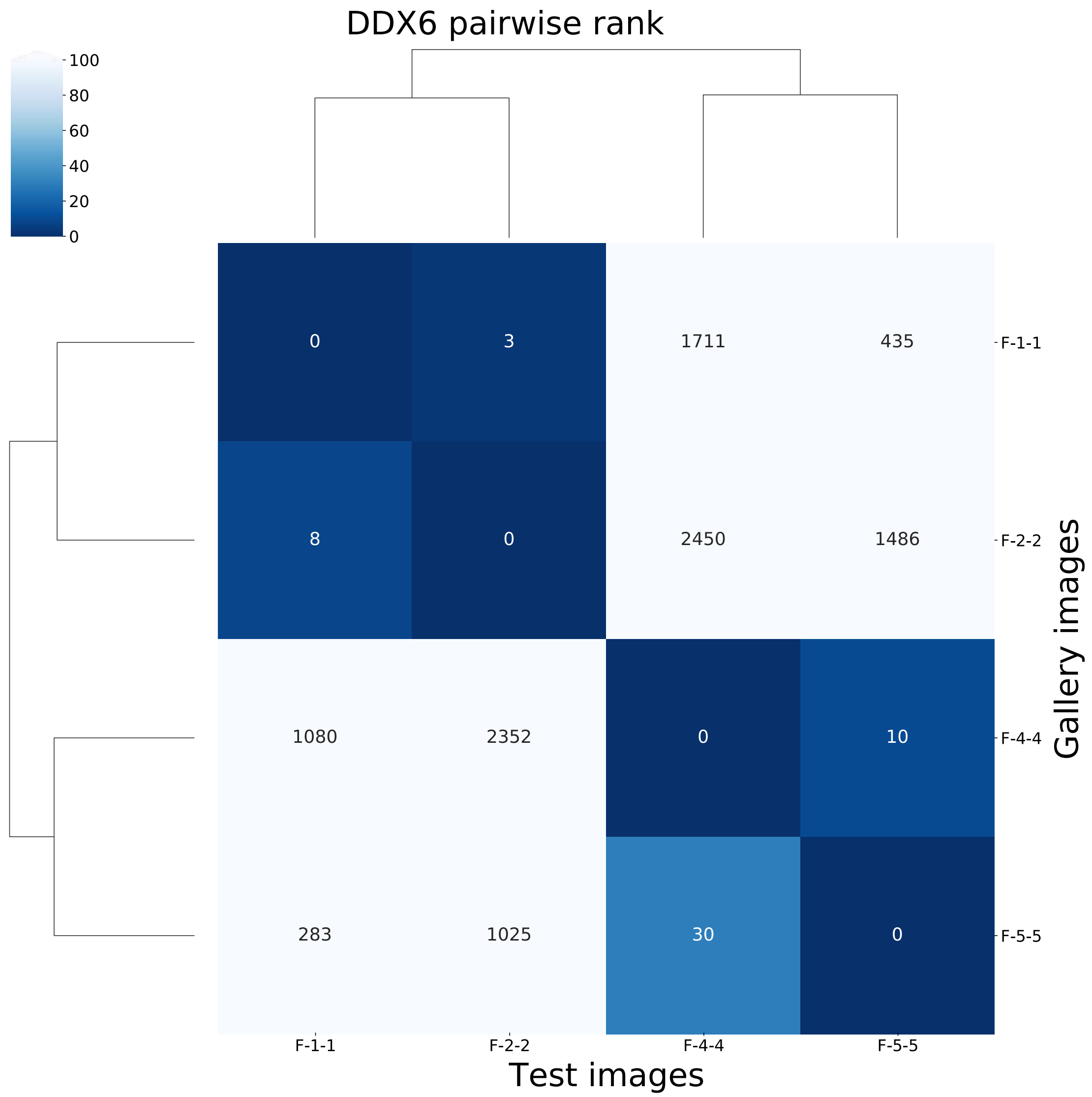

DDX6 pairwise rank
100
80
60
40
20
0
0
3
1711
435
8
0
2450
1486
1080
2352
0
10
283
1025
30
0
F-1-1
F-2-2
F-4-4
F-5-5
Gallery images
F-1-1
F-2-2
F-4-4
F-5-5
Test images

**f**

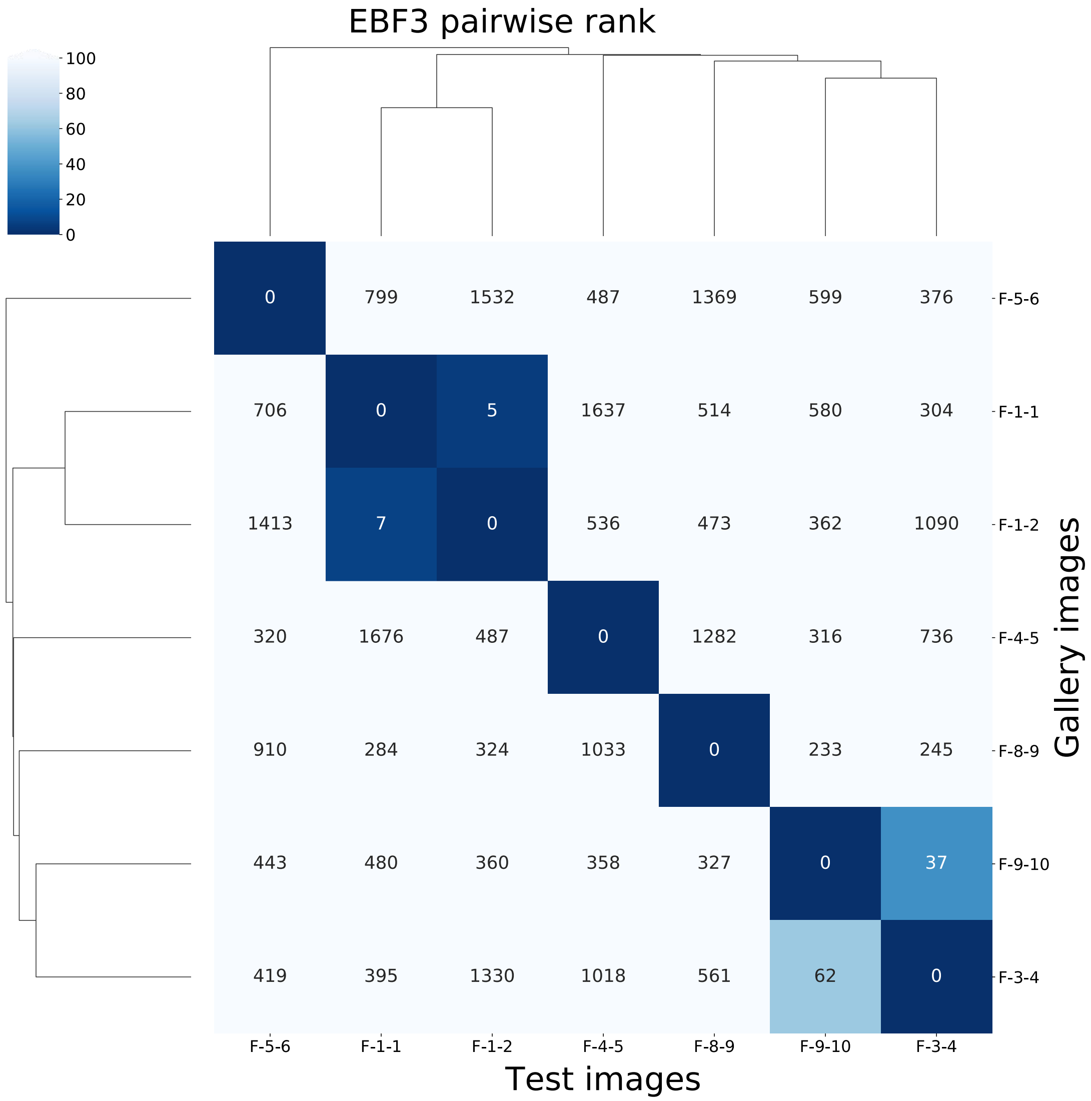

EBF3 pairwise rank
100
80
60
40
20
0
0 799 1532 487 1369 599 376
706 0 5 1637 514 580 304
1413 7 0 536 473 362 1090
320 1676 487 0 1282 316 736
910 284 324 1033 0 233 245
443 480 360 358 327 0 37
419 395 1330 1018 561 62 0
F-5-6
F-1-1
F-1-2
F-4-5
F-8-9
F-9-10
F-3-4
Gallery images
F-5-6 F-1-1 F-1-2 F-4-5 F-8-9 F-9-10 F-3-4
Test images

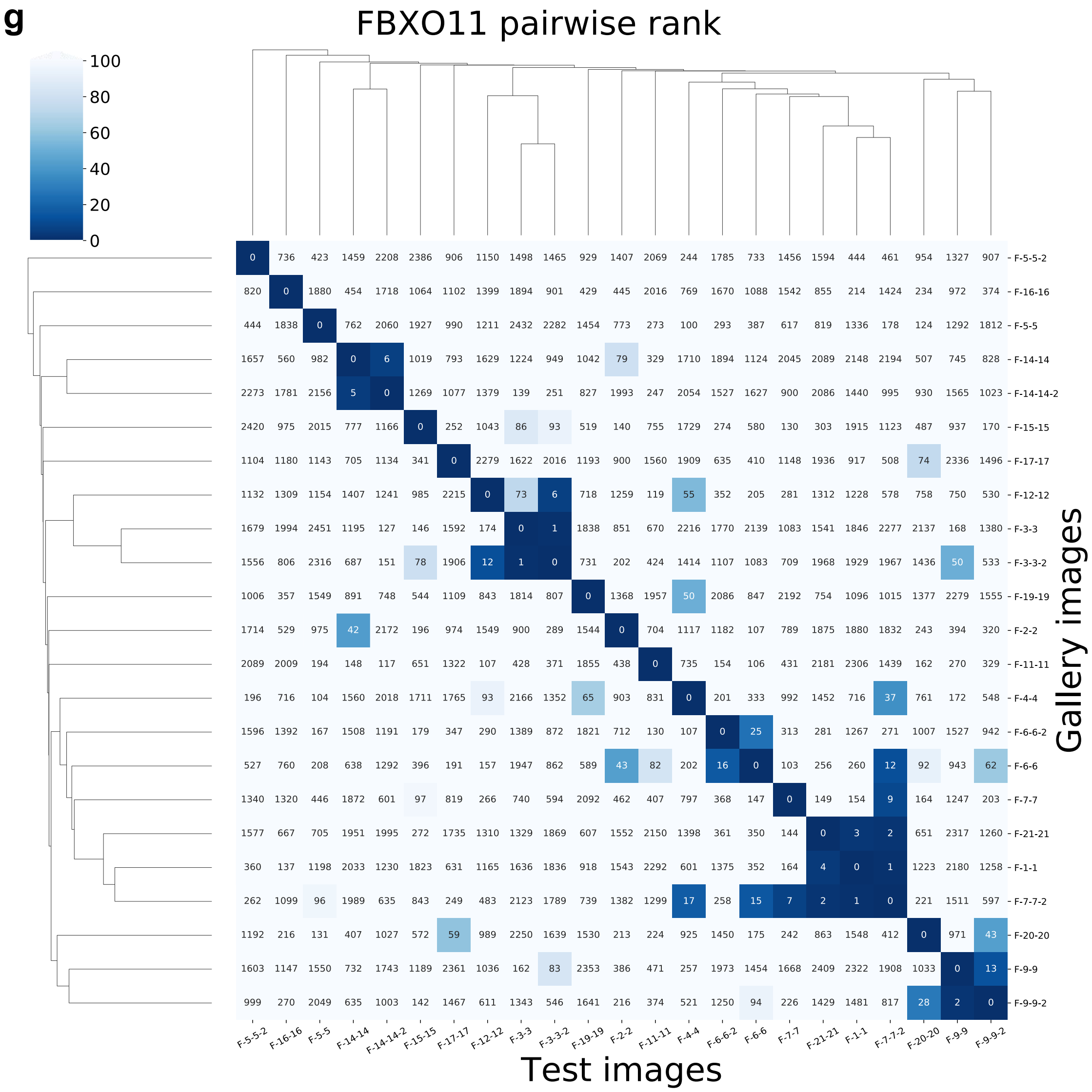
g
FBXO11 pairwise rank
100
80
60
40
20
0
Gallery images
Test images
F-5-5-2
F-16-16
F-5-5
F-14-14
F-14-14-2
F-15-15
F-17-17
F-12-12
F-3-3
F-3-3-2
F-19-19
F-2-2
F-11-11
F-4-4
F-6-6-2
F-6-6
F-7-7
F-21-21
F-1-1
F-7-7-2
F-20-20
F-9-9
F-9-9-2

h

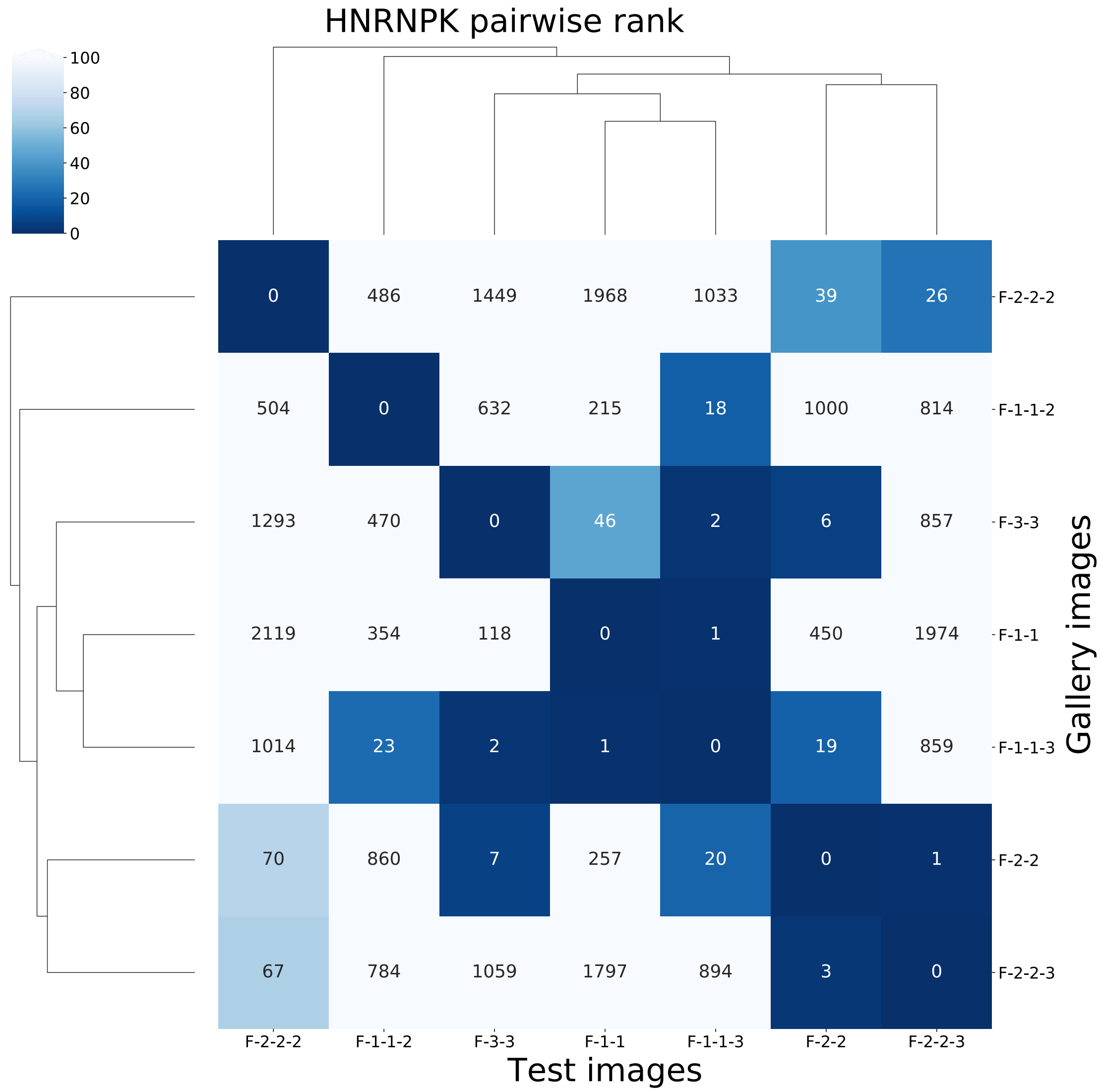
HNRNPK pairwise rank
100
80
60
40
20
0
0 486 1449 1968 1033 39 26 F-2-2-2
504 0 632 215 18 1000 814 F-1-1-2
1293 470 0 46 2 6 857 F-3-3
2119 354 118 0 1 450 1974 F-1-1
1014 23 2 1 0 19 859 F-1-1-3
70 860 7 257 20 0 1 F-2-2
67 784 1059 1797 894 3 0 F-2-2-3
F-2-2-2 F-1-1-2 F-3-3 F-1-1 F-1-1-3 F-2-2 F-2-2-3
Test images
Gallery images

i

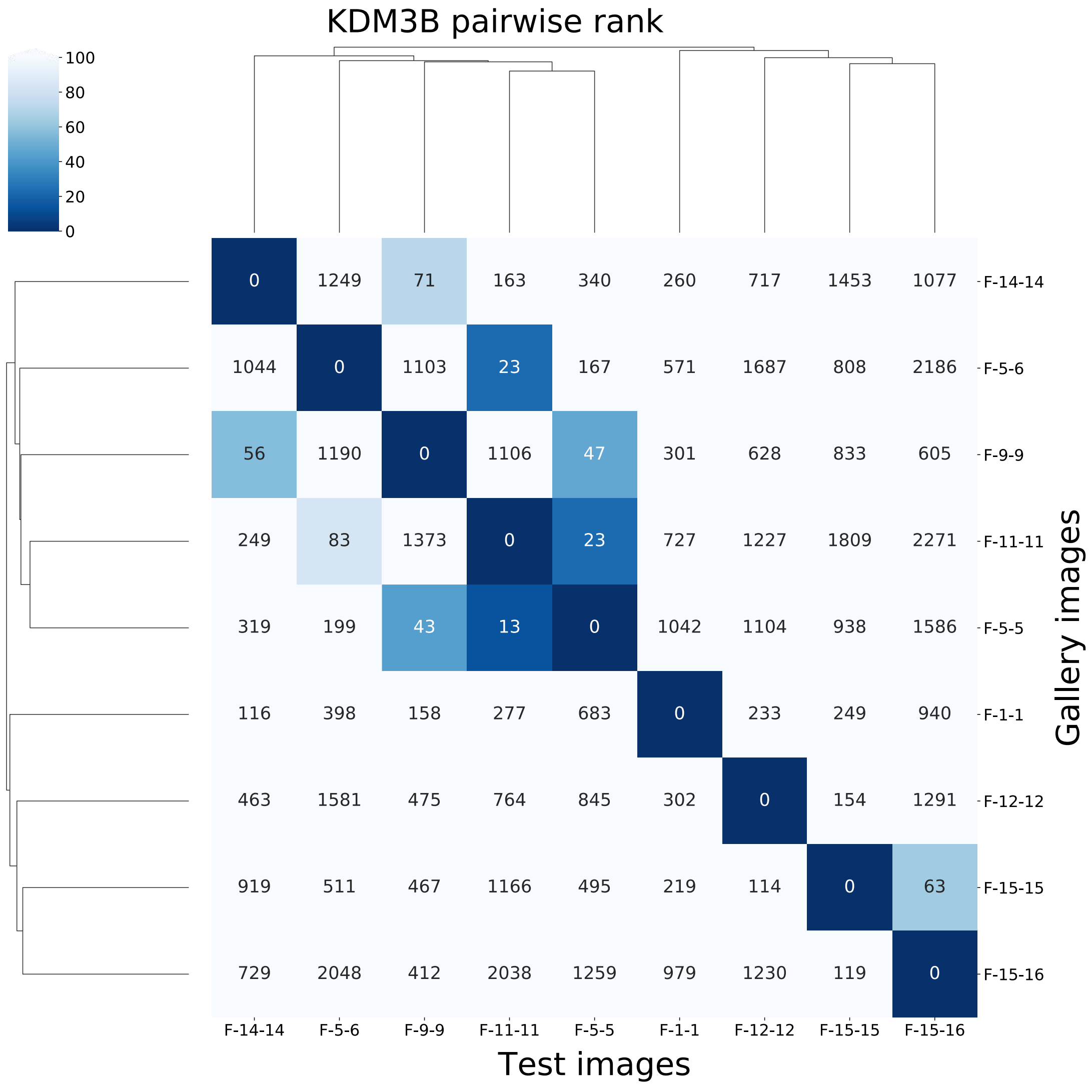
KDM3B pairwise rank
100
80
60
40
20
0
0 1249 71 163 340 260 717 1453 1077 F-14-14
1044 0 1103 23 167 571 1687 808 2186 F-5-6
56 1190 0 1106 47 301 628 833 605 F-9-9
249 83 1373 0 23 727 1227 1809 2271 F-11-11
319 199 43 13 0 1042 1104 938 1586 F-5-5
116 398 158 277 683 0 233 249 940 F-1-1
463 1581 475 764 845 302 0 154 1291 F-12-12
919 511 467 1166 495 219 114 0 63 F-15-15
729 2048 412 2038 1259 979 1230 119 0 F-15-16
F-14-14 F-5-6 F-9-9 F-11-11 F-5-5 F-1-1 F-12-12 F-15-15 F-15-16
Test images
Gallery images

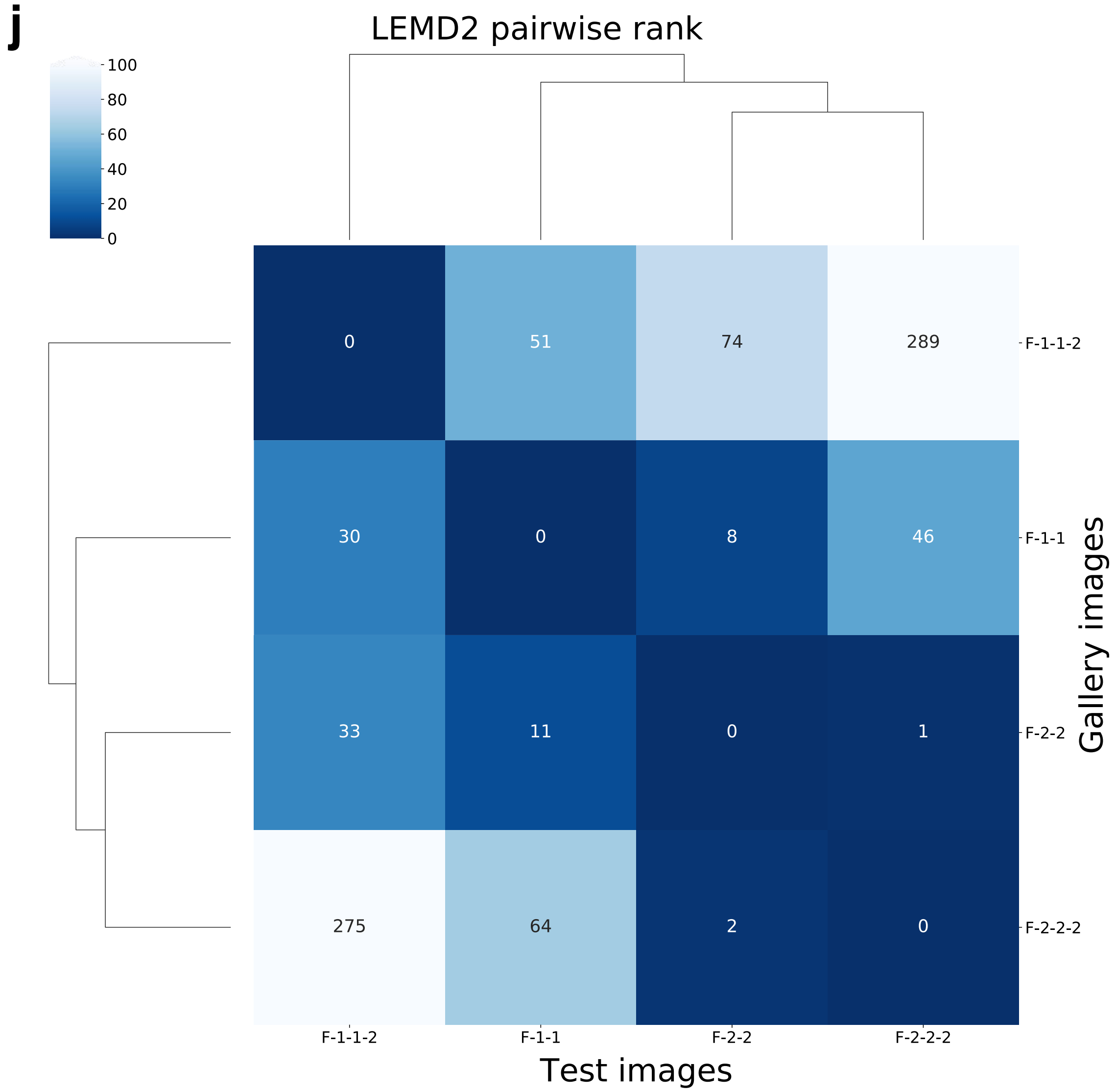
j
LEMD2 pairwise rank
100
80
60
40
20
0
0
51
74
289
30
0
8
46
33
11
0
1
275
64
2
0
F-1-1-2
F-1-1
F-2-2
F-2-2-2
Gallery images
F-1-1-2
F-1-1
F-2-2
F-2-2-2
Test images

k

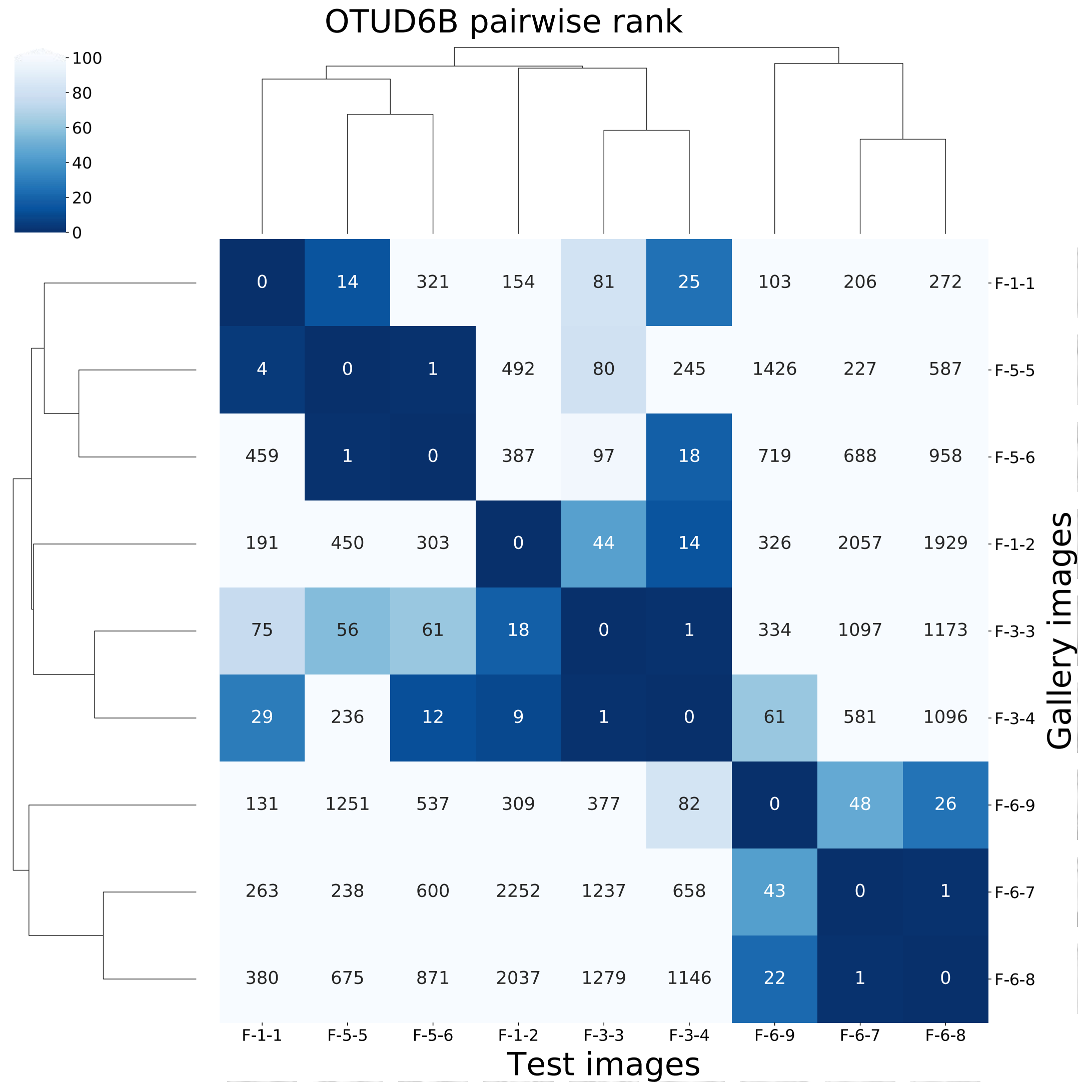


| | F-1-1 | F-5-5 | F-5-6 | F-1-2 | F-3-3 | F-3-4 | F-6-9 | F-6-7 | F-6-8 |
|---|---|---|---|---|---|---|---|---|---|
| F-1-1 | 0 | 14 | 321 | 154 | 81 | 25 | 103 | 206 | 272 |
| F-5-5 | 4 | 0 | 1 | 492 | 80 | 245 | 1426 | 227 | 587 |
| F-5-6 | 459 | 1 | 0 | 387 | 97 | 18 | 719 | 688 | 958 |
| F-1-2 | 191 | 450 | 303 | 0 | 44 | 14 | 326 | 2057 | 1929 |
| F-3-3 | 75 | 56 | 61 | 18 | 0 | 1 | 334 | 1097 | 1173 |
| F-3-4 | 29 | 236 | 12 | 9 | 1 | 0 | 61 | 581 | 1096 |
| F-6-9 | 131 | 1251 | 537 | 309 | 377 | 82 | 0 | 48 | 26 |
| F-6-7 | 263 | 238 | 600 | 2252 | 1237 | 658 | 43 | 0 | 1 |
| F-6-8 | 380 | 675 | 871 | 2037 | 1279 | 1146 | 22 | 1 | 0 |

I

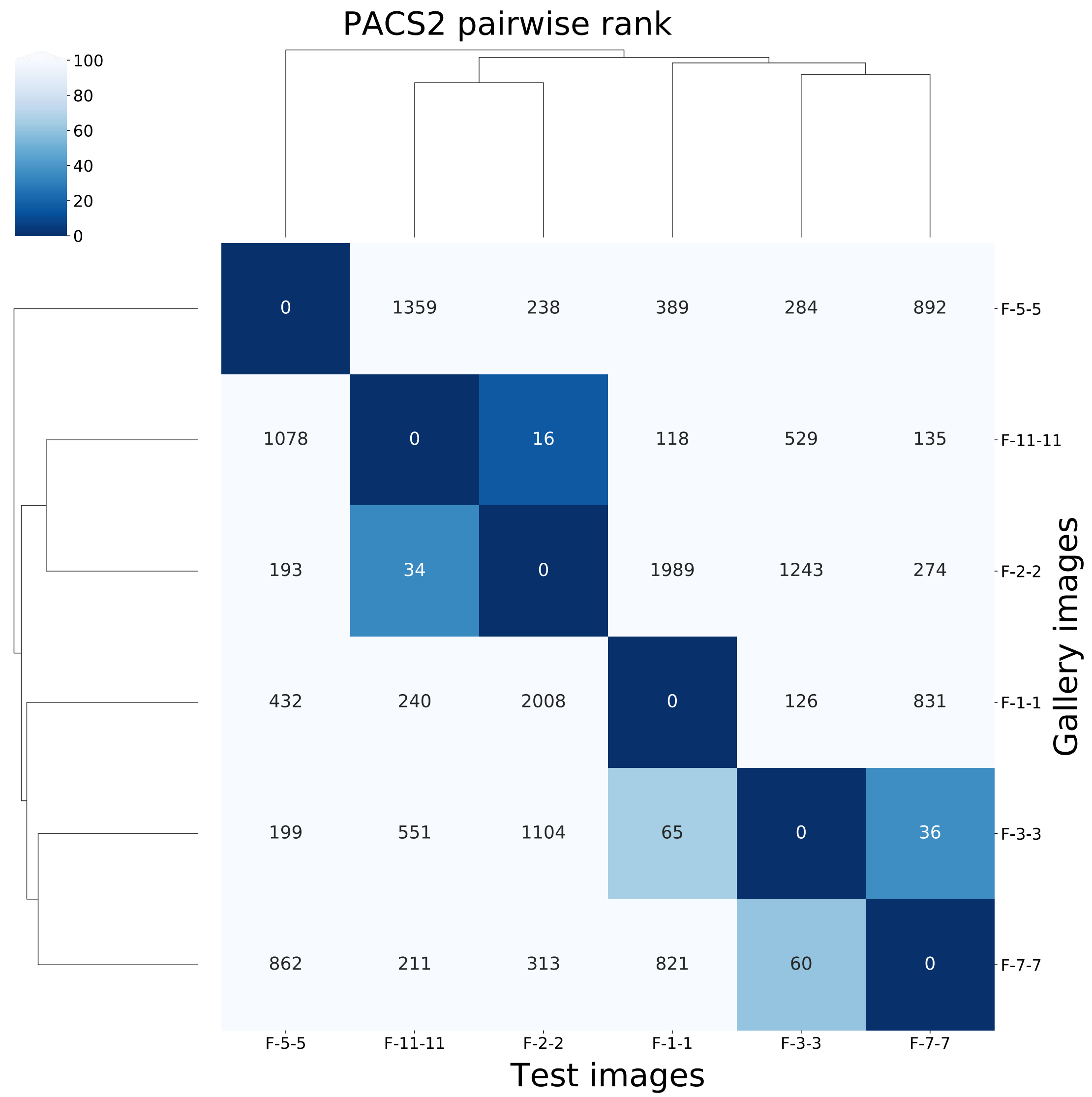

PACS2 pairwise rank
100
80
60
40
20
0
0 1359 238 389 284 892 F-5-5
1078 0 16 118 529 135 F-11-11
193 34 0 1989 1243 274 F-2-2
432 240 2008 0 126 831 F-1-1
199 551 1104 65 0 36 F-3-3
862 211 313 821 60 0 F-7-7
F-5-5 F-11-11 F-2-2 F-1-1 F-3-3 F-7-7
Test images
Gallery images

**m**

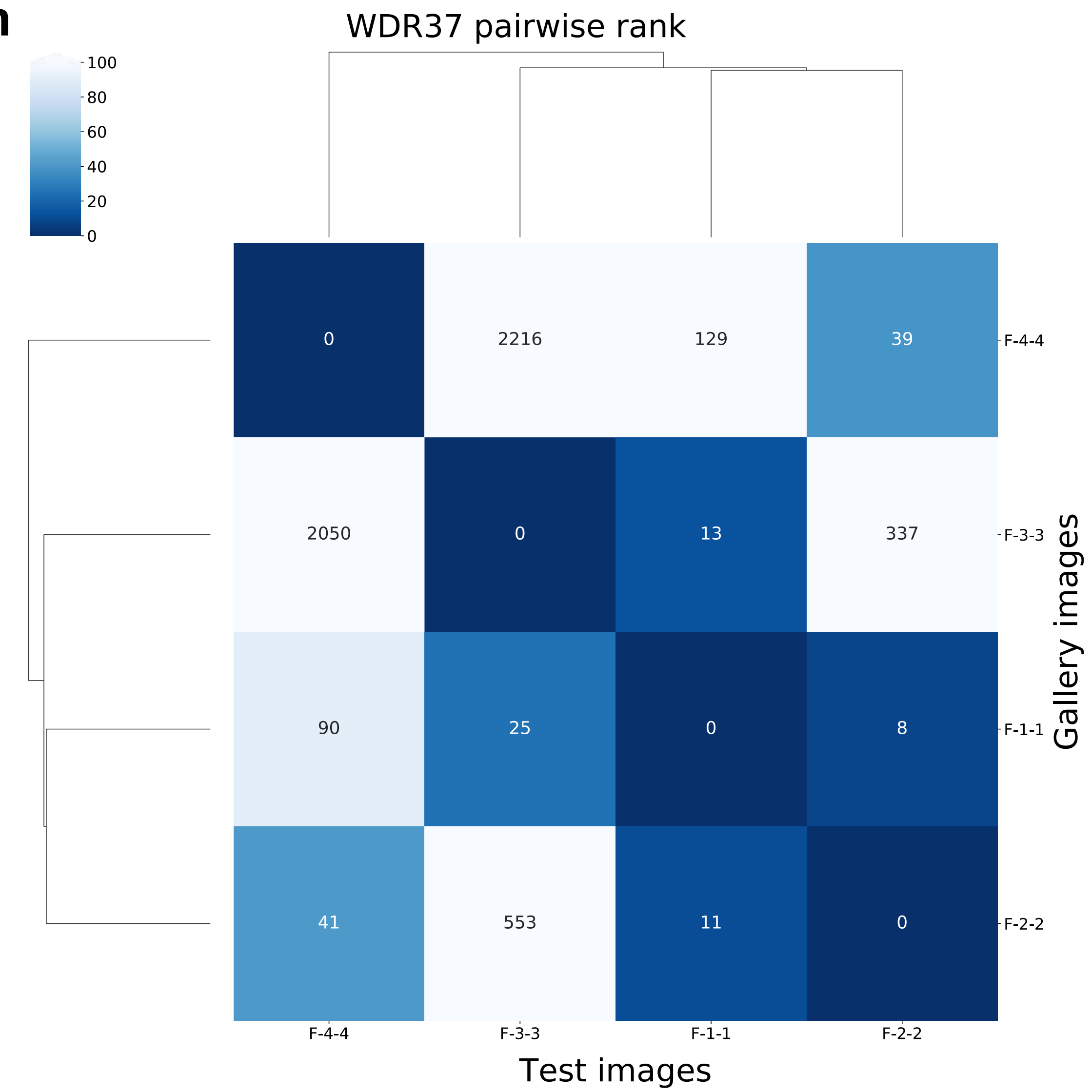

WDR37 pairwise rank
100
80
60
40
20
0
0
2216
129
39
2050
0
13
337
90
25
0
8
41
553
11
0
F-4-4
F-3-3
F-1-1
F-2-2
Gallery images
F-4-4
F-3-3
F-1-1
F-2-2
Test images

**n**

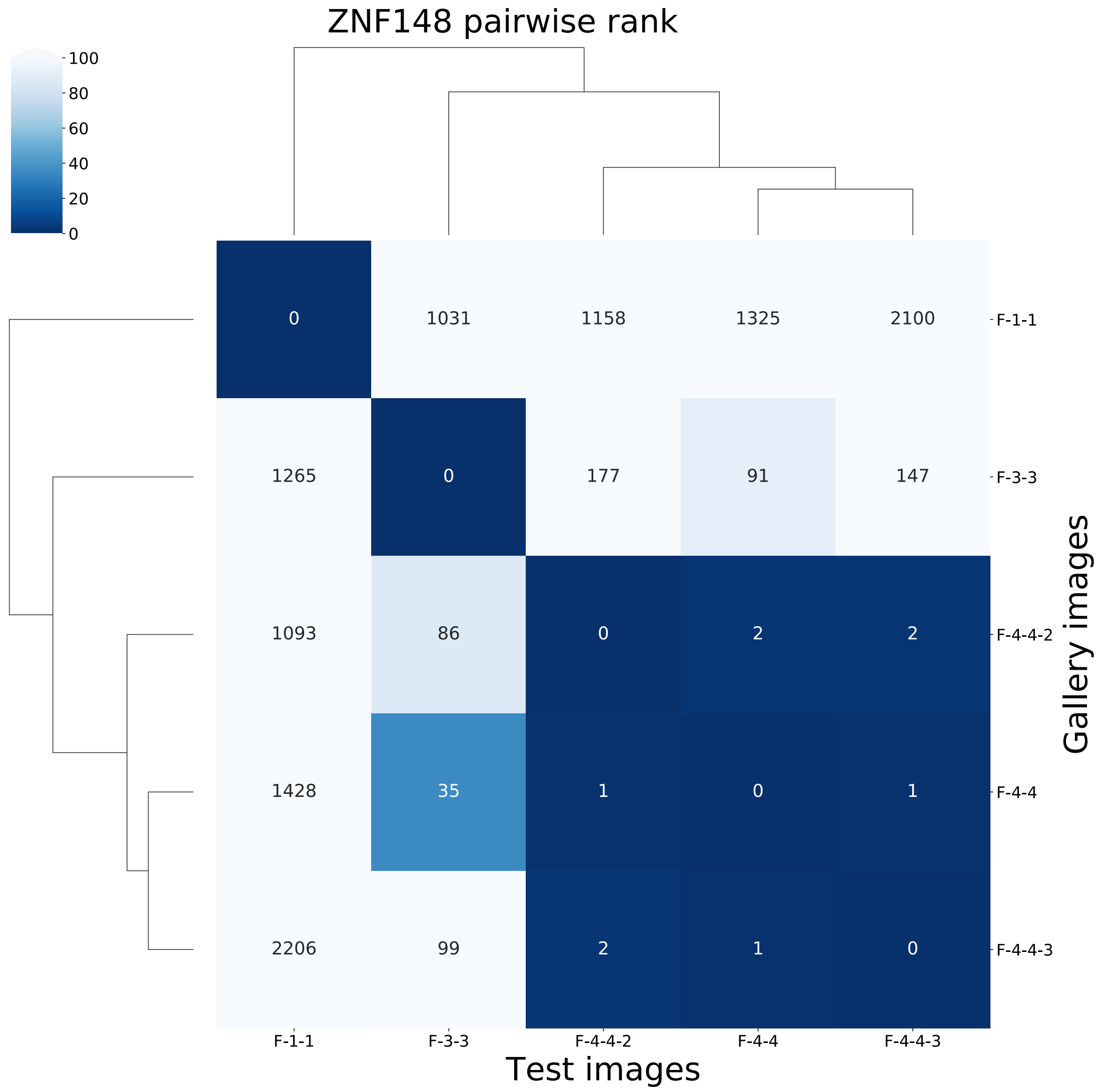
ZNF148 pairwise rank
100
80
60
40
20
0
0
1031
1158
1325
2100
1265
0
177
91
147
1093
86
0
2
2
1428
35
1
0
1
2206
99
2
1
0
F-1-1
F-3-3
F-4-4-2
F-4-4
F-4-4-3
F-1-1
F-3-3
F-4-4-2
F-4-4
F-4-4-3
Test images
Gallery images

**Supplementary Figure 8: Pairwise rank of 14 GeneMatcher validation studies (excluding *TMEM96*).** Each subfigure is the pairwise rank of one GeneMatcher validation study. The label for each image consists of a family component and a subject component. The subject numbering is the same as the numbering in the original publication[1–14]. If there were multiple photos from the same person, we added an image number to the end of its label; for example, F-4-4-2 means the fourth family, fourth subject, second photo.

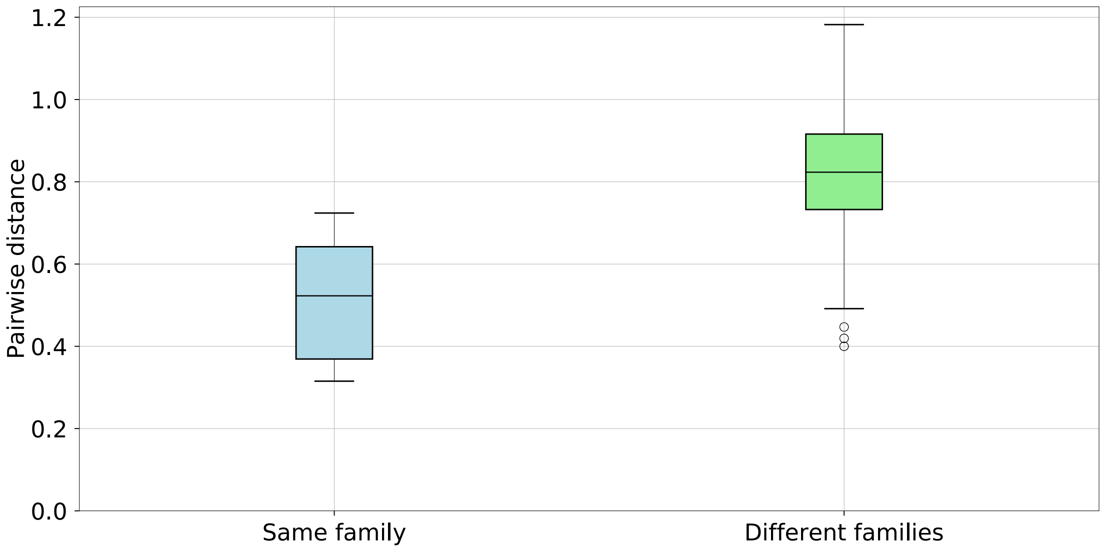


**Supplementary Figure 9: Comparison of the pairwise distance distribution between subjects in the same family and subjects in different families with the same disease-causing gene.** The median distance within affected individuals from the same family is 0.522, and the median distance between different families is 0.823.

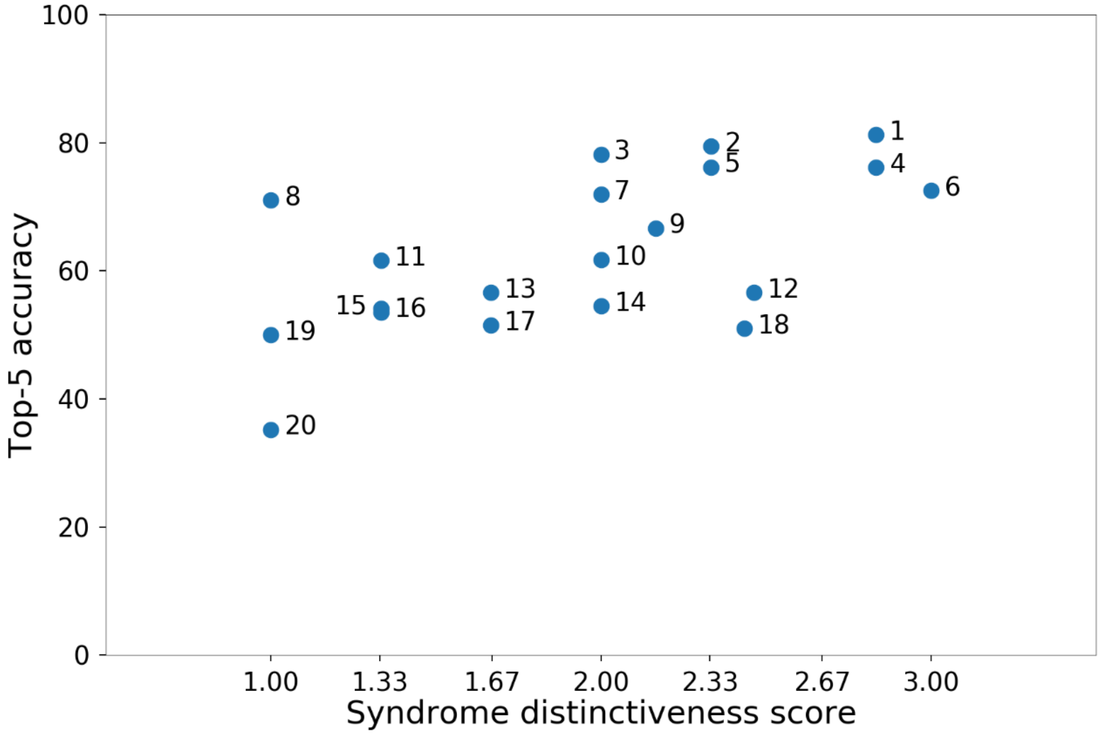


**Supplementary Figure 10: Correlation among distinctiveness score and top-5 accuracy on the GMDB dataset.** The Spearman rank correlation coefficient was 0.600 ($P$ = 0.005). The details of each syndrome can be found in Supplementary Table 7 using the syndrome ID shown in the figure. The y-axis shows the average top-5 accuracy of the experiments over 100 iterations

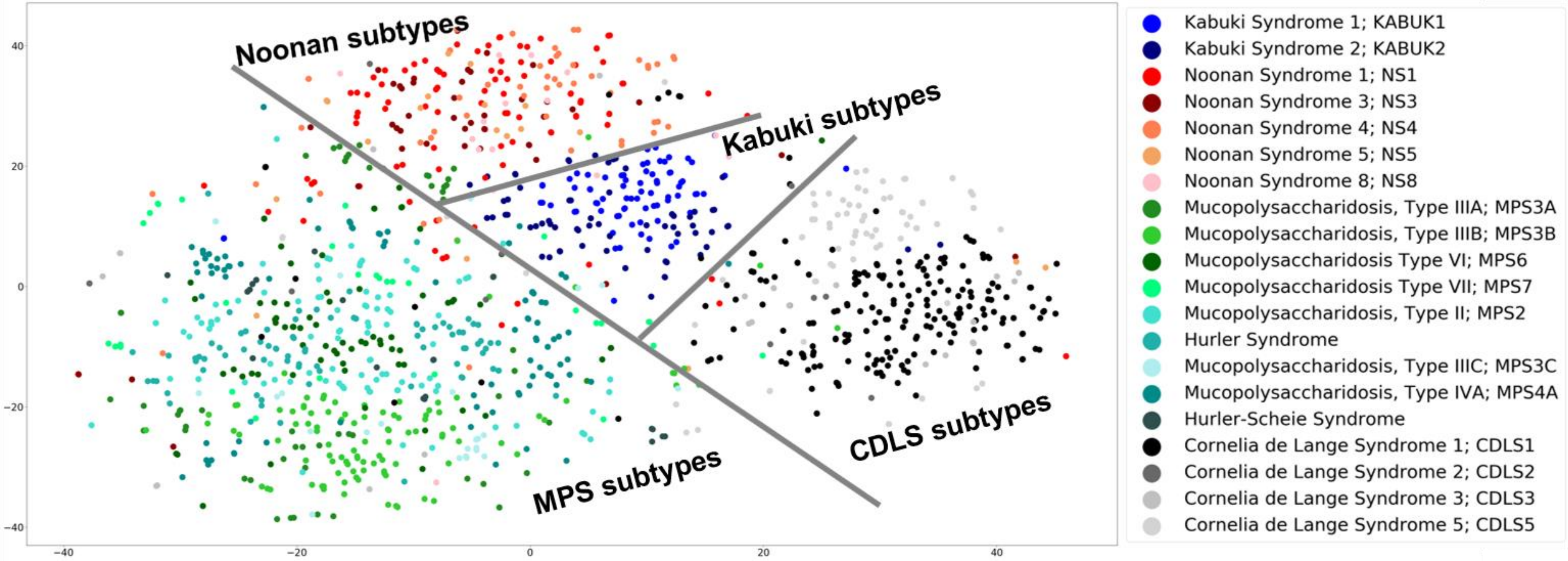


**Supplementary Figure 11: Hierarchical clustering of four phenotypic series, Kabuki syndrome, Noonan syndrome, mucopolysaccharidosis, and Cornelia de Lange syndrome, using a *t*-SNE projection of the Facial Phenotypic Descriptors.**

**a**

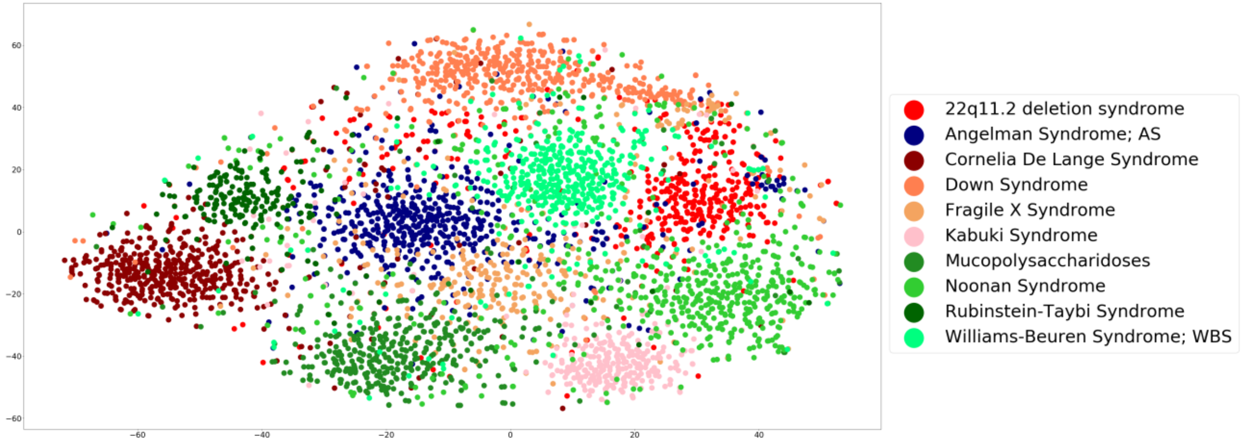


**b**

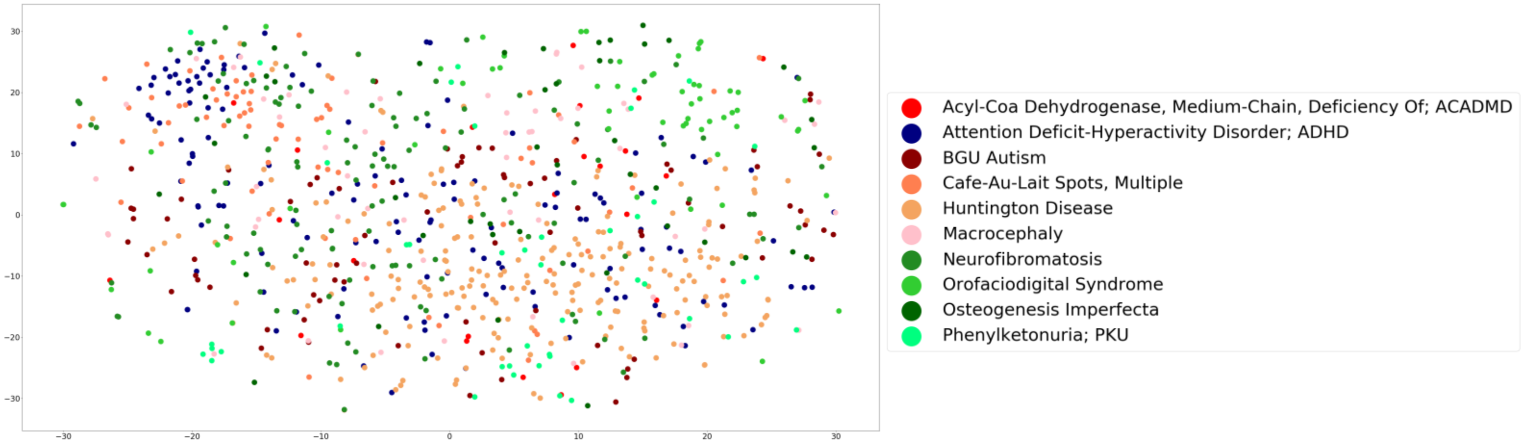


**Supplementary Figure 12: *t*-SNE visualization of Facial Phenotypic Descriptors of ten syndromes with (a) and ten syndromes without facial dysmorphism (b).**

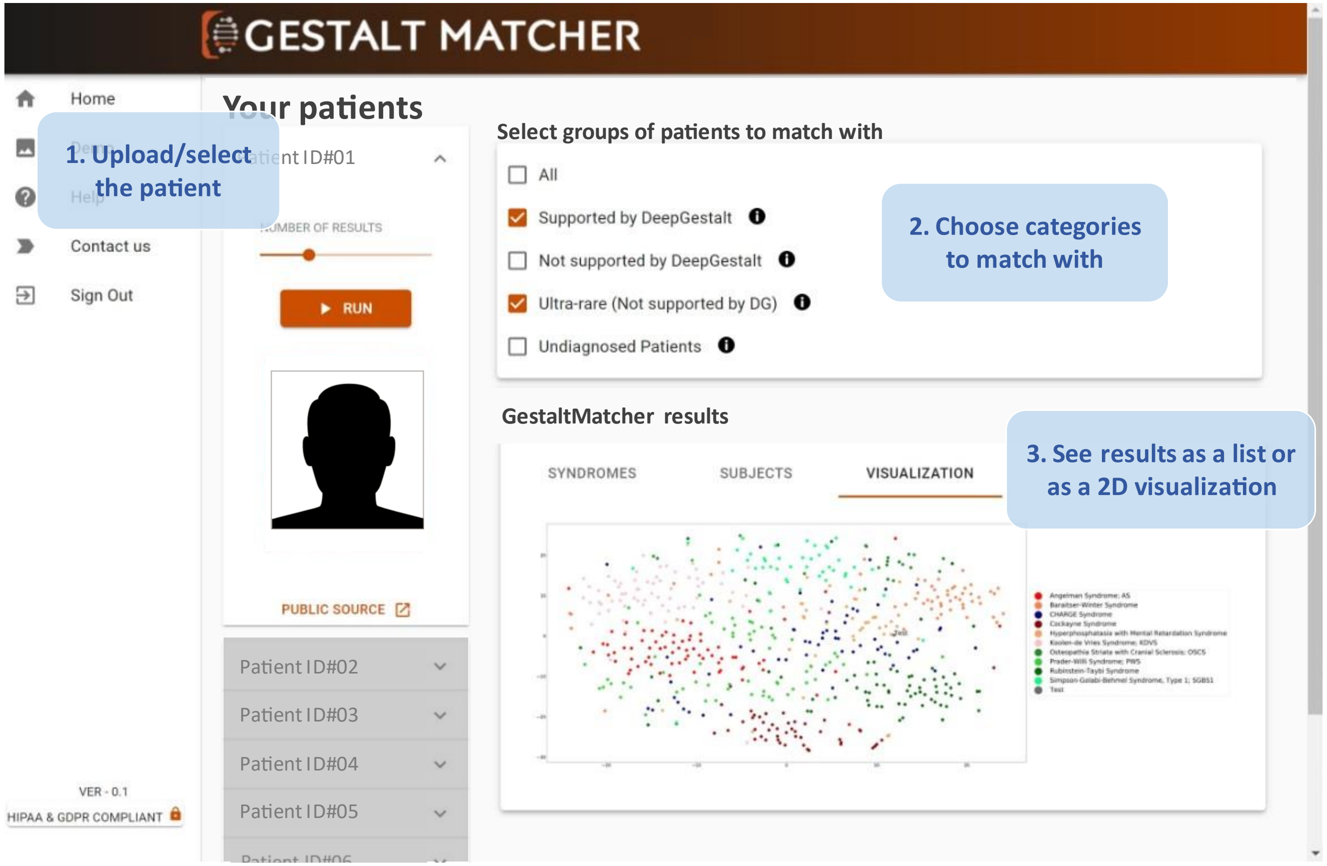


**Supplementary Figure 13: Screenshot of the GestaltMatcher web service.** Users can upload a patient photo to match the patients in the selected categories and can also visualize the clustering of patients by *t*-SNE. Access can be requested from gestaltmatcher.org.

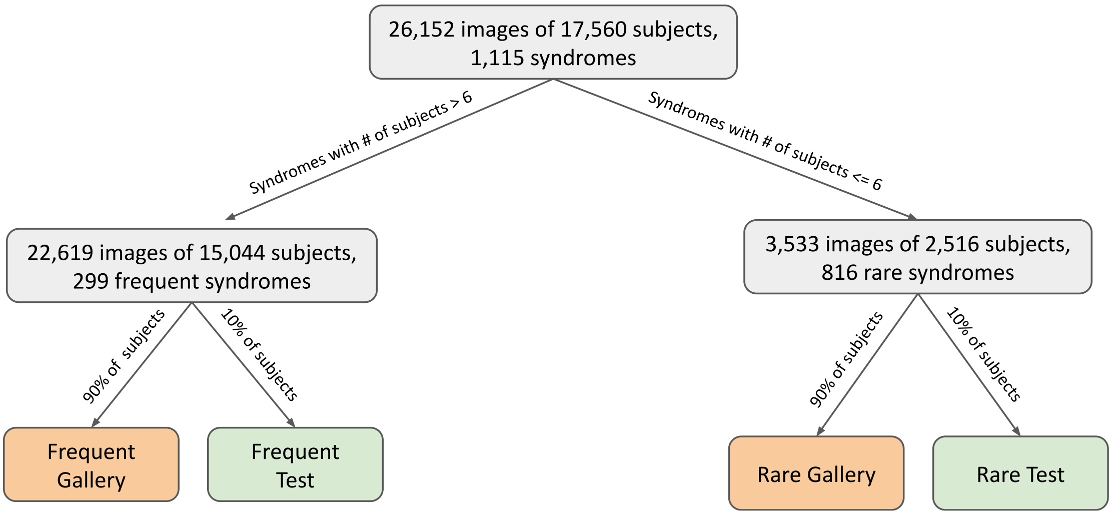


**Supplementary Figure 14: Overview of Face2Gene data categorization in GestaltMatcher.** The data were first divided by the number of subjects in each syndrome. Syndromes with more than six subjects were denoted frequent syndromes, and those with six or fewer as rare syndromes. Frequent syndromes were also recognized by DeepGestalt. Each category was further divided into a gallery and a test set. For each frequent syndrome, 90% of subjects were assigned to the gallery and used for model training; the remaining 10% of subjects were kept for validating the model training and were sampled in the test set. We performed 10-fold cross-validation on rare syndromes. In each syndrome, 90% of subjects were assigned to the gallery and 10% of subjects were assigned to the test set.

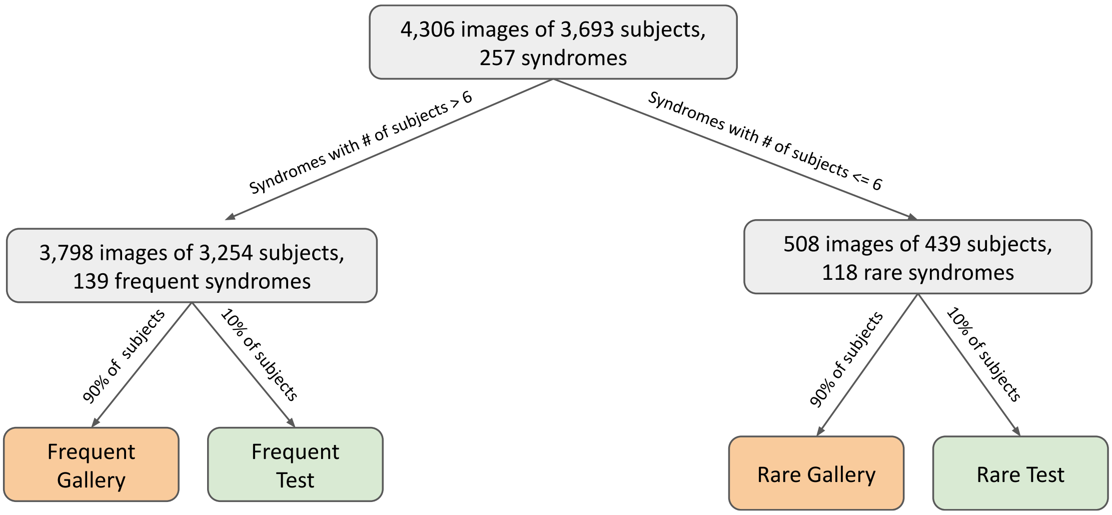


**Supplementary Figure 15: Overview of GMDB data categorization.** GMDB data were separated in the same way as Face2Gene data (see Supplementary Figure 14).

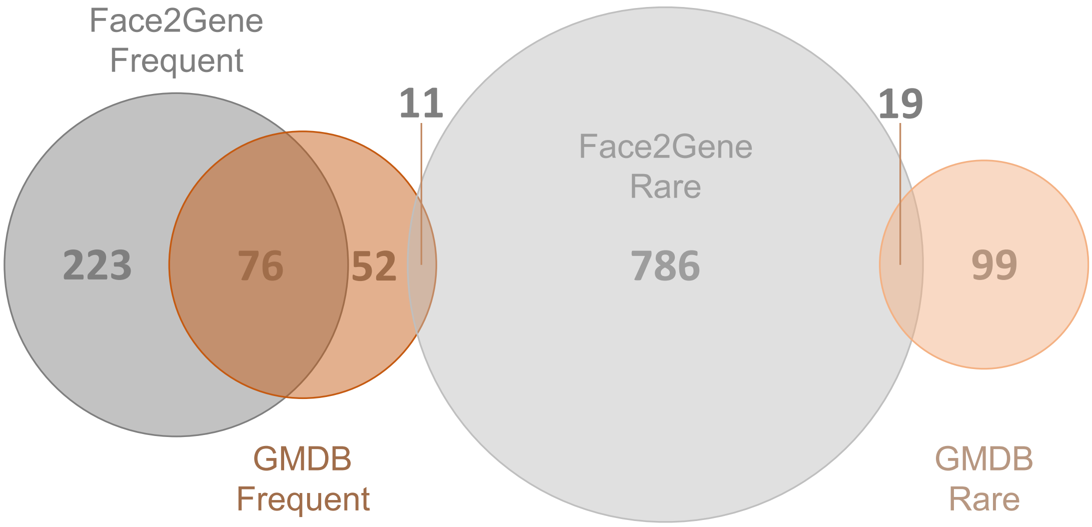


**Supplementary Figure 16: Venn diagram of numbers of syndromes in the Face2Gene and GMDB datasets.**

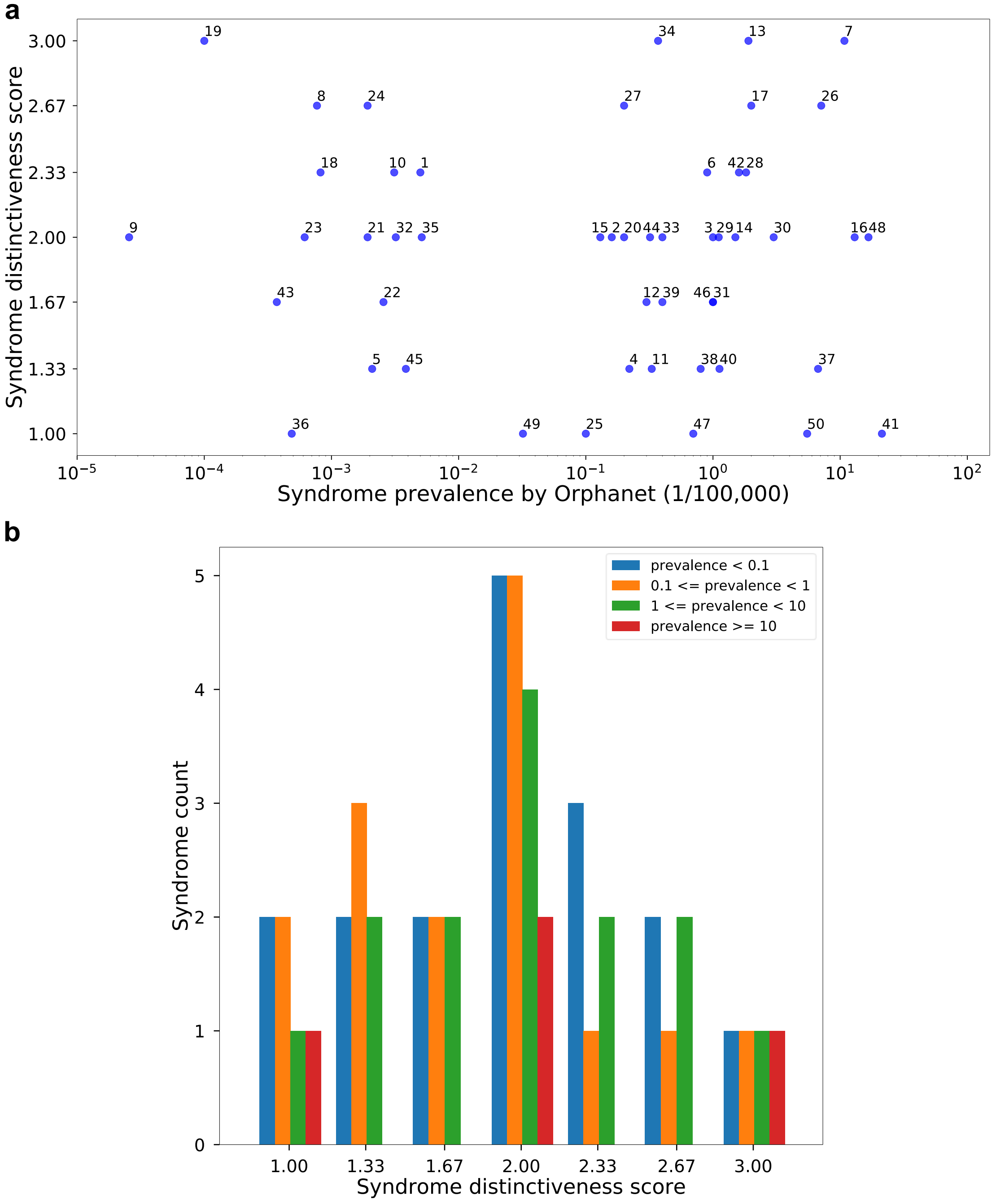


**Supplementary Figure 17: Distinctiveness scores and syndrome prevalence distribution of 50 selected syndromes. a**, Scatter plot of the 50 selected syndromes. **b**, Histogram of syndromes in each gestalt grade group. Prevalence is per 100,000 population.

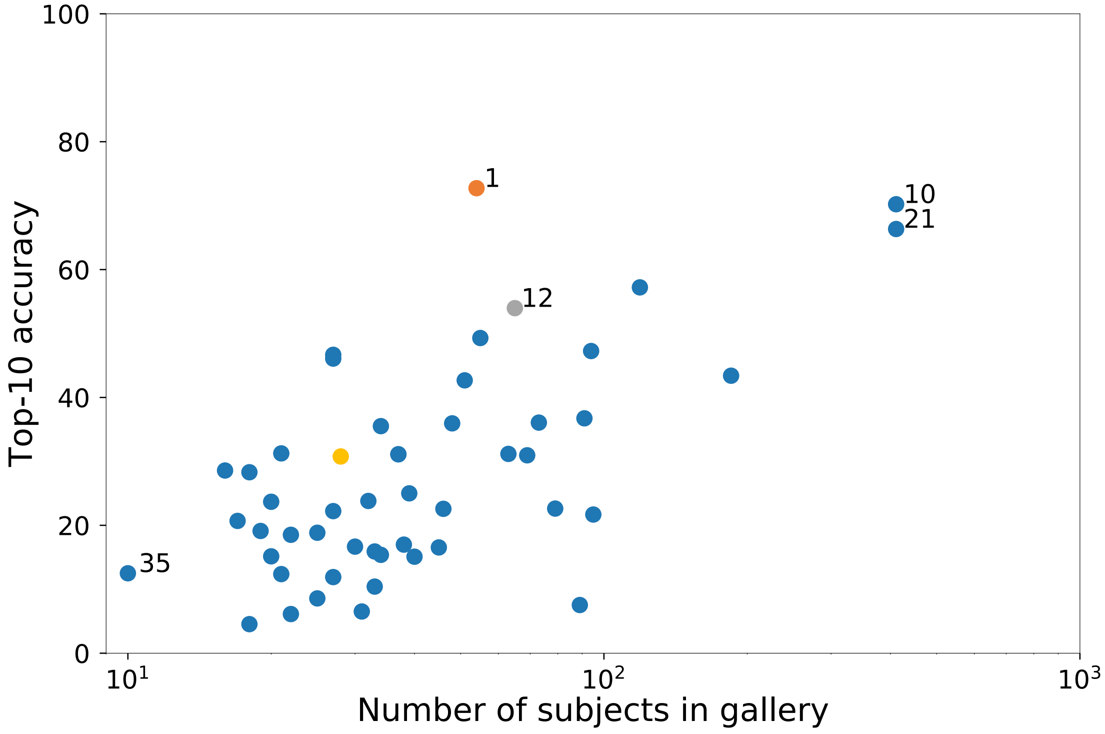


**Supplementary Figure 18: Correlation among the number of subjects in the gallery and top-10 accuracy on the Face2Gene dataset.** The Spearman rank correlation coefficient was 0.531 ($P < 0.001$). The details of each syndrome can be found in Supplementary Table 6 using the syndrome ID shown in the figure. The color dots are the same syndromes colored in Figure 5. The y-axis shows the average top-10 accuracy of the experiments over 10 random splits.

**Supplementary Table 1: Performance comparison of the Enc-F2G, Enc-GMDB, and Enc-healthy with the GMDB frequent and rare test sets.**

| Test set | Model | Gallery | | Test Images | Null top-1 accuracy | Top-1 | Top-5 | Top-10 | Top-30 |
|---|---|---|---|---|---|---|---|---|---|
| | | Images | Syndromes | | | | | | |
| GMDB-frequent | Enc-GMDB (softmax) | - | 139 | 360 | 0.71% | **29.98%** | **48.31%** | **66.30%** | **81.71%** |
| GMDB-frequent | Enc-GMDB | 3438 | 139 | 360 | 0.71% | 21.86% | 40.09% | 53.59% | 74.28% |
| GMDB-frequent | Enc-healthy | 3438 | 139 | 360 | 0.71% | 17.04% | 33.26% | 44.03% | 63.46% |
| GMDB-rare | Enc-F2G | 369.2 | 118 | 138.8 | 0.84% | 17.21% | **39.86%** | **49.30%** | 71.31% |
| GMDB-rare | Enc-GMDB | 369.2 | 118 | 138.8 | 0.84% | **18.51%** | 37.50% | 47.36% | **71.93%** |
| GMDB-rare | Enc-healthy | 369.2 | 118 | 138.8 | 0.84% | 14.85% | 30.53% | 40.43% | 61.65% |
| GMDB-frequent | Enc-GMDB | 3812[a] | 257[c] | 360 | 0.38% | **20.98%** | **38.25%** | **51.05%** | **71.37%** |
| GMDB-frequent | Enc-healthy | 3812[a] | 257[c] | 360 | 0.38% | 15.14% | 31.14% | 42.20% | 62.48% |
| GMDB-rare | Enc-F2G | 3807.2[b] | 257[c] | 138.8 | 0.38% | 8.43% | 17.65% | **23.39%** | **40.79%** |
| GMDB-rare | Enc-GMDB | 3807.2[b] | 257[c] | 138.8 | 0.38% | **8.47%** | **18.33%** | 23.19% | 37.62% |
| GMDB-rare | Enc-healthy | 3807.2[b] | 257[c] | 138.8 | 0.38% | 7.10% | 14.36% | 19.34% | 30.77% |

Enc-GMDB and Enc-F2G training were initiated with CASIA-WebFace and further fine-tuned on photos of patients in the Face2Gene frequent set. Enc-GMDB (softmax) used softmax to perform the prediction. For the top-1 to top-30 columns, the best performance in each set is boldfaced. The numbers of images and syndromes in the rare set are averaged over ten splits.

[a] Number of images in the frequent gallery and rare gallery.

[b] Average of ten splits in the frequent gallery and rare gallery.

[c] Number of syndromes in the frequent gallery and rare gallery.

**Supplementary Table 2: Comparison of GestaltMatcher and DeepGestalt on the LMD validation set.**

| Method | Gallery images | Supported Syndromes | Null top-1 accuracy | Top-1 | Top-5 | Top-10 | Top-30 |
|---|---|---|---|---|---|---|---|
| Enc-F2G (softmax) | - | 299 | 1.10% | **47.14%** | **71.19%** | **81.28%** | **88.34%** |
| Enc-F2G | 19,950 | 299 | 1.10% | 29.49% | 53.93% | 64.30% | 86.59% |
| Enc-F2G | **22,298** | **1,115** | 1.10% | 26.31% | 51.35% | 61.90% | 81.42% |

The results of 323 images from LMD, validated by GestaltMatcher (Enc-F2G) and DeepGestalt [Enc-F2G (softmax)]. We evaluated the GestaltMatcher approach on two different galleries, frequent (n = 299) and unified (n = 1,115). The best performance and the largest number of images and supported syndromes among the three conditions is boldfaced.

**Supplementary Table 3: Results of LMD validation analysis.** Each photo contains two results, from DeepGestalt and GestaltMatcher. The GestaltMatcher results were obtained with the frequent gallery.

| Index | Syndrome Name | OMIM ID | Links | DeepGestalt Rank | GestaltMatcher Rank |
|---|---|---|---|---|---|
| 1 | Alagille Syndrome | 118450 | https://app.face2gene.com/lmd/110/photo/345 | 52 | 55 |
| 2 | Nijmegen Breakage Syndrome; NBS | 251260 | https://app.face2gene.com/lmd/347/photo/4734 | 1 | 2 |
| 3 | Nijmegen Breakage Syndrome; NBS | 251260 | https://app.face2gene.com/lmd/347/photo/4730 | 2 | 1 |
| 4 | Craniodiaphyseal Dysplasia; CDD | 218300 | https://app.face2gene.com/lmd/373/photo/1088 | 2 | 1 |
| 5 | Noonan Syndrome | 163950 | https://app.face2gene.com/lmd/1946/photo/5976 | 39 | 10 |
| 6 | Noonan Syndrome | 163950 | https://app.face2gene.com/lmd/1946/photo/5985 | 8 | 23 |
| 7 | Smith-Lemli-Opitz Syndrome; SLOS | 270400 | https://app.face2gene.com/lmd/1608/photo/4893 | 5 | 32 |
| 8 | Kabuki Syndrome | 147920 | https://app.face2gene.com/lmd/893/photo/2567 | 1 | 1 |
| 9 | Focal Facial Dermal Dysplasia 3, Setleis Type; FFDD3 | 227260 | https://app.face2gene.com/lmd/1570/photo/4760 | 2 | 46 |
| 10 | Burn-Mckeown Syndrome; BMKS | 608572 | https://app.face2gene.com/lmd/2907/photo/6717 | 1 | 6 |
| 11 | Burn-Mckeown Syndrome; BMKS | 608572 | https://app.face2gene.com/lmd/2907/photo/6716 | 1 | 35 |
| 12 | Burn-Mckeown Syndrome; BMKS | 608572 | https://app.face2gene.com/lmd/2907/photo/6719 | 49 | 176 |
| 13 | Burn-Mckeown Syndrome; BMKS | 608572 | https://app.face2gene.com/lmd/2907/photo/6720 | 2 | 38 |
| 14 | Meier-Gorlin Syndrome 1; MGORS1 | 224690 | https://app.face2gene.com/lmd/808/photo/2269 | 5 | 16 |
| 15 | Williams-Beuren Syndrome; WBS | 194050 | https://app.face2gene.com/lmd/1829/photo/5615 | 1 | 1 |
| 16 | Trichorhinophalangeal Syndrome, Type II; TRPS2 | 150230 | https://app.face2gene.com/lmd/971/photo/2778 | 1 | 1 |
| 17 | Acrocallosal Syndrome; ACLS | 200990 | https://app.face2gene.com/lmd/23/photo/80 | 1 | 4 |
| 18 | Cohen Syndrome; COH1 | 216550 | https://app.face2gene.com/lmd/333/photo/1003 | 1 | 1 |
| 19 | Schwartz-Jampel Syndrome, Type 1; SJS1 | 255800 | https://app.face2gene.com/lmd/1548/photo/4685 | 26 | 21 |
| 20 | Treacher Collins Syndrome 1; TCS1 | 154500 | https://app.face2gene.com/lmd/1720/photo/5245 | 2 | 1 |
| 21 | Trichorhinophalangeal Syndrome | 190350 | https://app.face2gene.com/lmd/1730/photo/5290 | 1 | 1 |
| 22 | Velocardiofacial Syndrome | 192430 | https://app.face2gene.com/lmd/1762/photo/8285 | 2 | 2 |
| 23 | Smith-Lemli-Opitz Syndrome; SLOS | 270400 | https://app.face2gene.com/lmd/1608/photo/8342 | 1 | 1 |
| 24 | Marfan Syndrome; MFS | 154700 | https://app.face2gene.com/lmd/1057/photo/8255 | 6 | 12 |
| 25 | Williams-Beuren Syndrome; WBS | 194050 | https://app.face2gene.com/lmd/1829/photo/5603 | 1 | 1 |
| 26 | Mowat-Wilson Syndrome; MOWS | 235730 | https://app.face2gene.com/lmd/4211/photo/13183 | 1 | 2 |
| 27 | Mowat-Wilson Syndrome; MOWS | 235730 | https://app.face2gene.com/lmd/4211/photo/13743 | 1 | 1 |
| 28 | Mowat-Wilson Syndrome; MOWS | 235730 | https://app.face2gene.com/lmd/4211/photo/13744 | 1 | 1 |
| 29 | 3MC Syndrome 3; 3MC3 | 248340 | https://app.face2gene.com/lmd/1048/photo/14730 | 4 | 5 |

| | | | | | |
|---|---|---|---|---|---|
| 30 | Sotos Syndrome 2; SOTOS2 | 614753 | https://app.face2gene.com/lmd/1617/photo/15152 | 1 | 1 |
| 31 | Treacher Collins Syndrome 1; TCS1 | 154500 | https://app.face2gene.com/lmd/1720/photo/5227 | 2 | 4 |
| 32 | Coffin-Lowry Syndrome; CLS | 303600 | https://app.face2gene.com/lmd/328/photo/958 | 1 | 3 |
| 33 | Phelan-Mcdermid Syndrome; PHMDS | 606232 | https://app.face2gene.com/lmd/4459/photo/16283 | 4 | 36 |
| 34 | Donnai-Barrow Syndrome | 222448 | https://app.face2gene.com/lmd/3473/photo/16950 | 107 | 221 |
| 35 | Floating-Harbor Syndrome; FLHS | 136140 | https://app.face2gene.com/lmd/596/photo/17148 | 61 | 114 |
| 36 | Pitt-Hopkins Syndrome; PTHS | 610954 | https://app.face2gene.com/lmd/8892/photo/17151 | 1 | 1 |
| 37 | Marshall-Smith Syndrome; MRSHSS | 602535 | https://app.face2gene.com/lmd/7513/photo/17204 | 10 | 27 |
| 38 | 3MC Syndrome 3; 3MC3 | 248340 | https://app.face2gene.com/lmd/1048/photo/17445 | 70 | 194 |
| 39 | Williams-Beuren Syndrome; WBS | 194050 | https://app.face2gene.com/lmd/1829/photo/5608 | 1 | 2 |
| 40 | Waardenburg Syndrome, Type 1; WS1 | 193500 | https://app.face2gene.com/lmd/1781/photo/5438 | 20 | 57 |
| 41 | Nasopalpebral lipoma-coloboma syndrome; NPLCS | 167730 | https://app.face2gene.com/lmd/1208/photo/8174 | 1 | 1 |
| 42 | Sotos Syndrome 2; SOTOS2 | 614753 | https://app.face2gene.com/lmd/1617/photo/4945 | 1 | 2 |
| 43 | Pitt-Hopkins Syndrome; PTHS | 610954 | https://app.face2gene.com/lmd/8892/photo/17999 | 1 | 1 |
| 44 | Kabuki Syndrome | 147920 | https://app.face2gene.com/lmd/893/photo/18077 | 1 | 1 |
| 45 | Waardenburg Syndrome, Type 1; WS1 | 193500 | https://app.face2gene.com/lmd/1781/photo/8786 | 1 | 3 |
| 46 | Waardenburg Syndrome, Type 1; WS1 | 193500 | https://app.face2gene.com/lmd/1781/photo/8794 | 5 | 47 |
| 47 | Smith-Magenis Syndrome; SMS | 182290 | https://app.face2gene.com/lmd/3562/photo/9086 | 3 | 13 |
| 48 | Holoprosencephaly | 236100 | https://app.face2gene.com/lmd/787/photo/9105 | 12 | 29 |
| 49 | Seckel Syndrome | 210600 | https://app.face2gene.com/lmd/739/photo/9429 | 3 | 1 |
| 50 | Rothmund-Thomson Syndrome; RTS | 268400 | https://app.face2gene.com/lmd/1487/photo/9782 | 49 | 81 |
| 51 | Aarskog-Scott Syndrome; AAS | 305400 | https://app.face2gene.com/lmd/2/photo/9962 | 1 | 1 |
| 52 | Coffin-Siris Syndrome1; CSS1 | 135900 | https://app.face2gene.com/lmd/329/photo/18535 | 62 | 129 |
| 53 | Bohring-Opitz Syndrome; BOPS | 605039 | https://app.face2gene.com/lmd/4337/photo/18830 | 1 | 8 |
| 54 | SHORT syndrome | 269880 | https://app.face2gene.com/lmd/1574/photo/4787 | 2 | 1 |
| 55 | Schwartz-Jampel Syndrome, Type 1; SJS1 | 255800 | https://app.face2gene.com/lmd/1548/photo/4686 | 33 | 9 |
| 56 | Hajdu-Cheney Syndrome; HJCYS | 102500 | https://app.face2gene.com/lmd/722/photo/2067 | 10 | 40 |
| 57 | Coffin-Lowry Syndrome; CLS | 303600 | https://app.face2gene.com/lmd/328/photo/967 | 2 | 3 |
| 58 | Baraitser-Winter Syndrome 1; BRWS1 | 243310 | https://app.face2gene.com/lmd/149/photo/430 | 2 | 2 |
| 59 | Beckwith-Wiedemann Syndrome; BWS | 130650 | https://app.face2gene.com/lmd/173/photo/480 | 2 | 2 |
| 60 | Baraitser-Winter Syndrome 1; BRWS1 | 243310 | https://app.face2gene.com/lmd/149/photo/432 | 2 | 2 |
| 61 | Angelman Syndrome; AS | 105830 | https://app.face2gene.com/lmd/81/photo/279 | 1 | 1 |
| 62 | Angelman Syndrome; AS | 105830 | https://app.face2gene.com/lmd/81/photo/268 | 1 | 1 |
| 63 | Angelman Syndrome; AS | 105830 | https://app.face2gene.com/lmd/7322/photo/276 | 1 | 2 |
| 64 | Sotos Syndrome 2; SOTOS2 | 614753 | https://app.face2gene.com/lmd/1617/photo/4944 | 1 | 1 |
| 65 | Cockayne Syndrome | 216400 | https://app.face2gene.com/lmd/326/photo/946 | 2 | 2 |
| 66 | Fetal Alcohol Syndrome; FAS | - | https://app.face2gene.com/lmd/571/photo/1612 | 2 | 9 |

| | | | | | |
|---|---|---|---|---|---|
| 67 | Ectodermal Dysplasia 1, Hypohidrotic, X-Linked; XHED | 305100 | https://app.face2gene.com/lmd/508/photo/1407 | 1 | 1 |
| 68 | Coffin-Lowry Syndrome; CLS | 303600 | https://app.face2gene.com/lmd/328/photo/960 | 7 | 8 |
| 69 | Mucolipidosis II Alpha/beta | 252500 | https://app.face2gene.com/lmd/838/photo/2386 | 4 | 27 |
| 70 | Beckwith-Wiedemann Syndrome; BWS | 130650 | https://app.face2gene.com/lmd/173/photo/484 | 18 | 22 |
| 71 | Craniodiaphyseal Dysplasia; CDD | 218300 | https://app.face2gene.com/lmd/373/photo/1093 | 7 | 1 |
| 72 | Cockayne Syndrome | 216400 | https://app.face2gene.com/lmd/326/photo/948 | 13 | 29 |
| 73 | Floating-Harbor Syndrome; FLHS | 136140 | https://app.face2gene.com/lmd/596/photo/1761 | 1 | 1 |
| 74 | Rubinstein-Taybi Syndrome 1; RSTS1 | 180849 | https://app.face2gene.com/lmd/1490/photo/4474 | 3 | 2 |
| 75 | Coffin-Lowry Syndrome; CLS | 303600 | https://app.face2gene.com/lmd/328/photo/970 | 1 | 2 |
| 76 | Coffin-Lowry Syndrome; CLS | 303600 | https://app.face2gene.com/lmd/328/photo/971 | 1 | 2 |
| 77 | Apert Syndrome | 101200 | https://app.face2gene.com/lmd/99/photo/326 | 1 | 1 |
| 78 | Branchiooculofacial Syndrome; BOFS | 113620 | https://app.face2gene.com/lmd/633/photo/8345 | 2 | 7 |
| 79 | Smith-Lemli-Opitz Syndrome; SLOS | 270400 | https://app.face2gene.com/lmd/1608/photo/12105 | 1 | 2 |
| 80 | Smith-Lemli-Opitz Syndrome; SLOS | 270400 | https://app.face2gene.com/lmd/1608/photo/12105 | 31 | 6 |
| 81 | Coffin-Siris Syndrome1; CSS1 | 135900 | https://app.face2gene.com/lmd/329/photo/12535 | 7 | 3 |
| 82 | Coffin-Siris Syndrome1; CSS1 | 135900 | https://app.face2gene.com/lmd/329/photo/12535 | 11 | 2 |
| 83 | Kabuki Syndrome | 147920 | https://app.face2gene.com/lmd/893/photo/13114 | 1 | 1 |
| 84 | Kabuki Syndrome | 147920 | https://app.face2gene.com/lmd/893/photo/13116 | 1 | 1 |
| 85 | Mowat-Wilson Syndrome; MOWS | 235730 | https://app.face2gene.com/lmd/4211/photo/13734 | 1 | 1 |
| 86 | Mowat-Wilson Syndrome; MOWS | 235730 | https://app.face2gene.com/lmd/4211/photo/13736 | 84 | 111 |
| 87 | Mowat-Wilson Syndrome; MOWS | 235730 | https://app.face2gene.com/lmd/4211/photo/13742 | 1 | 1 |
| 88 | Borjeson-Forssman-Lehmann Syndrome; BFLS | 301900 | https://app.face2gene.com/lmd/216/photo/14759 | 47 | 27 |
| 89 | Smith-Magenis Syndrome; SMS | 182290 | https://app.face2gene.com/lmd/3562/photo/14969 | 3 | 14 |
| 90 | Aarskog-Scott Syndrome; AAS | 305400 | https://app.face2gene.com/lmd/2/photo/15758 | 4 | 9 |
| 91 | Aarskog-Scott Syndrome; AAS | 305400 | https://app.face2gene.com/lmd/2/photo/15758 | 1 | 7 |
| 92 | Opitz GBBB Syndrome, Type II; GBBB2 | 145410 | https://app.face2gene.com/lmd/640/photo/15940 | 2 | 42 |
| 93 | Silver-Russell Syndrome; SRS | 180860 | https://app.face2gene.com/lmd/1495/photo/16387 | 1 | 8 |
| 94 | Phelan-Mcdermid Syndrome; PHMDS | 606232 | https://app.face2gene.com/lmd/4459/photo/16512 | 1 | 20 |
| 95 | Sotos Syndrome 2; SOTOS2 | 614753 | https://app.face2gene.com/lmd/1617/photo/17023 | 1 | 1 |
| 96 | Sotos Syndrome 2; SOTOS2 | 614753 | https://app.face2gene.com/lmd/1617/photo/17024 | 13 | 16 |
| 97 | CHARGE Syndrome | 214800 | https://app.face2gene.com/lmd/301/photo/17094 | 1 | 3 |
| 98 | Floating-Harbor Syndrome; FLHS | 136140 | https://app.face2gene.com/lmd/596/photo/17124 | 1 | 1 |
| 99 | Prader-Willi Syndrome; PWS | 176270 | https://app.face2gene.com/lmd/1388/photo/4095 | 1 | 1 |
| 100 | Craniometaphyseal Dysplasia | 123000 | https://app.face2gene.com/lmd/371/photo/7667 | 58 | 120 |
| 101 | Craniosynostosis, Adelaide Type; CRSA | 600593 | https://app.face2gene.com/lmd/3793/photo/7990 | 132 | 252 |
| 102 | Pitt-Hopkins Syndrome; PTHS | 610954 | https://app.face2gene.com/lmd/8892/photo/18000 | 1 | 1 |

| | | | | | |
|---|---|---|---|---|---|
| 103 | Baraitser-Winter Syndrome 1; BRWS1 | 243310 | https://app.face2gene.com/lmd/149/photo/10154 | 6 | 1 |
| 104 | Baraitser-Winter Syndrome 1; BRWS1 | 243310 | https://app.face2gene.com/lmd/149/photo/10156 | 43 | 4 |
| 105 | Baraitser-Winter Syndrome 1; BRWS1 | 243310 | https://app.face2gene.com/lmd/149/photo/10156 | 85 | 3 |
| 106 | 3MC Syndrome 3; 3MC3 | 248340 | https://app.face2gene.com/lmd/1048/photo/10305 | 2 | 8 |
| 107 | Potocki-Shaffer Syndrome | 601224 | https://app.face2gene.com/lmd/3929/photo/10549 | 9 | 36 |
| 108 | Smith-Magenis Syndrome; SMS | 182290 | https://app.face2gene.com/lmd/3562/photo/18744 | 1 | 2 |
| 109 | Smith-Magenis Syndrome; SMS | 182290 | https://app.face2gene.com/lmd/3562/photo/18745 | 3 | 4 |
| 110 | Smith-Magenis Syndrome; SMS | 182290 | https://app.face2gene.com/lmd/3562/photo/18747 | 1 | 4 |
| 111 | Smith-Magenis Syndrome; SMS | 182290 | https://app.face2gene.com/lmd/3562/photo/18749 | 1 | 1 |
| 112 | Angelman Syndrome; AS | 105830 | https://app.face2gene.com/lmd/81/photo/263 | 4 | 2 |
| 113 | Pierre Robin Sequence with Cleft Mandible and Limb Anomalies | 268305 | https://app.face2gene.com/lmd/2127/photo/6384 | 5 | 3 |
| 114 | Pierre Robin Sequence with Cleft Mandible and Limb Anomalies | 268305 | https://app.face2gene.com/lmd/2127/photo/6385 | 1 | 5 |
| 115 | Crouzon Syndrome | 123500 | https://app.face2gene.com/lmd/383/photo/1133 | 1 | 2 |
| 116 | Angelman Syndrome; AS | 105830 | https://app.face2gene.com/lmd/81/photo/265 | 50 | 4 |
| 117 | Beckwith-Wiedemann Syndrome; BWS | 130650 | https://app.face2gene.com/lmd/173/photo/41779 | 1 | 1 |
| 118 | Otopalatodigital Syndrome | 311300 | https://app.face2gene.com/lmd/1266/photo/41780 | 1 | 1 |
| 119 | Waardenburg Syndrome, Type 1; WS1 | 193500 | https://app.face2gene.com/lmd/1781/photo/41781 | 1 | 1 |
| 120 | Rubinstein-Taybi Syndrome | 180849 | https://app.face2gene.com/lmd/1490/photo/41785 | 1 | 2 |
| 121 | Treacher Collins Syndrome | 154500 | https://app.face2gene.com/lmd/1720/photo/41799 | 1 | 3 |
| 122 | Cornelia De Lange Syndrome | 122470 | https://app.face2gene.com/lmd/423/photo/41811 | 1 | 1 |
| 123 | Cornelia De Lange Syndrome | 122470 | https://app.face2gene.com/lmd/423/photo/41812 | 1 | 1 |
| 124 | Cornelia De Lange Syndrome | 122470 | https://app.face2gene.com/lmd/423/photo/41813 | 1 | 1 |
| 125 | Cornelia De Lange Syndrome | 122470 | https://app.face2gene.com/lmd/423/photo/41814 | 1 | 1 |
| 126 | Sotos Syndrome | 117550 | https://app.face2gene.com/lmd/1617/photo/41817 | 1 | 4 |
| 127 | Ectodermal Dysplasia 1, Hypohidrotic, X-Linked; XHED | 305100 | https://app.face2gene.com/lmd/508/photo/41823 | 1 | 13 |
| 128 | Down Syndrome | 190685 | https://app.face2gene.com/lmd/6905/photo/41830 | 1 | 1 |
| 129 | Wolf-Hirschhorn Syndrome; WHS | 194190 | https://app.face2gene.com/lmd/4706/photo/41832 | 1 | 1 |
| 130 | Williams-Beuren Syndrome; WBS | 194050 | https://app.face2gene.com/lmd/1829/photo/41840 | 11 | 2 |
| 131 | Hutchinson-Gilford Progeria Syndrome; HGPS | 176670 | https://app.face2gene.com/lmd/1396/photo/41849 | 1 | 1 |
| 132 | Cri-Du-Chat Syndrome | 123450 | https://app.face2gene.com/lmd/13213/photo/41863 | 1 | 4 |
| 133 | Hurler Syndrome | 607014 | https://app.face2gene.com/lmd/13858/photo/41873 | 4 | 5 |
| 134 | Hurler Syndrome | 607014 | https://app.face2gene.com/lmd/13858/photo/41874 | 1 | 1 |
| 135 | Mucopolysaccharidosis, Type II; MPS2 | 309900 | https://app.face2gene.com/lmd/801/photo/41875 | 3 | 1 |
| 136 | Mucopolysaccharidosis, Type II; MPS2 | 309900 | https://app.face2gene.com/lmd/801/photo/41876 | 1 | 1 |
| 137 | Mucopolysaccharidosis, Type II; MPS2 | 309900 | https://app.face2gene.com/lmd/801/photo/41878 | 2 | 1 |

| | | | | | |
|---|---|---|---|---|---|
| 138 | Mucopolysaccharidosis, Type II; MPS2 | 309900 | https://app.face2gene.com/lmd/801/photo/41879 | 1 | 1 |
| 139 | Mucolipidosis II Alpha/beta | 252500 | https://app.face2gene.com/lmd/838/photo/41883 | 1 | 3 |
| 140 | Cleidocranial Dysplasia; CCD | 119600 | https://app.face2gene.com/lmd/13331/photo/41890 | 2 | 1 |
| 141 | Cleidocranial Dysplasia; CCD | 119600 | https://app.face2gene.com/lmd/13331/photo/41891 | 2 | 25 |
| 142 | Seckel Syndrome | 210600 | https://app.face2gene.com/lmd/739/photo/41904 | 6 | 23 |
| 143 | Silver-Russell Syndrome; SRS | 180860 | https://app.face2gene.com/lmd/1495/photo/41913 | 2 | 3 |
| 144 | Crouzon Syndrome | 123500 | https://app.face2gene.com/lmd/383/photo/41914 | 5 | 1 |
| 145 | Williams-Beuren Syndrome; WBS | 194050 | https://app.face2gene.com/lmd/1829/photo/41923 | 1 | 1 |
| 146 | Williams-Beuren Syndrome; WBS | 194050 | https://app.face2gene.com/lmd/1829/photo/41924 | 1 | 2 |
| 147 | Williams-Beuren Syndrome; WBS | 194050 | https://app.face2gene.com/lmd/1829/photo/41925 | 1 | 3 |
| 148 | Williams-Beuren Syndrome; WBS | 194050 | https://app.face2gene.com/lmd/1829/photo/41926 | 1 | 1 |
| 149 | Williams-Beuren Syndrome; WBS | 194050 | https://app.face2gene.com/lmd/1829/photo/41927 | 6 | 3 |
| 150 | Sotos Syndrome | 117550 | https://app.face2gene.com/lmd/1617/photo/41940 | 1 | 1 |
| 151 | Sotos Syndrome | 117550 | https://app.face2gene.com/lmd/1617/photo/41941 | 1 | 8 |
| 152 | Silver-Russell Syndrome; SRS | 180860 | https://app.face2gene.com/lmd/1495/photo/41945 | 1 | 1 |
| 153 | Hallermann-Streiff Syndrome; HSS | 234100 | https://app.face2gene.com/lmd/730/photo/41946 | 1 | 1 |
| 154 | Cleidocranial Dysplasia; CCD | 119600 | https://app.face2gene.com/lmd/13331/photo/41947 | 1 | 1 |
| 155 | Treacher Collins Syndrome | 154500 | https://app.face2gene.com/lmd/1720/photo/41956 | 3 | 5 |
| 156 | Treacher Collins Syndrome | 154500 | https://app.face2gene.com/lmd/1720/photo/41958 | 5 | 13 |
| 157 | Williams-Beuren Syndrome; WBS | 194050 | https://app.face2gene.com/lmd/1829/photo/41964 | 1 | 1 |
| 158 | Williams-Beuren Syndrome; WBS | 194050 | https://app.face2gene.com/lmd/1829/photo/41965 | 1 | 1 |
| 159 | Williams-Beuren Syndrome; WBS | 194050 | https://app.face2gene.com/lmd/1829/photo/41966 | 1 | 1 |
| 160 | Fragile X Mental Retardation Syndrome | 300624 | https://app.face2gene.com/lmd/13243/photo/41973 | 1 | 3 |
| 161 | Angelman Syndrome; AS | 105830 | https://app.face2gene.com/lmd/81/photo/41983 | 4 | 3 |
| 162 | Smith-Magenis Syndrome; SMS | 182290 | https://app.face2gene.com/lmd/3562/photo/15947 | 5 | 7 |
| 163 | Pierpont Syndrome; PRPTS | 602342 | https://app.face2gene.com/lmd/4198/photo/41638 | 4 | 4 |
| 164 | Pierpont Syndrome; PRPTS | 602342 | https://app.face2gene.com/lmd/4198/photo/41640 | 1 | 10 |
| 165 | SHORT syndrome | 269880 | https://app.face2gene.com/lmd/1574/photo/4780 | 1 | 11 |
| 166 | Kabuki Syndrome | 147920 | https://app.face2gene.com/lmd/893/photo/2566 | 1 | 1 |
| 167 | Kabuki Syndrome | 147920 | https://app.face2gene.com/lmd/893/photo/2560 | 1 | 1 |
| 168 | Kabuki Syndrome | 147920 | https://app.face2gene.com/lmd/893/photo/2562 | 38 | 40 |
| 169 | Otopalatodigital Syndrome | 311300 | https://app.face2gene.com/lmd/1266/photo/3719 | 1 | 1 |
| 170 | Coffin-Lowry Syndrome; CLS | 303600 | https://app.face2gene.com/lmd/328/photo/977 | 1 | 1 |
| 171 | Ohdo Syndrome, SBBYS Variant; SBBYSS | 603736 | https://app.face2gene.com/lmd/1255/photo/8343 | 2 | 9 |
| 172 | Rubinstein-Taybi Syndrome 1; RSTS1 | 180849 | https://app.face2gene.com/lmd/358/photo/11625 | 1 | 1 |
| 173 | Auriculocondylar Syndrome 1; ARCND1 | 602483 | https://app.face2gene.com/lmd/4192/photo/12354 | 2 | 56 |
| 174 | Coffin-Siris Syndrome1; CSS1 | 135900 | https://app.face2gene.com/lmd/329/photo/12530 | 6 | 22 |
| 175 | Urofacial Syndrome 1; UFS1 | 236730 | https://app.face2gene.com/lmd/1236/photo/13084 | 3 | 15 |

| | | | | | |
|---|---|---|---|---|---|
| 176 | 3MC Syndrome 3; 3MC3 | 248340 | https://app.face2gene.com/lmd/1048/photo/13477 | 7 | 9 |
| 177 | Smith-Lemli-Opitz Syndrome; SLOS | 270400 | https://app.face2gene.com/lmd/1608/photo/14313 | 11 | 32 |
| 178 | 3MC Syndrome 3; 3MC3 | 248340 | https://app.face2gene.com/lmd/1048/photo/14727 | 3 | 3 |
| 179 | Phelan-Mcdermid Syndrome; PHMDS | 606232 | https://app.face2gene.com/lmd/4459/photo/16282 | 12 | 12 |
| 180 | Lig4 Syndrome | 606593 | https://app.face2gene.com/lmd/4654/photo/17338 | 5 | 5 |
| 181 | Craniometaphyseal Dysplasia | 123000 | https://app.face2gene.com/lmd/371/photo/1078 | 1 | 2 |
| 182 | Angelman Syndrome; AS | 105830 | https://app.face2gene.com/lmd/81/photo/284 | 1 | 1 |
| 183 | Angelman Syndrome; AS | 105830 | https://app.face2gene.com/lmd/81/photo/274 | 1 | 2 |
| 184 | Angelman Syndrome; AS | 105830 | https://app.face2gene.com/lmd/81/photo/264 | 2 | 1 |
| 185 | Craniofrontonasal Syndrome; CFNS | 304110 | https://app.face2gene.com/lmd/376/photo/1113 | 1 | 1 |
| 186 | Angelman Syndrome; AS | 105830 | https://app.face2gene.com/lmd/81/photo/269 | 1 | 1 |
| 187 | Floating-Harbor Syndrome; FLHS | 136140 | https://app.face2gene.com/lmd/596/photo/1756 | 1 | 1 |
| 188 | Rubinstein-Taybi Syndrome 1; RSTS1 | 180849 | https://app.face2gene.com/lmd/1490/photo/4488 | 1 | 7 |
| 189 | Coffin-Siris Syndrome1; CSS1 | 135900 | https://app.face2gene.com/lmd/329/photo/981 | 1 | 1 |
| 190 | Rubinstein-Taybi Syndrome 1; RSTS1 | 180849 | https://app.face2gene.com/lmd/1490/photo/4489 | 1 | 2 |
| 191 | Noonan Syndrome | 163950 | https://app.face2gene.com/lmd/1229/photo/3585 | 3 | 1 |
| 192 | Floating-Harbor Syndrome; FLHS | 136140 | https://app.face2gene.com/lmd/596/photo/8231 | 3 | 7 |
| 193 | Smith-Lemli-Opitz Syndrome; SLOS | 270400 | https://app.face2gene.com/lmd/1608/photo/12105 | 1 | 17 |
| 194 | Kabuki Syndrome | 147920 | https://app.face2gene.com/lmd/893/photo/13113 | 1 | 2 |
| 195 | Kabuki Syndrome | 147920 | https://app.face2gene.com/lmd/893/photo/13113 | 1 | 2 |
| 196 | Sotos Syndrome 2; SOTOS2 | 614753 | https://app.face2gene.com/lmd/1617/photo/15150 | 1 | 4 |
| 197 | Phelan-Mcdermid Syndrome; PHMDS | 606232 | https://app.face2gene.com/lmd/4459/photo/16512 | 1 | 2 |
| 198 | Phelan-Mcdermid Syndrome; PHMDS | 606232 | https://app.face2gene.com/lmd/4459/photo/16512 | 1 | 10 |
| 199 | Sotos Syndrome 2; SOTOS2 | 614753 | https://app.face2gene.com/lmd/1617/photo/17025 | 1 | 1 |
| 200 | Potocki-Shaffer Syndrome | 601224 | https://app.face2gene.com/lmd/3929/photo/10549 | 6 | 40 |
| 201 | Smith-Magenis Syndrome; SMS | 182290 | https://app.face2gene.com/lmd/3562/photo/18745 | 1 | 7 |
| 202 | Smith-Magenis Syndrome; SMS | 182290 | https://app.face2gene.com/lmd/3562/photo/18745 | 4 | 2 |
| 203 | Birk-Barel Mental Retardation Dysmorphism Syndrome | 612292 | https://app.face2gene.com/lmd/5569/photo/19317 | 23 | 128 |
| 204 | Blepharophimosis, Ptosis, and Epicanthus Inversus; BPES | 110100 | https://app.face2gene.com/lmd/202/photo/580 | 61 | 2 |
| 205 | Focal Dermal Hypoplasia; FDH | 305600 | https://app.face2gene.com/lmd/13389/photo/41764 | 1 | 1 |
| 206 | Williams-Beuren Syndrome; WBS | 194050 | https://app.face2gene.com/lmd/1829/photo/41783 | 1 | 1 |
| 207 | Williams-Beuren Syndrome; WBS | 194050 | https://app.face2gene.com/lmd/1829/photo/41784 | 1 | 4 |
| 208 | Moebius Syndrome; MBS | 157900 | https://app.face2gene.com/lmd/1150/photo/41804 | 1 | 1 |
| 209 | Moebius Syndrome; MBS | 157900 | https://app.face2gene.com/lmd/1150/photo/41805 | 1 | 4 |
| 210 | Cornelia De Lange Syndrome | 122470 | https://app.face2gene.com/lmd/423/photo/41808 | 3 | 1 |
| 211 | Cornelia De Lange Syndrome | 122470 | https://app.face2gene.com/lmd/423/photo/41810 | 1 | 1 |

| | | | | | |
|---|---|---|---|---|---|
| 212 | Sotos Syndrome | 117550 | https://app.face2gene.com/lmd/1617/photo/41818 | 1 | 1 |
| 213 | Moebius Syndrome; MBS | 157900 | https://app.face2gene.com/lmd/1150/photo/41821 | 1 | 6 |
| 214 | Focal Dermal Hypoplasia; FDH | 305600 | https://app.face2gene.com/lmd/13389/photo/41822 | 14 | 8 |
| 215 | Mucopolysaccharidosis, Type II; MPS2 | 309900 | https://app.face2gene.com/lmd/801/photo/41824 | 6 | 2 |
| 216 | Prader-Willi Syndrome; PWS | 176270 | https://app.face2gene.com/lmd/1388/photo/41837 | 1 | 1 |
| 217 | Crouzon Syndrome | 123500 | https://app.face2gene.com/lmd/383/photo/41842 | 1 | 1 |
| 218 | Rubinstein-Taybi Syndrome | 180849 | https://app.face2gene.com/lmd/1490/photo/41844 | 1 | 1 |
| 219 | Hutchinson-Gilford Progeria Syndrome; HGPS | 176670 | https://app.face2gene.com/lmd/1396/photo/41845 | 1 | 1 |
| 220 | Hutchinson-Gilford Progeria Syndrome; HGPS | 176670 | https://app.face2gene.com/lmd/1396/photo/41846 | 1 | 2 |
| 221 | Hutchinson-Gilford Progeria Syndrome; HGPS | 176670 | https://app.face2gene.com/lmd/1396/photo/41847 | 1 | 2 |
| 222 | Hutchinson-Gilford Progeria Syndrome; HGPS | 176670 | https://app.face2gene.com/lmd/1396/photo/41848 | 1 | 1 |
| 223 | Hutchinson-Gilford Progeria Syndrome; HGPS | 176670 | https://app.face2gene.com/lmd/1396/photo/41850 | 4 | 1 |
| 224 | Noonan Syndrome | 163950 | https://app.face2gene.com/lmd/1229/photo/41851 | 1 | 13 |
| 225 | Down Syndrome | 190685 | https://app.face2gene.com/lmd/6905/photo/41853 | 1 | 11 |
| 226 | Down Syndrome | 190685 | https://app.face2gene.com/lmd/6905/photo/41854 | 1 | 3 |
| 227 | Rubinstein-Taybi Syndrome | 180849 | https://app.face2gene.com/lmd/1490/photo/41859 | 1 | 1 |
| 228 | Achondroplasia; ACH | 100800 | https://app.face2gene.com/lmd/17/photo/41871 | 4 | 13 |
| 229 | Hurler Syndrome | 607014 | https://app.face2gene.com/lmd/13858/photo/41872 | 15 | 9 |
| 230 | Cleidocranial Dysplasia; CCD | 119600 | https://app.face2gene.com/lmd/13331/photo/41899 | 3 | 11 |
| 231 | Silver-Russell Syndrome; SRS | 180860 | https://app.face2gene.com/lmd/1495/photo/41908 | 4 | 27 |
| 232 | Saethre-Chotzen Syndrome; SCS | 101400 | https://app.face2gene.com/lmd/1504/photo/41909 | 1 | 17 |
| 233 | Cleidocranial Dysplasia; CCD | 119600 | https://app.face2gene.com/lmd/13331/photo/41948 | 10 | 44 |
| 234 | Treacher Collins Syndrome | 154500 | https://app.face2gene.com/lmd/1720/photo/41951 | 1 | 4 |
| 235 | Treacher Collins Syndrome | 154500 | https://app.face2gene.com/lmd/1720/photo/41952 | 1 | 1 |
| 236 | Treacher Collins Syndrome | 154500 | https://app.face2gene.com/lmd/1720/photo/41954 | 1 | 2 |
| 237 | Treacher Collins Syndrome | 154500 | https://app.face2gene.com/lmd/1720/photo/41959 | 1 | 6 |
| 238 | Treacher Collins Syndrome | 154500 | https://app.face2gene.com/lmd/1720/photo/41960 | 1 | 1 |
| 239 | Fragile X Mental Retardation Syndrome | 300624 | https://app.face2gene.com/lmd/13243/photo/41982 | 3 | 3 |
| 240 | Pitt-Hopkins Syndrome; PTHS | 610954 | https://app.face2gene.com/lmd/8892/photo/18003 | 1 | 5 |
| 241 | Nijmegen Breakage Syndrome; NBS | 251260 | https://app.face2gene.com/lmd/347/photo/4723 | 1 | 1 |
| 242 | Nijmegen Breakage Syndrome; NBS | 251260 | https://app.face2gene.com/lmd/347/photo/4736 | 35 | 51 |
| 243 | Noonan Syndrome | 163950 | https://app.face2gene.com/lmd/1946/photo/5983 | 20 | 2 |
| 244 | Kabuki Syndrome | 147920 | https://app.face2gene.com/lmd/893/photo/2565 | 1 | 1 |
| 245 | Craniodiaphyseal Dysplasia; CDD | 218300 | https://app.face2gene.com/lmd/373/photo/1091 | 1 | 1 |
| 246 | Ectodermal Dysplasia 1, Hypohidrotic, X-Linked; XHED | 305100 | https://app.face2gene.com/lmd/508/photo/1400 | 1 | 1 |
| 247 | Mowat-Wilson Syndrome; MOWS | 235730 | https://app.face2gene.com/lmd/4211/photo/13735 | 1 | 1 |
| 248 | 3MC Syndrome 3; 3MC3 | 248340 | https://app.face2gene.com/lmd/1048/photo/13467 | 2 | 1 |

| | | | | | |
|---|---|---|---|---|---|
| 249 | Auriculocondylar Syndrome 1; ARCND1 | 602483 | https://app.face2gene.com/lmd/4192/photo/14125 | 5 | 131 |
| 250 | Nicolaides-Baraitser Syndrome; NCBRS | 601358 | https://app.face2gene.com/lmd/3394/photo/14255 | 1 | 2 |
| 251 | Smith-Magenis Syndrome; SMS | 182290 | https://app.face2gene.com/lmd/3562/photo/14454 | 2 | 2 |
| 252 | Nicolaides-Baraitser Syndrome; NCBRS | 601358 | https://app.face2gene.com/lmd/3394/photo/14700 | 1 | 16 |
| 253 | Noonan Syndrome | 163950 | https://app.face2gene.com/lmd/1229/photo/3581 | 1 | 3 |
| 254 | KBG Syndrome; KBGS | 148050 | https://app.face2gene.com/lmd/912/photo/15604 | 1 | 1 |
| 255 | Angelman Syndrome; AS | 105830 | https://app.face2gene.com/lmd/81/photo/272 | 3 | 3 |
| 256 | Aarskog-Scott Syndrome; AAS | 305400 | https://app.face2gene.com/lmd/2/photo/9965 | 2 | 26 |
| 257 | Beckwith-Wiedemann Syndrome; BWS | 130650 | https://app.face2gene.com/lmd/173/photo/481 | 1 | 2 |
| 258 | Craniofrontonasal Syndrome; CFNS | 304110 | https://app.face2gene.com/lmd/376/photo/1115 | 1 | 1 |
| 259 | Floating-Harbor Syndrome; FLHS | 136140 | https://app.face2gene.com/lmd/596/photo/1760 | 1 | 1 |
| 260 | Coffin-Lowry Syndrome; CLS | 303600 | https://app.face2gene.com/lmd/328/photo/974 | 6 | 6 |
| 261 | Sotos Syndrome 2; SOTOS2 | 614753 | https://app.face2gene.com/lmd/1617/photo/15146 | 1 | 1 |
| 262 | Smith-Magenis Syndrome; SMS | 182290 | https://app.face2gene.com/lmd/3562/photo/15950 | 2 | 5 |
| 263 | Phelan-Mcdermid Syndrome; PHMDS | 606232 | https://app.face2gene.com/lmd/4459/photo/16512 | 1 | 6 |
| 264 | Lateral Meningocele Syndrome; LMNS | 130720 | https://app.face2gene.com/lmd/983/photo/10028 | 44 | 11 |
| 265 | KBG Syndrome; KBGS | 148050 | https://app.face2gene.com/lmd/912/photo/10163 | 30 | 54 |
| 266 | Smith-Magenis Syndrome; SMS | 182290 | https://app.face2gene.com/lmd/3562/photo/18749 | 14 | 12 |
| 267 | Aarskog Syndrome | 305400 | https://app.face2gene.com/lmd/2/photo/41775 | 1 | 4 |
| 268 | Bardet-Biedl Syndrome | 209900 | https://app.face2gene.com/lmd/13278/photo/41796 | 1 | 5 |
| 269 | Cornelia De Lange Syndrome | 122470 | https://app.face2gene.com/lmd/423/photo/41807 | 1 | 1 |
| 270 | Cri-Du-Chat Syndrome | 123450 | https://app.face2gene.com/lmd/13213/photo/41826 | 1 | 2 |
| 271 | Crouzon Syndrome | 123500 | https://app.face2gene.com/lmd/383/photo/41828 | 1 | 2 |
| 272 | Prader-Willi Syndrome; PWS | 176270 | https://app.face2gene.com/lmd/1388/photo/41911 | 1 | 6 |
| 273 | Rubinstein-Taybi Syndrome | 180849 | https://app.face2gene.com/lmd/1490/photo/41858 | 1 | 1 |
| 274 | Rubinstein-Taybi Syndrome | 180849 | https://app.face2gene.com/lmd/1490/photo/41860 | 1 | 1 |
| 275 | Mucopolysaccharidosis, Type II; MPS2 | 309900 | https://app.face2gene.com/lmd/801/photo/41877 | 2 | 2 |
| 276 | Trichorhinophalangeal Syndrome | 190350 | https://app.face2gene.com/lmd/1730/photo/41886 | 1 | 4 |
| 277 | Trichorhinophalangeal Syndrome | 190350 | https://app.face2gene.com/lmd/1730/photo/41887 | 1 | 1 |
| 278 | Cleidocranial Dysplasia; CCD | 119600 | https://app.face2gene.com/lmd/13331/photo/41900 | 25 | 112 |
| 279 | Saethre-Chotzen Syndrome; SCS | 101400 | https://app.face2gene.com/lmd/1504/photo/41910 | 32 | 18 |
| 280 | Treacher Collins Syndrome | 154500 | https://app.face2gene.com/lmd/1720/photo/41961 | 1 | 1 |
| 281 | Frontometaphyseal Dysplasia; FMD | 305620 | https://app.face2gene.com/lmd/622/photo/41685 | 1 | 4 |
| 282 | Kabuki Syndrome | 147920 | https://app.face2gene.com/lmd/893/photo/2569 | 1 | 1 |
| 283 | Nijmegen Breakage Syndrome; NBS | 251260 | https://app.face2gene.com/lmd/347/photo/4728 | 6 | 15 |
| 284 | Coffin-Lowry Syndrome; CLS | 303600 | https://app.face2gene.com/lmd/328/photo/978 | 1 | 4 |
| 285 | Hutchinson-Gilford Progeria Syndrome; HGPS | 176670 | https://app.face2gene.com/lmd/1396/photo/4126 | 1 | 1 |
| 286 | Costello Syndrome; CSTLO | 218040 | https://app.face2gene.com/lmd/2014/photo/6166 | 1 | 1 |

| | | | | | |
|---|---|---|---|---|---|
| 287 | Kabuki Syndrome | 147920 | https://app.face2gene.com/lmd/893/photo/13115 | 3 | 16 |
| 288 | Noonan Syndrome | 163950 | https://app.face2gene.com/lmd/4716/photo/14088 | 22 | 19 |
| 289 | Noonan Syndrome | 163950 | https://app.face2gene.com/lmd/4716/photo/14090 | 3 | 2 |
| 290 | Noonan Syndrome | 163950 | https://app.face2gene.com/lmd/4716/photo/14091 | 1 | 4 |
| 291 | Noonan Syndrome | 163950 | https://app.face2gene.com/lmd/4716/photo/14092 | 6 | 3 |
| 292 | KBG Syndrome; KBGS | 148050 | https://app.face2gene.com/lmd/912/photo/15603 | 1 | 1 |
| 293 | Opitz GBBB Syndrome, Type II; GBBB2 | 145410 | https://app.face2gene.com/lmd/640/photo/15945 | 3 | 20 |
| 294 | Gapo Syndrome | 230740 | https://app.face2gene.com/lmd/642/photo/16435 | 9 | 13 |
| 295 | Velocardiofacial Syndrome | 192430 | https://app.face2gene.com/lmd/1762/photo/19049 | 9 | 7 |
| 296 | Velocardiofacial Syndrome | 192430 | https://app.face2gene.com/lmd/1762/photo/5395 | 80 | 16 |
| 297 | Trichorhinophalangeal Syndrome | 190350 | https://app.face2gene.com/lmd/1730/photo/5284 | 2 | 2 |
| 298 | Blepharophimosis, Ptosis, and Epicanthus Inversus; BPES | 110100 | https://app.face2gene.com/lmd/202/photo/581 | 44 | 16 |
| 299 | Noonan Syndrome | 163950 | https://app.face2gene.com/lmd/1229/photo/3582 | 49 | 31 |
| 300 | Craniofrontonasal Syndrome; CFNS | 304110 | https://app.face2gene.com/lmd/376/photo/1114 | 1 | 2 |
| 301 | Trichorhinophalangeal Syndrome | 190350 | https://app.face2gene.com/lmd/1730/photo/5282 | 1 | 1 |
| 302 | Alagille Syndrome | 118450 | https://app.face2gene.com/lmd/110/photo/349 | 1 | 1 |
| 303 | Smith-Lemli-Opitz Syndrome; SLOS | 270400 | https://app.face2gene.com/lmd/1608/photo/12105 | 13 | 60 |
| 304 | Opitz GBBB Syndrome, Type II; GBBB2 | 145410 | https://app.face2gene.com/lmd/640/photo/15943 | 2 | 4 |
| 305 | Lubs X-Linked Mental Retardation Syndrome; MRXSL | 300260 | https://app.face2gene.com/lmd/5107/photo/16174 | 10 | 17 |
| 306 | Phelan-Mcdermid Syndrome; PHMDS | 606232 | https://app.face2gene.com/lmd/4459/photo/16512 | 1 | 2 |
| 307 | Cornelia de Lange Syndrome 1; CDLS1 | 122470 | https://app.face2gene.com/lmd/423/photo/17342 | 1 | 1 |
| 308 | Craniometaphyseal Dysplasia | 123000 | https://app.face2gene.com/lmd/371/photo/7667 | 7 | 6 |
| 309 | Coffin-Lowry Syndrome; CLS | 303600 | https://app.face2gene.com/lmd/328/photo/17870 | 1 | 12 |
| 310 | Coffin-Lowry Syndrome; CLS | 303600 | https://app.face2gene.com/lmd/328/photo/17868 | 1 | 1 |
| 311 | Coffin-Lowry Syndrome; CLS | 303600 | https://app.face2gene.com/lmd/328/photo/17868 | 17 | 17 |
| 312 | Branchiooculofacial Syndrome; BOFS | 113620 | https://app.face2gene.com/lmd/633/photo/9227 | 32 | 22 |
| 313 | Smith-Magenis Syndrome; SMS | 182290 | https://app.face2gene.com/lmd/3562/photo/18747 | 3 | 21 |
| 314 | Oculodentodigital Dysplasia | 164200 | https://app.face2gene.com/lmd/1246/photo/19085 | 3 | 4 |
| 315 | Birk-Barel Mental Retardation Dysmorphism Syndrome | 612292 | https://app.face2gene.com/lmd/5569/photo/19317 | 16 | 15 |
| 316 | Blepharophimosis, Ptosis, and Epicanthus Inversus; BPES | 110100 | https://app.face2gene.com/lmd/202/photo/580 | 91 | 17 |
| 317 | Craniometaphyseal Dysplasia | 123000 | https://app.face2gene.com/lmd/371/photo/41763 | 1 | 4 |
| 318 | Rubinstein-Taybi Syndrome | 180849 | https://app.face2gene.com/lmd/1490/photo/41843 | 1 | 1 |
| 319 | Seckel Syndrome | 210600 | https://app.face2gene.com/lmd/739/photo/41855 | 26 | 23 |
| 320 | Prader-Willi Syndrome; PWS | 176270 | https://app.face2gene.com/lmd/1388/photo/41911 | 2 | 7 |

| | | | | | |
|---|---|---|---|---|---|
| 321 | Crouzon Syndrome | 123500 | https://app.face2gene.com/lmd/383/photo/41942 | 6 | 14 |
| 322 | Treacher Collins Syndrome | 154500 | https://app.face2gene.com/lmd/1720/photo/41957 | 18 | 31 |
| 323 | Frontometaphyseal Dysplasia; FMD | 305620 | https://app.face2gene.com/lmd/622/photo/41569 | 1 | 1 |

**Supplementary Table 4: Results of the GeneMatcher validation set using Enc-healthy.**

| Gene | PMID | Total families (Subjects) | Connected families [a] (subjects) | |
|---|---|---|---|---|
| | | | Top-10 | Top-30 |
| *BPTF* | 28942966 | 6 (6) | 4 (4) | 6 (6) |
| *CCDC47* | 30401460 | 4 (4) | 0 (0) | 0 (0) |
| *CHAMP1* | 27148580 | 4 (4) | 2 (2) | 2 (2) |
| *CHD4* | 27616479 | 3 (3) | 0 (0) | 0 (0) |
| *DDX6* | 31422817 | 4 (4) | 2 (4) | 2 (4) |
| *EBF3* | 28017373 | 6 (7) | 2 (2) | 2 (2) |
| *FBXO11* | 30679813 | 17 (17) | 0 (0) | 10 (10) |
| *HNRNPK* | 26173930 | 3 (3) | 0 (0) | 0 (0) |
| *KDM3B* | 30929739 | 9 (9) | 0 (0) | 0 (0) |
| *LEMD2* | 30905398 | 2 (2) | 0 (0) | 2 (2) |
| *OTUD6B* | 28343629 | 4 (9) | 0 (0) | 0 (0) |
| *PACS2* | 29656858 | 6 (6) | 0 (0) | 0 (0) |
| *TMEM94* | 30526868 | 6 (10) | 5 (7) | 6 (10) |
| *WDR37* | 31327508 | 4 (4) | 0 (0) | 2 (2) |
| *ZNF148* | 27964749 | 3 (3) | 2 (2) | 2 (2) |
| Total | - | 79 (91) | 17 (21) | 34 (40) |
| Average | - | - | 21.52% (23.08%) | 43.04% (43.96%) |

[a] Number of families matched by a photo from another family in the top-10 or top-30 rank.

**Supplementary Table 5: Summary of all the encoders.**

| Name | Training set | Number of syndromes | Evaluation method |
|---|---|---|---|
| Enc-healthy | CASIA-WebFace | 0 | cosine |
| Enc-F2G (softmax) | Face2Gene frequent set | 299 | softmax |
| Enc-F2G | Face2Gene frequent set | 299 | cosine |
| Enc-F2G-rare | Face2Gene rare set (syndromes with more than 2 subjects) | 467 | cosine |
| Enc-F2G-exclude-50 | Face2Gene frequent set excluding 50 selected syndromes | 249 | cosine |
| Enc-GMDB (softmax) | GMDB frequent set | 139 | softmax |
| Enc-GMDB | GMDB frequent set | 139 | cosine |
| Enc-GMDB-exclude-20 | GMDB frequent set excluding 20 selected syndromes | 119 | cosine |

### Supplementary Table 6: The 50 selected syndromes used in the random downsampling experiment, sorted by top-10 accuracy.

| ID | Syndrome | Top-10 | Grade[a] | Prevalence[b] | Subjects |
|---|---|---|---|---|---|
| **1** | Hutchinson-Gilford Progeria Syndrome; HGPS | 93.33 | 2.33 | 0.005 | 60 |
| **2** | Nijmegen Breakage Syndrome; NBS | 82.2 | 2 | 1 | 30 |
| **3** | Blepharophimosis, Ptosis, and Epicanthus Inversus; BPES | 81.95 | 3 | 0.0001 | 42 |
| **4** | Coffin-Lowry Syndrome; CLS | 81.78 | 2 | 1.5 | 133 |
| **5** | Schuurs-Hoeijmakers syndrome; SHMS | 80.13 | 2 | 2.564E-05 | 25 |
| **6** | Sotos Syndrome | 77.42 | 2.67 | 7.1 | 215 |
| **7** | Baraitser-Winter Syndrome | 77.36 | 2.67 | 0.00077 | 63 |
| **8** | Branchiooculofacial Syndrome; BOFS | 77.08 | 2 | 0.00192 | 36 |
| **9** | Williams-Beuren Region Duplication Syndrome | 75.86 | 1.33 | 0.00209 | 52 |
| **10** | Williams-Beuren Syndrome; WBS | 75.5 | 3 | 10.8 | 457 |
| **11** | Barth Syndrome; BTHS | 74.77 | 1.33 | 0.22 | 24 |
| **12** | Crouzon Syndrome | 73.5 | 2.33 | 0.9 | 73 |
| **13** | Seckel Syndrome | 72.31 | 2.67 | 0.2 | 37 |
| **14** | Campomelic Dysplasia | 70.24 | 1.33 | 0.33 | 19 |
| **15** | Aarskog-Scott syndrome ; AAS | 69.82 | 2 | 0.5 | 82 |
| **16** | Wolf-Hirschhorn Syndrome; WHS | 69.27 | 2.67 | 2 | 105 |
| **17** | Oculodentodigital Dysplasia | 69.17 | 2.33 | 0.00312 | 62 |
| **18** | Rothmund-Thomson Syndrome; RTS | 69.16 | 2 | 0.00513 | 42 |
| **19** | Ectrodactyly, Ectodermal Dysplasia, and Cleft Lip/palate Syndrome 1; EEC1 | 67.83 | 2 | 1.11 | 18 |
| **20** | Renpenning Syndrome 1; RENS1 | 67.77 | 2.33 | 0.00082 | 32 |
| **21** | Cornelia De Lange Syndrome | 66.67 | 3 | 1.9 | 479 |
| **22** | Mucopolysaccharidosis Type VI; MPS6 | 66.67 | 2 | 0.16 | 42 |
| **23** | Weaver Syndrome; WVS | 66.41 | 2 | 0.00062 | 37 |
| **24** | Mucolipidosis III Alpha/beta | 66.09 | 2 | 13 | 27 |
| **25** | Hyper-IgE Recurrent Infection Syndrome | 66 | 1 | 0.1 | 24 |
| **26** | Pycnodysostosis | 65.35 | 2 | 0.13 | 21 |
| **27** | Mental Retardation X-Linked 102; MRX102 | 65.25 | 1 | 0.00049 | 43 |
| **28** | Laron Syndrome | 65.13 | 1.67 | 0.3 | 24 |
| **29** | Myotonic Dystrophy | 64.9 | 1.33 | 6.7 | 30 |
| **30** | Koolen-de Vries Syndrome; KDVS | 63.5 | 2.33 | 1.82 | 103 |
| **31** | Opitz GBBB Syndrome, Type II; GBBB2 | 63.01 | 2 | 3 | 30 |
| **32** | Hyperphosphatasia with Mental Retardation Syndrome | 61.6 | 2 | 0.00031 | 62 |
| **33** | Mucopolysaccharidosis, Type IIIA; MPS3A | 57.94 | 2 | 0.32 | 27 |
| **34** | Alagille Syndrome | 57.93 | 1.33 | 0.8 | 80 |
| **35** | Johanson-Blizzard Syndrome; JBS | 57.69 | 2 | 0.4 | 12 |
| **36** | Ehlers-Danlos syndrome, vascular type; EDSVASC | 57.55 | 1.67 | 1 | 34 |
| **37** | Filippi Syndrome; FLPIS | 56.64 | 1.67 | 0.00037 | 23 |
| **38** | Holt-Oram Syndrome; HOS | 55.56 | 1 | 0.7 | 35 |
| **39** | Neurofibromatosis, Type I; NF1 | 54.01 | 1 | 21.3 | 106 |
| **40** | Focal Dermal Hypoplasia; FDH | 52.54 | 1.33 | 0.00385 | 45 |
| **41** | Hallermann-Streiff Syndrome; HSS | 52.46 | 2.67 | 0.00192 | 30 |
| **42** | Miller-Dieker Lissencephaly Syndrome; MDLS | 49.78 | 1.67 | 1 | 33 |
| **43** | Larsen Syndrome; LRS | 46.43 | 1.67 | 0.4 | 38 |

| | | | | | |
|---|---|---|---|---|---|
| **44** | Tetrasomy 18p | 46.34 | 1 | 0.03205 | 44 |
| **45** | Lubs X-Linked Mental Retardation Syndrome; MRXSL | 46.08 | 1.67 | 0.00256 | 70 |
| **46** | Sturge-Weber Syndrome; SWS | 45.65 | 2.33 | 3.5 | 37 |
| **47** | Joubert Syndrome | 43.92 | 1.33 | 1.125 | 56 |
| **48** | Waardenburg Syndrome | 42.97 | 3 | 0.37 | 99 |
| **49** | Trisomy 18 Syndrome | 37.17 | 2 | 16.7 | 25 |
| **50** | Turner Syndrome | 34.81 | 1 | 5.5 | 99 |

[a] Average of the scores from three clinicians.

[b] Obtained from Orphanet; prevalence is per 100,000 population.

### Supplementary Table 7: The 20 selected syndromes from GMDB used in the random downsampling experiment, sorted by top-5 accuracy.

| ID | Syndrome | Top-5 | Grade [a] | Subjects |
|---|---|---|---|---|
| **1** | Hutchinson-Gilford progeria | 81.29 | 2.83 | 44 |
| **2** | COFFIN-LOWRY SYNDROME; CLS | 79.46 | 2.33 | 26 |
| **3** | Rubinstein-Taybi syndrome | 78.21 | 2 | 35 |
| **4** | OCULODENTODIGITAL DYSPLASIA; ODDD | 76.19 | 2.83 | 26 |
| **5** | Hyperphosphatasia with mental retardation syndrome | 76.14 | 2.33 | 52 |
| **6** | BLEPHAROPHIMOSIS, PTOSIS, AND EPICANTHUS INVERSUS; BPES | 72.54 | 3 | 22 |
| **7** | RENPENNING SYNDROME 1; RENS1 | 71.96 | 2 | 19 |
| **8** | Alagille syndrome | 71.02 | 1 | 22 |
| **9** | SCHUURS-HOEIJMAKERS SYNDROME; SHMS | 66.66 | 2.16 | 18 |
| **10** | WEAVER SYNDROME; WVS | 61.71 | 2 | 23 |
| **11** | INTELLECTUAL DEVELOPMENTAL DISORDER, X-LINKED, SYNDROMIC, SNIJDERS BLOK TYPE; MRXSSB | 61.61 | 1.33 | 28 |
| **12** | Baraitser-Winter syndrome | 56.61 | 2.46 | 50 |
| **13** | NIJMEGEN BREAKAGE SYNDROME; NBS | 56.60 | 1.66 | 20 |
| **14** | Meier-Gorlin syndrome | 54.54 | 2 | 27 |
| **15** | MENTAL RETARDATION, AUTOSOMAL DOMINANT 21; MRD21 | 54.12 | 1.33 | 23 |
| **16** | FOCAL DERMAL HYPOPLASIA; FDH | 53.53 | 1.33 | 16 |
| **17** | LARSEN SYNDROME; LRS | 51.51 | 1.66 | 21 |
| **18** | Sotos syndrome | 50.98 | 2.43 | 53 |
| **19** | MENTAL RETARDATION, AUTOSOMAL DOMINANT 31; MRD31 | 50 | 1 | 26 |
| **20** | Joubert syndrome | 35.23 | 1 | 22 |

[a] Average of the scores from three clinicians.

### Supplementary Table 8: Comparison of different models for matching target syndromes.

| Model | Method | Images | | Syndromes | Null top-1 accuracy | Top 1 | Top 5 | Top 10 | Top 30 |
|---|---|---|---|---|---|---|---|---|---|
| | | Gallery | Test | | | | | | |
| Enc-F2G | cosine | 2073.6 | 684.4 | 467 | 0.21% | 11.54% | 21.29% | 27.41% | 40.50% |
| Enc-F2G-rare | cosine | 2073.6 | 684.4 | 467 | 0.21% | **11.78%** | **21.77%** | **28.42%** | **42.69%** |
| Enc-F2G-rare | softmax | - | 649 | 467 | 0.21% | 11.73% | 19.20% | 23.58% | 36.86% |

Enc-F2G is the encoder trained on 299 frequent syndromes from Face2Gene dataset, and Enc-F2G-rare is the encoder trained on 467 rare syndromes that contain from three to six subjects. The table shows the results of using different models and methods to match patients with rare syndromes. The first two rows are the average of 10-fold cross-validation, and the last row is the result using the softmax method of the DeepGestalt model to predict the syndrome, so it did not use the gallery to match images. To evaluate the same Enc-F2G-rare model and make sure the test images were not trained in Enc-F2G-rare, we did not perform 10-fold cross-validation for the softmax method. The best performance among the three conditions is boldfaced.

The encoder trained on target syndromes, Enc-F2G-rare, provided the highest top-1 to top-30 accuracy when cosine distance was used to define proximity; it even outperformed the trained softmax classifier. Enc-F2G, which trained on frequent syndromes and did not see any of the rare syndromes during its training, showed results very similar to those of Enc-F2G-rare and is therefore a good choice for frequent syndromes because it would not require retraining the encoder. Moreover, training the model on rare syndromes, which have very few high-quality photos, in addition to frequent syndromes might lead to poor performance due to the extremely imbalanced training dataset.

**Supplementary Table 9: Summary of Face2Gene and GMDB datasets.**

Please find it in the attachment.
